# Somatic instability of the *FGF14*-SCA27B GAA•TTC repeat reveals a marked expansion bias in the cerebellum

**DOI:** 10.1101/2024.07.01.24309777

**Authors:** David Pellerin, Jean-Loup Méreaux, Susana Boluda, Matt C. Danzi, Marie-Josée Dicaire, Claire-Sophie Davoine, David Genis, Guinevere Spurdens, Catherine Ashton, Jillian M. Hammond, Brandon J. Gerhart, Viorica Chelban, Phuong U. Le, Maryam Safisamghabadi, Christopher Yanick, Hamin Lee, Sathiji K. Nageshwaran, Gabriel Matos-Rodrigues, Zane Jaunmuktane, Kevin Petrecca, Schahram Akbarian, André Nussenzweig, Karen Usdin, Mathilde Renaud, Céline Bonnet, Gianina Ravenscroft, Mario A. Saporta, Jill S. Napierala, Henry Houlden, Ira W. Deveson, Marek Napierala, Alexis Brice, Laura Molina Porcel, Danielle Seilhean, Stephan Zuchner, Alexandra Durr, Bernard Brais

**Affiliations:** Dr. John T. Macdonald Foundation Department of Human Genetics and John P. Hussman Institute for Human Genomics, University of Miami Miller School of Medicine, Miami, FL, USA; Department of Neurology and Neurosurgery, Montreal Neurological Hospital and Institute, McGill University, Montreal, QC, Canada; Department of Neuromuscular Diseases, UCL Queen Square Institute of Neurology and The National Hospital for Neurology and Neurosurgery, University College London, London, United Kingdom; Sorbonne Université, Institut du Cerveau - Paris Brain Institute- ICM, Inserm, CNRS, APHP, University Hospital Pitié-Salpêtrière, Paris, France; Ataxia and Hereditary Spastic Paraplegia Unit, Service of Neurology, Hospital Universitari de Girona Dr. Josep Trueta (ICS) & Hospital Santa Caterina IAS, Institut d’Investigació Biomèdica de Girona (IDIBGI), Girona, Spain; Department of Neurology, Royal Perth Hospital, Perth, WA, Australia; Genomics and Inherited Disease Program, Garvan Institute of Medical Research, Sydney, NSW, Australia; Centre for Population Genomics, Garvan Institute of Medical Research and Murdoch Children’s Research Institute, Australia; Department of Neurology, Peter O’Donnell Jr. Brain Institute, University of Texas Southwestern Medical Center, Dallas, TX, USA; Neurobiology and Medical Genetics Laboratory, “Nicolae Testemitanu” State University of Medicine and Pharmacy, Chisinau, Republic of Moldova; Department of Neurology, University of Miami Miller School of Medicine, Miami, FL, USA; Department of Psychiatry, Department of Neuroscience and Department of Genetics and Genomic Sciences, Friedman Brain Institute Icahn School of Medicine at Mount Sinai, New York, NY, USA; Neurogenetics Program, Department of Neurology, David Geffen School of Medicine, University of California Los Angeles, Los Angeles, CA, USA; Laboratory of Genome Integrity, National Cancer Institute, NIH, Bethesda, MD, USA; Division of Neuropathology, The National Hospital for Neurology and Neurosurgery, University College London NHS Foundation Trust, London, United Kingdom; Department of Clinical and Movement Neurosciences and Queen Square Brain Bank for Neurological Disorders, UCL Queen Square Institute of Neurology, University College London, London, United Kingdom; Laboratory of Cell and Molecular Biology, National Institute of Diabetes and Digestive and Kidney Diseases, National Institutes of Health, Bethesda, MD, USA; INSERM-U1256 NGERE, Université de Lorraine, Nancy, France; Service de Neurologie, CHRU de Nancy, Nancy, France; Service de Génétique Clinique, CHRU de Nancy, Nancy, France; Laboratoire de Génétique, CHRU de Nancy, Nancy, France; Centre for Medical Research University of Western Australia and Harry Perkins Institute of Medical Research, Perth, Western Australia, Australia; Faculty of Medicine, University of New South Wales, Sydney, NSW, Australia; Alzheimer’s Disease and other Cognitive Disorders Unit, Service of Neurology, Hospital Clínic, Fundació de Recerca Clínic Barcelona-Institut d’Investigacions Biomediques August Pi i Sunyer (FRCB-IDIBAPS), University of Barcelona, Barcelona, Spain; Neurological Tissue Brain Bank, Biobanc-Hospital Clínic-FRCB-IDIBAPS, Barcelona, Spain; Department of Human Genetics, McGill University, Montreal, QC, Canada

**Keywords:** SCA27B, spinocerebellar ataxia 27B, GAA-FGF14 ataxia, repeat expansion disorder, mosaicism, expansion

## Abstract

Spinocerebellar ataxia 27B (SCA27B) is a common autosomal dominant ataxia caused by an intronic GAA•TTC repeat expansion in *FGF14*. Neuropathological studies have shown that neuronal loss is largely restricted to the cerebellum. Although the repeat locus is highly unstable during intergenerational transmission, it remains unknown whether it exhibits cerebral mosaicism and progressive instability throughout life. We conducted an analysis of the *FGF14* GAA•TTC repeat somatic instability across 156 serial blood samples from 69 individuals, fibroblasts, induced pluripotent stem cells, and post-mortem brain tissues from six controls and six patients with SCA27B, alongside methylation profiling using targeted long-read sequencing. Peripheral tissues exhibited minimal somatic instability, which did not significantly change over periods of more than 20 years. In post-mortem brains, the GAA•TTC repeat was remarkably stable across all regions, except in the cerebellar hemispheres and vermis. The levels of somatic expansion in the cerebellar hemispheres and vermis were, on average, 3.15 and 2.72 times greater relative to other examined brain regions, respectively. Additionally, levels of somatic expansion in the brain increased with repeat length and tissue expression of *FGF14*. We found no significant difference in methylation of wild-type and expanded *FGF14* alleles in post-mortem cerebellar hemispheres between patients and controls. In conclusion, our study revealed that the *FGF14* GAA•TTC repeat exhibits a cerebellar-specific expansion bias, which may explain the pure and late-onset cerebellar involvement in SCA27B.

## Introduction

Spinocerebellar ataxia 27B (SCA27B; GAA-*FGF14*-related ataxia) is a recently described autosomal dominant ataxia caused by the expansion of a polymorphic GAA•TTC repeat located within intron 1 of the fibroblast growth factor 14 (*FGF14*) gene.^1, 2^ SCA27B is phenotypically characterized by a late-onset, slowly progressive pan-cerebellar syndrome that is frequently associated with episodic symptoms and downbeat nystagmus.^1, 3–5^ The size of the GAA•TTC repeat expansion only shows a weak negative correlation with the age at onset^1, 6^ and has not been associated with disease severity or progression.^3, 4^ Furthermore, there exists significant phenotypic variability among patients carrying expansions of similar sizes, suggesting that additional factors influence disease expressivity.^3, 4, 6^ Neuropathological studies in SCA27B have shown that neuronal loss is largely restricted to the cerebellar cortex, involving the vermis more than the hemispheres.^1, 3^ However, it is noteworthy that significant pathological abnormalities are not found in other regions despite the widespread expression of *FGF14* in the brain, albeit at lower levels than in the cerebellum.^7^ This observation suggests that tissue-specific factors may increase the vulnerability of the cerebellum in SCA27B.

Although the expanded *FGF14* GAA•TTC repeat is highly unstable upon intergenerational transmission,^4, 8^ whether it exhibits somatic instability in the brain has not yet been investigated. Somatic mosaicism is a phenomenon observed in many repeat expansions, which have the ability to form alternative (non-B) DNA structures that predispose them to errors during DNA repair.^9–11^ The degree of somatic instability generally increases with the length of the repeat tract and age, and also shows a repeat-specific pattern of variation across different tissues and cell types.^12^ Remarkably, previous studies on coding CAG•CTG repeat expansions in SCA1, SCA2, SCA3, SCA7, and dentatorubral pallidoluysian atrophy (DRPLA) have shown that these diseases consistently have smaller expansion sizes and lower degrees of mosaicism in the cerebellum compared to other brain regions.^13–17^ In comparison, the intronic ATTTC pentanucleotide repeat insertion in *DAB1* exhibits longer sizes and an expansion bias in the cerebellum compared to blood samples and fibroblasts in patients with SCA37.^18^ The intronic *RFC1* AAGGG•CCCTT repeat expansion, however, shows little instability in the cerebellum of patients with cerebellar ataxia, neuropathy, and vestibular areflexia syndrome (CANVAS).^19^ Furthermore, in Friedreich ataxia, although the expanded intronic *FXN* GAA•TTC repeat is longer in the cerebellum than in other brain regions, large contractions occur significantly more frequently than large expansions.^20–23^

Here, to gain further insight into SCA27B biology, we performed a comprehensive analysis of the *FGF14* GAA•TTC repeat somatic instability in serial blood samples, fibroblasts, induced pluripotent stem cells (iPSCs), and post-mortem brains from individuals with SCA27B.

## Methods

Detailed methods are provided in the Supporting Information.

### Human sample collection

Human post-mortem brain tissue was collected from six non-ataxic control individuals and six persons with SCA27B to study the somatic stability of the *FGF14* GAA•TTC repeat tract (**Table 1**). Fresh frozen samples were obtained from various regions of the central nervous system (CNS), as detailed in **Supplementary Table 1**. Additionally, DNA extracted from blood and cerebellar hemispheres of another eight control individuals was obtained to compare allele sizes between both tissues. We also collected fresh frozen tissue of the cerebellar hemispheres from an additional four controls and one SCA27B patient to perform methylation profiling of *FGF14* with Oxford Nanopore Technologies (ONT), as detailed in the Supplementary Methods.

**Table 1:**
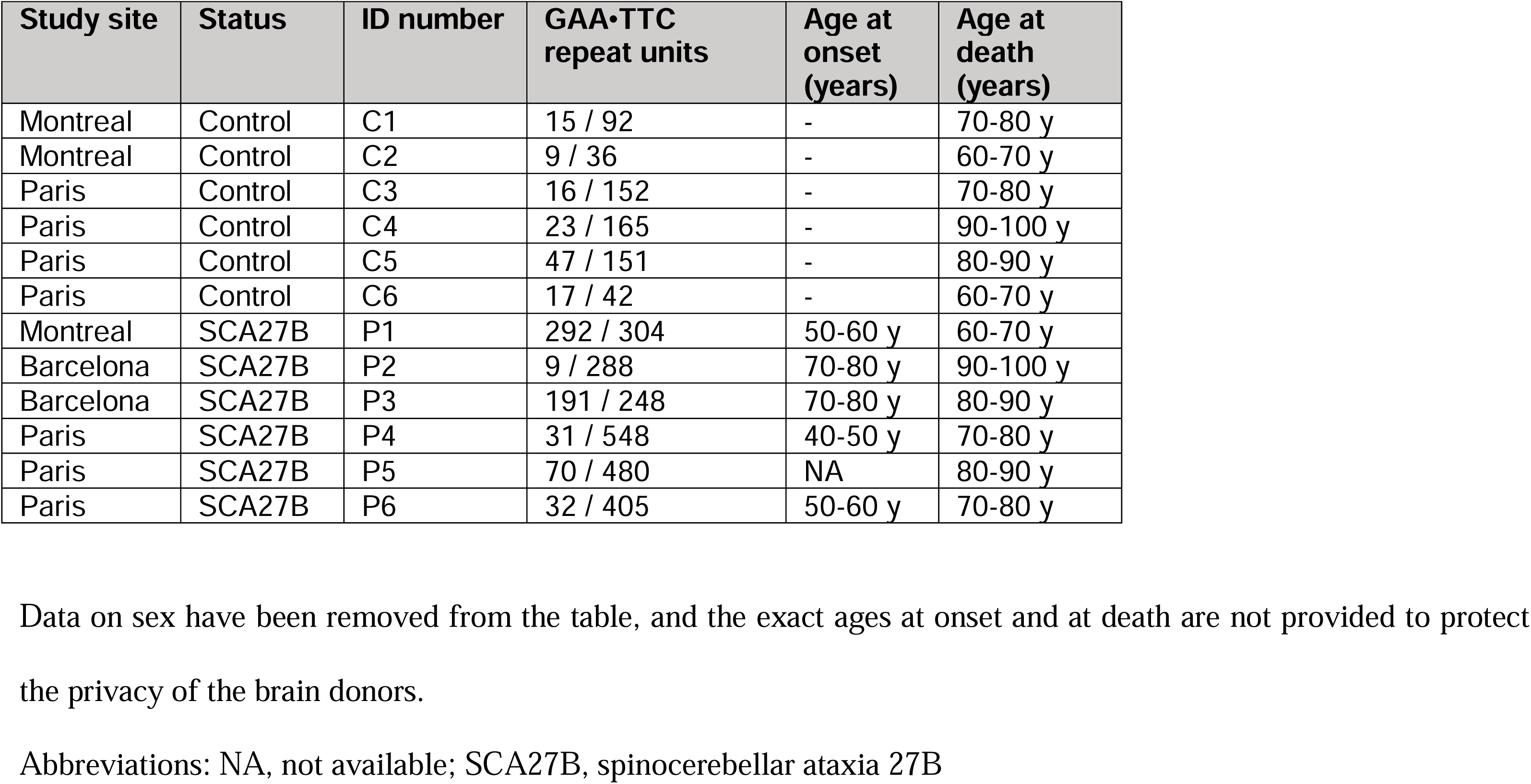
Demographic and genetic characteristics of the post-mortem brain samples analyzed for *FGF14* GAA•TTC repeat instability.

### Analysis of somatic instability

Somatic stability was studied by capillary electrophoresis or by agarose gel electrophoresis for samples carrying alleles >400-450 triplets. For samples analyzed by capillary electrophoresis, the GeneMapper software v6.0 (Applied Biosystems) or Peak Scanner software v1.0 (Applied Biosystems) was used to establish the ‘modal peak’ of each of the two alleles in each sample, which was defined as the peak with the highest height. Peaks to the right of the modal allele represent somatically expanded GAA•TTC repeats, while peaks to the left may include both contracted GAA•TTC repeats and PCR stutter products. Expansion indices (EI) were calculated by only taking into account peaks to the right of the ‘modal allele’ and using a 1% relative peak threshold (**Figure 1**), as described previously.^13, 24^ The EI was obtained by first dividing the height of each individual peak by the sum of the heights of all peaks above the threshold. The resulting normalized peak heights were then multiplied by the position of the peak and these normalized peak values were summed to generate the EI. Analysis of blood and CNS tissues revealed that the modal length was remarkably stable across all regions examined, except in the cerebellar hemispheres and vermis where it showed a tendency to expand. We therefore considered the modal allele length as measured in the blood and most CNS regions as the constitutional allele length, which we set as the main allele for calculation of the EI in tissues with an expansion shift of the modal repeat,^13^ tolerating a discrepancy of one triplet due to the variability of our genotyping assay.^25^ Finally, we correlated the somatic stability indices with the expression of *FGF14* in normal tissue data from the Human Protein Atlas (HPA v.23.0; proteinatlas.org accessed 27/03/2024).^26^

**Figure 1:**
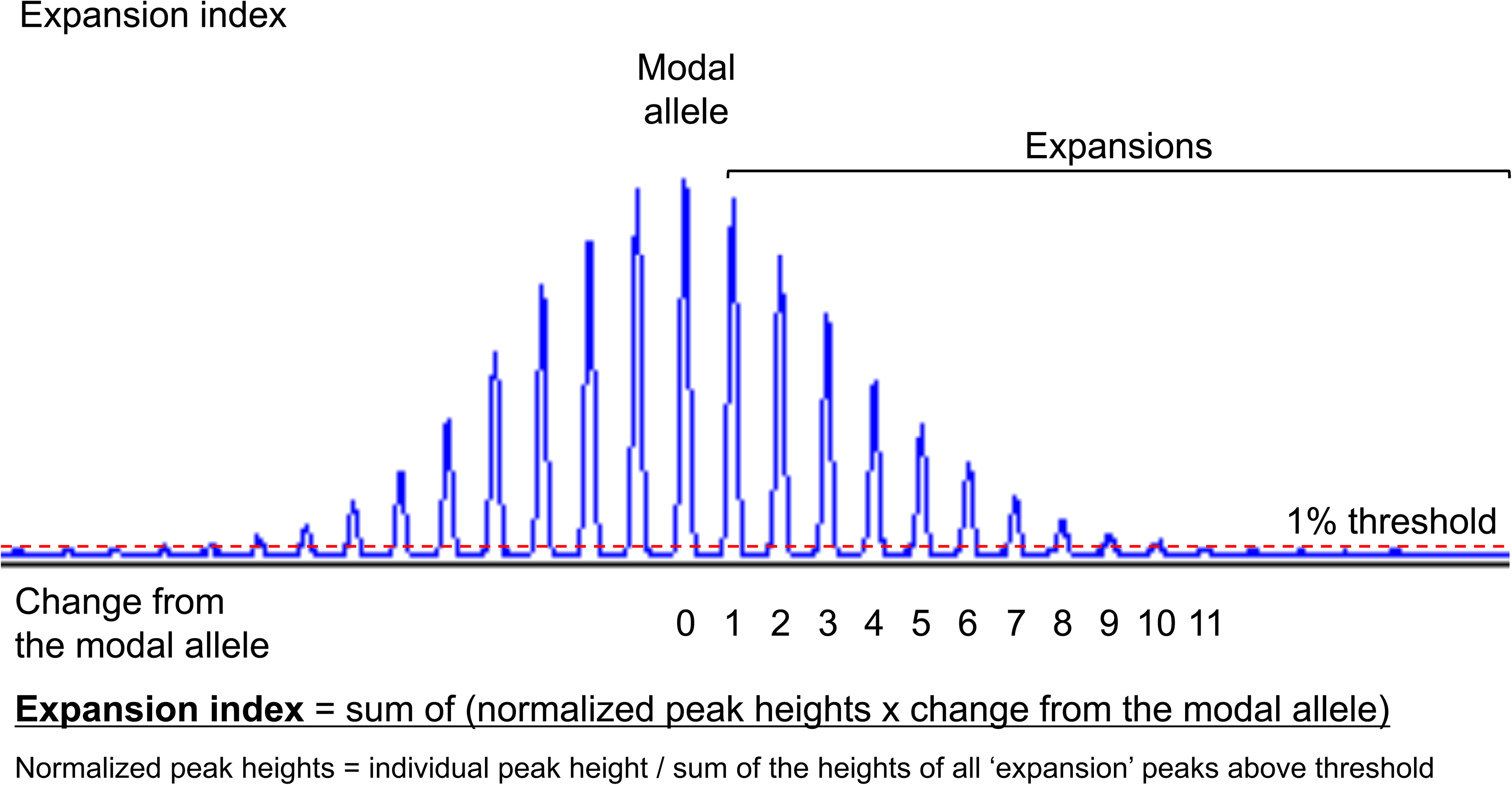
Determination of the *FGF14* GAA•TTC repeat expansion index. Method for calculating the expansion index of the *FGF14* GAA•TTC repeat. The expansion index is calculated by only taking into account peaks to the right of the modal allele, which represent somatically expanded GAA•TTC repeats, and using a 1% relative peak threshold. The modal allele corresponds to the peak with the highest intensity, as measured in relative fluorescence units in GeneMapper or Peak Scanner. The expansion index is calculated by first dividing the height of each individual peak by the sum of the heights of all peaks to the right of the modal allele that are above the set 1% threshold. The resulting normalized peak heights are then multiplied by the position of the peak (change from the modal allele) and these values are summed to generate the index.

### Ethics

The institutional review boards of the Montreal Neurological Hospital, Montreal (MPE-CUSM-15-915), the Centre Hospitalier de l’Université de Montréal, Montreal (ND02.045), the Dr. Josep Trueta University Hospital, Girona (2022.120), the Hôpital Pitié-Salpêtrière, Paris (SPATAX RBM 01-29 and RBM 03-48, BIOMOV NCT05034172), and the University College London Hospitals, London (04/N034) approved this study. All brain donors signed an informed consent for brain tissue usage in medical research. The study complied with all relevant ethical regulations.

### Data availability

The data supporting this study may be shared at the request of any qualified investigator on reasonable request.

## Results

### Longitudinal analysis of the FGF14 GAA•TTC repeat somatic instability in blood samples

We performed a longitudinal analysis to determine whether the length of the *FGF14* repeat tracts changes over time in 156 serial blood samples from 69 individuals (see Supplementary Results). Alleles longer than ≈400-450 triplets could not be measured by capillary electrophoresis and were not included in this analysis. The median interval between the first and last blood sample was 8.9 years (interquartile range [IQR]: 2.9 to 13.9). We observed no significant change of the modal *FGF14* repeat size between the first and last blood samples (**Figure 2a**; median difference [size of last sample – size of first sample], 0 triplet; IQR: 0 to 0; Sign test, *p*=0.22), and found that elapsed time had no significant effect on the modal repeat size as assessed by a linear mixed-effects model (estimate = - 0.441, standard error [SE] = 0.678, *p*=0.52).

**Figure 2:**
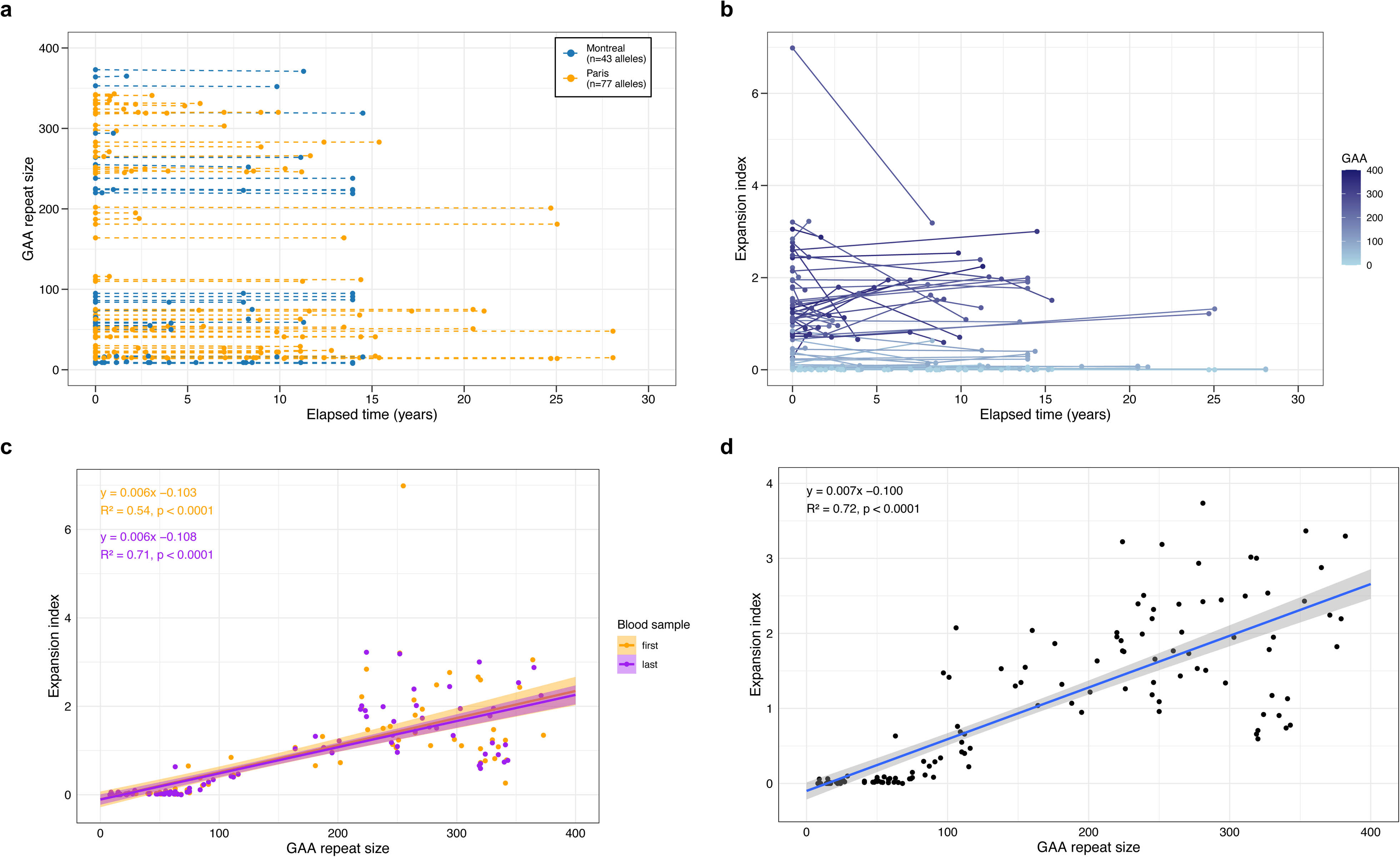
Longitudinal analysis of the *FGF14* GAA•TTC repeat somatic stability in blood samples. Longitudinal analysis of (a) the *FGF14* GAA•TTC repeat tract size, expressed in triplet repeat counts, and (b) expansion index across 69 individuals (120 alleles analyzed) who underwent serial blood collections over a median period of 8.9 years (interquartile range [IQR]: 2.9 to 13.9). Observations from the same person are connected by a line. In panel b, the color gradient shows the GAA•TTC allele size of each data point. (c) Positive linear relationship between the *FGF14* GAA•TTC repeat tract size, expressed in triplet repeat counts, and expansion index in each of the first (orange) and last (purple) longitudinal blood samples across 69 individuals (*n*=120 alleles). The Pearson’s correlation coefficient is r = 0.73 (95% confidence interval [CI]: 0.64 to 0.81) for the first blood samples and r = 0.84 (95% CI: 0.78 to 0.89) for the last blood samples. Both regression lines did not differ significantly (mixed-effect analysis, *p*=0.72), indicating relative stability of the expansion index over time. (d) Positive linear relationship between the *FGF14* GAA•TTC repeat tract size and expansion index across 100 individuals (*n*=173 alleles), including the results from the last blood samples of the 69 individuals featured in panels b and c. The Pearson’s correlation coefficient is r = 0.85 (95% CI: 0.80 to 0.88). In panels c and d, the shaded areas display the 95% confidence intervals.

As for repeat sizes, the EI did not significantly change over time in repeated blood samples (**Figure 2b-c**, median of differences [EI of last sample – EI of first sample], 0 [IQR: −0.02 to 0.01]; Sign test, *p*=0.22), nor in the subset of alleles longer than 200 triplets (median of differences, 0 [IQR: −0.28 to 0.43]), Sign test, *p*=1). In a linear mixed-effects model, we found that allele size (estimate = 5.530e-03, SE = 3.253e-04, *p*<0.0001) had a significant effect on the EI, while elapsed time (estimate = −7.634e-03, SE = 6.945e-03, *p*=0.27), the interaction between allele size and elapsed time (estimate = 9.008e-05, SE = 4.839e-05, *p*=0.064), and sex (estimate = −5.148e-02, SE = 9.654e-02, *p*=0.60) did not. Regression analysis conducted on 173 alleles from 100 individuals (the EI of alleles longer than ≈400-450 triplets could not be adequately determined by capillary electrophoresis and were not included in this analysis) confirmed a linear increase in somatic expansion with *FGF14* GAA•TTC repeat size (r=0.85 [95% confidence interval: 0.80 to 0.88], slope = 0.007, *p*<0.0001) (**Figure 2d**). Furthermore, we compared the size and EI of the *FGF14* repeats in two pairs of monozygotic twins with SCA27B who had blood samples drawn at the age of 65-70 and 60-65 years, respectively. We observed a difference of a single triplet between them (15/225 vs 15/226 and 9/427 vs 9/428 triplets, respectively), which falls within the expected range of variability of our genotyping assay.^25^ The short alleles of both pairs of twins did not exhibit somatic expansion (EI=0), while the EI of the long allele of the first pair of twins differed by only 0.5 (EI = 1.76 vs 1.26; the EI of the long allele of the second pair of twins could not be calculated due to their large size). Overall, our results demonstrate that *FGF14* GAA•TTC repeat size and somatic expansion in blood samples remain largely stable over periods of more than 20 years.

### Instability of the FGF14 GAA•TTC repeat tract in fibroblasts and induced pluripotent stem cells

To further assess somatic instability of the *FGF14* GAA•TTC repeat in peripheral tissues, we analyzed allele sizes in paired blood and fibroblasts obtained from three patients with SCA27B. These patients respectively carried alleles of 292/304 (patient P1), 9/508 (patient P8), and 16/389 (patient P9) repeat units, as measured in blood samples. We found minimal difference in the number of GAA•TTC repeats and EI between paired blood and fibroblasts, providing further evidence for the lack of significant instability of the *FGF14* repeat locus in peripheral tissues (**Supplementary Figures 4-5 and Supplementary Table 2**). Moreover, the sizes of both wild type and expanded alleles remained unchanged across ten passages in fibroblasts from two SCA27B patients (patient P8: 9/518 GAA•TTC repeats; patient P9: 16/409 GAA•TTC repeats).

Reprogramming of fibroblasts into iPSCs resulted in variable increase of the modal peak for most expanded alleles, but not wild type alleles (**Supplementary Figures 4-5 and Supplementary Table 2**). While two iPSC clones from patient P1 exhibited minimal changes in modal alleles compared to fibroblasts, one clone harbored an expansion of 11 triplets of its longer allele, corresponding to an increase in length of 3.6% (**Supplementary Figure 6**). During reprogramming, the longer allele in all four iPSC clones from patient P9 remained relatively stable (modal difference ranging from −19 to +2 triplets; **Supplementary Figure 4**), while it expanded by 7 to 89 triplets in iPSC clones from patient P8 (**Supplementary Figure 5**). Finally, similar to fibroblasts, the sizes of alleles in control (*n*=3; alleles of 8/8, 9/58, and 12/138 repeat) and patient (*n*=3; alleles of 291/315, 9/597, 16/402) iPSCs remained unchanged across serial passages (**Supplementary Figure 7**).

### Analysis of the FGF14 GAA•TTC repeat somatic instability in post-mortem brain tissues

Because transcription through GAA•TTC repeat tracts enhances somatic instability,^27^ the very low expression levels of *FGF14* in blood and fibroblasts^1^ may account for the minimal instability of the repeat in these tissues. In comparison, *FGF14* is widely expressed in the CNS, with the highest expression levels observed in the cerebellum.^7^ Specifically, *FGF14* isoform 1b (*FGF14*-201, transcript 2; ENST00000376131.9; NM_175929.3), within which the GAA•TTC repeat is located in its first intron, has notably higher expression levels in the cerebellum compared to isoform 1a (*FGF14*-202, transcript 1; ENST00000376143.5; NM_004115.4) according to the Genotype-Tissue Expression (GTEx) Project database. We therefore hypothesized that higher degrees of somatic instability may be present in the CNS. To address this question, we conducted a comprehensive analysis of the somatic instability of *FGF14* GAA•TTC repeat tracts in multiple post-mortem CNS regions from six non-ataxic controls and six patients with SCA27B (**Table 1**).

In controls C1-C6 and patients P1-P3, all of whom carried alleles smaller than 400 GAA•TTC triplets, analysis of tissue-wide somatic instability in multiple brain regions revealed that the modal length and EI of both wild type and expanded alleles were remarkably stable across all analyzed regions, except in the cerebellar hemispheres and vermis, which showed a marked somatic expansion bias (**Figures 3** and **4a**; see also Supplementary Results). In control and patient brains, the modal length of wild type and expanded alleles was identical – within two triplets – across all non-cerebellar regions and blood samples, while, for alleles longer than ≈90 repeats, it increased on average by 10 ± 6% in the vermis and 11 ± 6% in the cerebellar hemispheres (**Figure 3a** and **Supplementary Figures 17-25**). Levels of somatic expansions, as measured by the EI, were markedly increased in the cerebellar hemispheres and vermis compared to other CNS regions (**Figure 4a**). Excluding (GAA)_9_ alleles, which did not exhibit somatic expansion, the EI in the cerebellar hemispheres was, on average, 3.23 times greater relative to other CNS regions outside the cerebellum. Furthermore, within the cerebellum itself, the EI in the hemisphere was 1.15 times greater relative to the vermis and 2.42 times greater relative to the dentate nucleus. We found that increasing allele length was associated with higher degrees of instability of the *FGF14* repeat locus, with instability even observed in alleles as short as 15 triplets, indicating that somatic instability is not exclusive to large, pathogenic *FGF14* alleles (**Supplementary Figure 26**). The pattern of somatic expansion in the CNS was closely correlated with the regional expression of *FGF14*, which is highest in the cerebellar hemispheres and vermis (**Figure 4**). This observation supports the hypothesis that higher levels of transcription through GAA•TTC repeat tracts promote their somatic expansion.^27^ Furthermore, we found a positive correlation between the EI and the repeat length in many of the examined tissues, with the steepest regression slopes observed in the vermis (r=0.96 [95% CI: 0.81 to 0.99], slope=0.130, *p*<0.0001) and the cerebellar hemispheres (r=0.93 [95% CI: 0.81 to 0.98], slope=0.133, *p*<0.0001) (**Supplementary Figure 26**). Using a multiple regression model, we found significant relationships between EI and repeat size (estimate = 0.047, SE = 0.005, *p*<0.0001), tissue expression (estimate = 0.133, SE = 0.026, *p*<0.0001), and female sex (estimate = 6.630, SE = 1.744, *p*=0.00022), although the latter association should be interpreted cautiously as it was based on two female cases. In contrast, age at death did not have a significant effect on EI (estimate = 0.040, SE = 0.061, *p*=0.51). These variables together explained approximately 58% of the variance in EI. Overall, our results demonstrate that the *FGF14* repeat locus exhibits a strong somatic expansion bias largely restricted to the cerebellum, that may in part be driven by the high level of *FGF14* expression in this tissue. This observation contrasts with a recent study on SCA1, SCA2, SCA3, and SCA7, which showed that despite high *ATXN1*, *ATXN2*, *ATXN3*, and *ATXN7* gene expression, the cerebellum exhibited the lowest degree of somatic mosaicism.^14^

**Figure 3:**
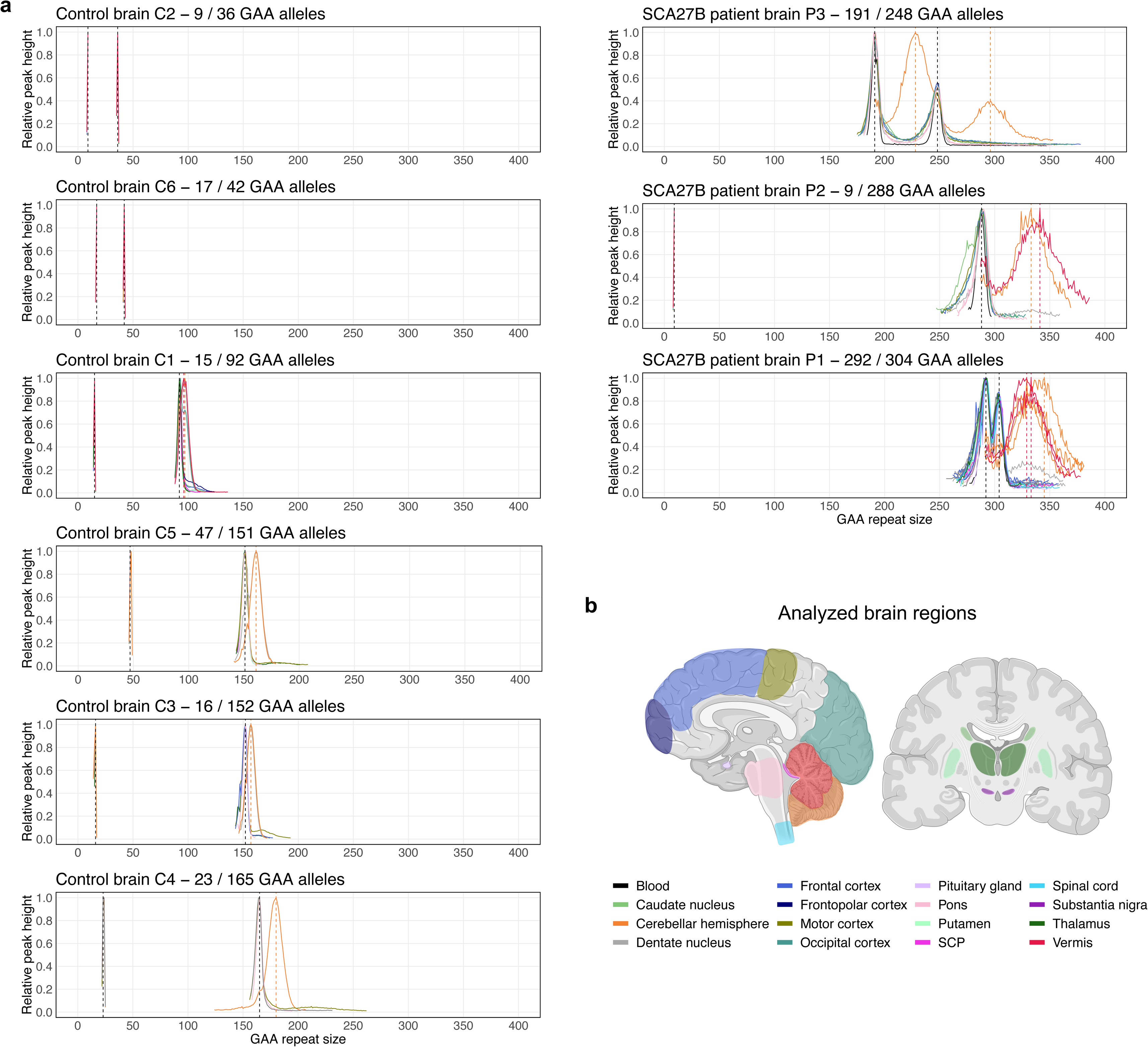
Somatic instability profile of the *FGF14* GAA•TTC repeat in brain regions. (a) Somatic instability profiles of the *FGF14* GAA•TTC repeat in different brain regions, derived from post-mortem samples of six non-ataxic control and three patients with SCA27B. Each plot shows the average instability profile for each of the two alleles within a given brain region, calculated from replicate PCR reactions. For regions where multiple tissue samples were analyzed, results for each sample are shown individually. Profiles were plotted by normalizing individual peak height data to the height of the modal allele within each brain region, except for SCA27B cases P1 and P3, where data were normalized to the height of the shorter modal allele due to the lack of significant size difference with the longer allele. Peaks left of the modal allele above a 10% threshold and those right of the modal allele above a 1% threshold were plotted. Vertical dashed black lines indicate the size of the modal alleles measured in the blood and/or non-cerebellar regions, while vertical dashed red and orange lines indicate the size of the modal alleles measured in the vermis and cerebellar hemispheres, respectively. Individual instability profiles in each region are shown in **Supplementary Figures 8-16**. (b) Schematic representation of the various brain regions analyzed for somatic instability in this study. The same color scheme is used to represent the brain regions and their corresponding instability profiles shown in panel a. Panel created with BioRender.com.

**Figure 4:**
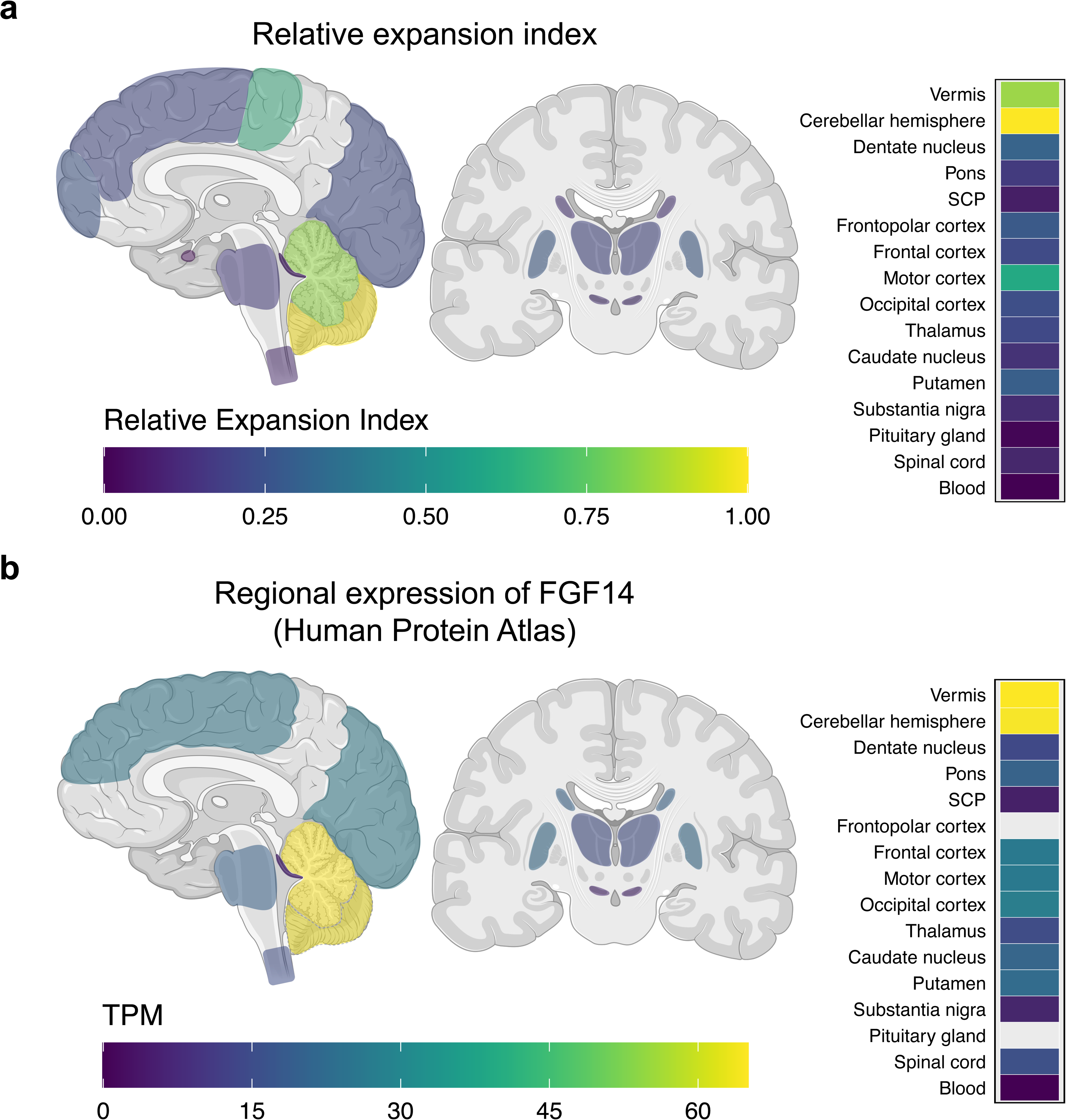
Expansion index of the *FGF14* GAA•TTC repeat in brain regions. Schematic representation of (a) the relative expansion index and (b) the regional expression of *FGF14* (normal tissue expression data in transcripts per million [TPM] from the Human Protein Atlas) across the analyzed brain regions shown in Figure 3. Expansion indices for each brain region were normalized relative to those in the cerebellar hemispheres. The relative expansion indices were calculated by averaging the relative expansion index of each allele, excluding (GAA)_9_ alleles, which did not exhibit somatic expansion. A corresponding heatmap of the results for each region is shown on the right side of both panels. Expression data were not available for the frontopolar cortex and the pituitary gland. Figure created with BioRender.com.

We also studied the somatic instability of the *FGF14* repeats by agarose gel electrophoresis in the post-mortem brains of three patients with SCA27B (P4-P6), each carrying expansions longer than 400 triplets. This analysis confirmed the cerebellar-specific somatic expansion bias of the *FGF14* repeat locus. Compared to other tissues, the expansion length in the cerebellar hemispheres was 87-159 triplets longer in patient P4, 129-185 triplets longer in patient P5, and 70-168 triplets longer in patient P6 (**Figure 5a, Supplementary Figures 53-55**). Furthermore, compared to blood samples, this corresponds to an increase in expansion length in the cerebellar hemispheres of 26% in patient P4 and 41% in patient P6 (**Figure 5b**). Among the three patients, cerebellar vermis was only available for study in patient P4 and exhibited a greater expansion length compared to the cerebellar hemispheres (709 vs. 692 GAA•TTC repeats). Of note, in keeping with the greater levels of *FGF14* expression in cerebellar neurons compared to glia,^26^ the cerebellar hemispheres and vermis showed longer expansion sizes relative to the cerebellar white matter in all three patients. Finally, although the cervical spinal cord exhibited the largest expansion size of all tissues examined from patient P6, this finding was not replicated in the two other patients (P1 and P5) for whom cervical spinal cord was available for study. This discrepancy could be due to differences in the specific regions of the spinal cord sampled for analysis.

**Figure 5:**
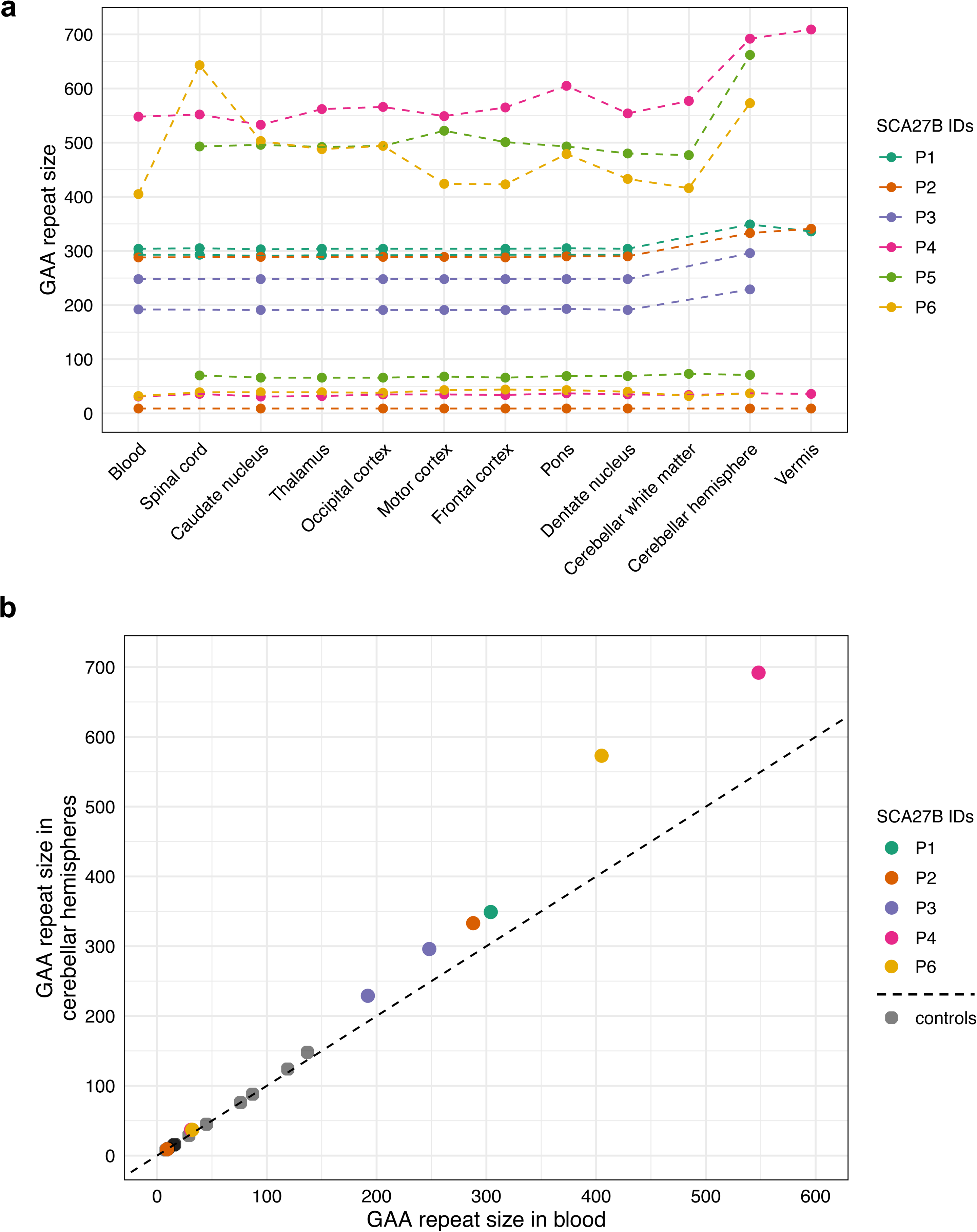
*FGF14* GAA•TTC repeat lengths in brain regions of SCA27B patients. (a) Distribution of *FGF14* GAA•TTC repeat lengths, expressed in triplet repeat counts, across post-mortem brain regions and blood samples in the six SCA27B patients included in this study. For brain regions where multiple tissue samples were analyzed, only the results of the sample with the largest allele size are shown. Observations for each of the two alleles from the same patient are connected by a dashed line. (b) Comparison of *FGF14* GAA•TTC repeat lengths in blood samples (*x*-axis) versus cerebellar hemispheres (*y*-axis) for five SCA27B patients and eight controls with available tissue samples. The dashed black line represents the identity line. In panels a and b, a single modal allele size is shown for patient P1 due to the complete blending of the instability profiles for the short and long alleles in the cerebellar hemispheres and vermis (**Supplementary Figure 33**), preventing the identification of the two distinct modal peaks in these tissues.

Comparison of modal lengths in blood samples and cerebellar hemispheres from five patients with SCA27B (P1, P2, P3, P4, P6) and eight control individuals (C7-C14) revealed that sizes were largely stable in both tissues for alleles shorter than ≈90 repeat units. However, for longer alleles, the modal length in the cerebellar hemispheres increased with allele size (r=0.99 [95% CI: 0.96 to 1.00], slope=1.365, *p*<0.0001) (**Figure 5b**). Remarkably, this threshold of ≈90 repeat units for instability in the cerebellar hemispheres is highly similar to the threshold of 75-100 repeat units for instability during intergenerational transmission.^4, 8^ Despite the similar instability thresholds, different mechanisms are likely responsible for instability in the cerebellum and the germline, given the comparatively lower *FGF14* expression in testes and ovaries and the repeat contraction observed with paternal transmission.^4, 8^

### Methylation profiling of FGF14 in post-mortem cerebellum

Preliminary investigations of patient-derived post-mortem cerebellum and iPSC-derived neurons have suggested that the intronic GAA•TTC repeat expansion likely leads to loss of function of isoform 1b (transcript 2) by interfering with *FGF14* transcription.^1^ Since the *FGF14* expansion shares the same repeat motif as the Friedreich ataxia’s *FXN* expansion (GAA•TTC), which is known to cause hypermethylation of the DNA upstream of the expanded repeat locus (differentially methylated region),^28^ we investigated whether altered methylation was present in post-mortem cerebellum from four patients with SCA27B. Using programmable targeted ONT long-read sequencing,^29^ we found no evidence of significant difference in 5mC methylation frequencies of *FGF14* (median frequency [IQR] in patients vs controls, 69.75 [68.65 to 69.80] vs 68.95 [67.43 to 69.80]; Mann-Whitney U test, *p*=0.63) and its two putative promoters (first promoter: 2.00 [1.35 to 2.27] vs 2.25 [1.40 to 3.25]; Mann-Whitney U test, *p*=0.54; and second promoter: 0.85 [0.57 to 1.35] vs 0.80 [0.40 to 0.90]; Mann-Whitney U test, *p*=0.63) in controls compared to patients (**Figure 6**). The region surrounding the repeat expansion (10 kb window centred on the repeat locus) showed slightly higher CpG methylation compared to the region surrounding wild type alleles, although the difference was not statistically significant (median frequency [IQR] in expanded alleles vs wild type alleles, 82.37 [79.96 to 84.76] vs 78.78 [76.66 to 80.55]; Mann-Whitney U test, *p*=0.11). These results do not exclude the possibility that the expansion is associated with hypermethylation of *FGF14*, which will require further study in a larger number of cases.

**Figure 6:**
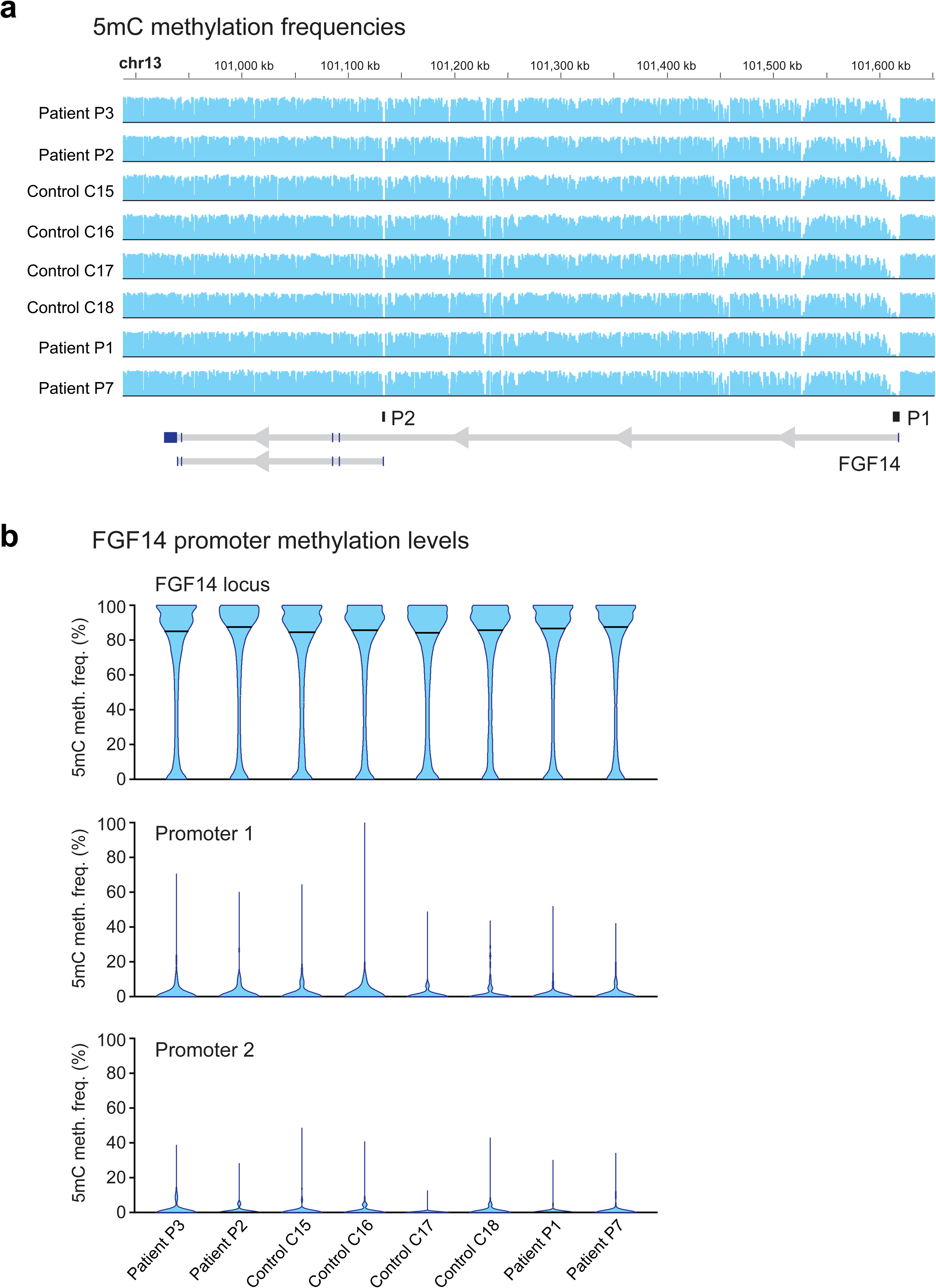
Methylation profiling of the *FGF14* gene in post-mortem cerebellum. Methylation analysis of the *FGF14* gene in post-mortem cerebellar hemispheres from four controls (C7-C10) and four patients with SCA27B (P1, P2, P3, P7) using programmable targeted long-read sequencing with Oxford Nanopore Technologies. (a) Methylation profiles, expressed as 5’-methylcytosine (5mC) methylation frequencies at all CpG sites within the *FGF14* locus (T2T-CHM13, chr13:100923763-101619864), in the four control and four patient post-mortem cerebellar hemispheres. A diagram of the *FGF14* gene with its two putative promoters (P1: promoter 1 and P2: promoter 2) is shown at the bottom of the panel. (b) Plots shows the distribution of 5mC methylation frequencies at all CpG sites within the *FGF14* locus (top plot), the first promoter (middle plot), and second promoter (bottom plot). The horizontal black bars show the median 5mC methylation frequency.

## Discussion

This study comprehensively assessed the somatic instability of the *FGF14* GAA•TTC repeat in peripheral tissues and post-mortem brains of patients with SCA27B and uncovered a marked tendency for the repeat to somatically expand in the cerebellar vermis and hemispheres. Our findings provide new insight into the biology of SCA27B, potentially explaining the selective vulnerability of the cerebellum – specifically the gray matter – and the relative resilience of other regions in this disease.^1, 3^ Although somatic expansion events were detected in all tissues analyzed, the expansion bias was considerably higher in the cerebellar hemispheres and vermis, which are the primary sites of neuronal loss in SCA27B.

Although brain mosaicism has only been studied in a few spinocerebellar ataxias, previous studies of SCA1, SCA2, SCA3, SCA7, and DRPLA have consistently shown that their causative coding CAG•CTG repeat expansions exhibit smaller sizes and lower degrees of mosaicism in the cerebellum compared to other brain regions.^13–17^ Studies of Huntington disease, myotonic dystrophy type 1, and spinal and bulbar muscular atrophy, all caused by CAG•CTG repeat expansions, have also shown that the smallest expansions and lowest somatic instability occur in the cerebellum.^13, 30–32^ In comparison, a recent study found that the pathogenic *DAB1* intronic ATTTC repeat insertion causing SCA37 exhibits longer sizes and greater somatic expansion in the cerebellum compared to blood samples and fibroblasts of two patients,^18^ suggesting that the instability pattern of CAG•CTG repeat expansions in the cerebellum may differ from that observed with other types of repeats. The factors underlying the distinct behavior of the *FGF14* GAA•TTC repeat expansion in the cerebellum compared to CAG•CTG repeat expansions remain unknown, although they may involve *cis*-acting elements, differential effects of mismatch repair pathways on CAG•CTG repeats compared to GAA•TTC repeats, or an increased ability of the long and uninterrupted *FGF14* GAA•TTC repeat to form alternative DNA structures that can become substrates for DNA repair machineries.^33^

In Friedreich ataxia, although the expanded *FXN* GAA•TTC repeat is longer in the cerebellum than in other CNS regions, large contractions occur significantly more frequently than large expansions in the cerebellum.^20–23^ Progressive expansion of the *FXN* repeat over time has been described in human cells, cerebellum, and other terminally differentiated tissues in both human and mouse models.^20–23, 34, 35^ DNA repair has been established as a major factor modulating GAA•TTC repeat stability.^9, 33^ Studies in a nondividing quiescent *S. cerevisiae* model have shown that large contractions of the GAA•TTC repeat expansion may be driven by non-homologous end joining repair of DNA breaks of the repeat mediated by mismatch repair complexes.^36^ Furthermore, activity of the mismatch repair complex MutSβ has been shown to promote *FXN* GAA•TTC expansion in Friedreich ataxia patient-derived cells.^37, 38^ Similarly, DNA repair mechanisms may also be involved in somatic expansion of the *FGF14* GAA•TTC repeat, especially considering the high expression levels of mismatch repair genes in the cerebellum compared to other brain regions.^14^ Moreover, 5’ GAA-strand and 5’ TTC-strand nicks (single-strand breaks) have recently been shown to induce expansion of GAA•TTC repeats in non-dividing yeast cells,^39^ raising the hypothesis that such phenomenon may contribute to somatic instability in post-mitotic neurons, which are known to accumulate single-strand breaks over time.^40^

Our findings suggest that tissue-specific transcriptional activity through the GAA•TTC repeat promotes its instability, which align with a previous study on GAA•TTC repeat stability in human cell lines.^27^ The GAA•TTC repeat is located within intron 1 of *FGF14* isoform 1b, the isoform predominantly expressed in the CNS, particularly in the cerebellum.^7^ Analysis of tissue-wide somatic instability revealed that the pattern of somatic expansion in the CNS closely correlates with the regional expression of *FGF14*. The cerebellar vermis and hemispheres, which have the highest and second highest levels of *FGF14* expression in the CNS, respectively, exhibited the greatest somatic expansion across all tissues analyzed. Moreover, the association between *FGF14* repeat instability and expression levels is further corroborated by our results showing longer expansions in the cerebellar hemispheres and vermis compared to the cerebellar white matter, in keeping with the substantially greater expression of *FGF14* in cerebellar neurons relative to glia.^26^ Alternative DNA structures called DNA triplexes are formed in expanded GAA•TTC repeats in Friedreich ataxia patient-derived cells,^41^ and transcription through these repeats has been shown to induce the formation of triplexes, which may fuel repeat instability.^9, 42^ This observation contrasts with a recent study of the somatic instability of CAG•CTG repeats, which are prone to form DNA hairpins,^43^ but not DNA triplexes, in the polyglutamine spinocerebellar ataxias SCA1, SCA2, SCA3, and SCA7. These pathogenic CAG•CTG repeats showed lower levels of expansion in the cerebellum despite high expression levels of the *ATXN1*, *ATXN2*, *ATXN3*, and *ATXN7* genes.^14^ The absence of significant somatic instability of the CAG•CTG repeat in the cerebellum, which is one of the most pathologically affected brain regions in these spinocerebellar ataxias,^44^ further supports the possibility that different alternative DNA structures and various repair pathways may differentially impact the stability of CAG•CTG repeats compared to GAA•TTC repeats in the cerebellum.

Although we observed no change in repeat size and EI in blood samples over time, somatic expansions of the *FGF14* mutant alleles in the cerebellar cortical neurons are likely to progress with age, with the rate of expansion increasing with the number of repeats. Indeed, we found that increasing allele length was associated with higher degrees of instability of the *FGF14* GAA•TTC repeat locus. While our study did not look at mosaicism in fetal brains, somatic instability is likely to progress throughout life rather than arise during CNS development (as previously shown in Friedreich ataxia^20^ and Huntington disease,^45^ for example) as regions with the same embryological origin, such as the cerebellar cortex, dentate nuclei, and pons, all deriving from the metencephalon,^46^ present dramatically different instability patterns. This hypothesis would also be consistent with the late age at onset of SCA27B, suggesting that the disease may only manifest when a critical proportion of cerebellar Purkinje cells accumulate expansions above a cell-specific pathogenic threshold, which is likely to be higher than the currently used ‘blood-based threshold’.^47^ It also raises the possibility that loss of function of *FGF14* isoform 1b may only occur when repeat sizes extend beyond this specific threshold in cerebellar Purkinje cells. This hypothesis also provides a potential explanation for the incomplete penetrance of certain *FGF14* expansions, suggesting that asymptomatic carriers may accumulate somatic expansions in the cerebellum at lower rates compared to symptomatic individuals, preventing them from accumulating a sufficiently large burden of somatic expansions to manifest the disease. Furthermore, variable somatic expansion, which may be influenced by variations in DNA repair genes, may underlie the significant phenotypic variability among patients carrying expansions of similar sizes.

Our study has several strengths, including the inclusion of a large number of post-mortem controls and SCA27B cases with a variety of allele sizes, the availability of serial blood samples for many patients over long periods of time, and the ability to accurately size and calculate expansion indices in samples carrying expansions shorter than ≈400-450 triplets. However, our results need to be interpreted in light of some limitations. The inability of capillary electrophoresis to resolve low-abundance alleles and alleles longer than ≈400-450 triplets prevented a comprehensive analysis of the full spectrum of allele length variability in the cerebellum. Analysis of bulk PCR products by capillary electrophoresis will preferentially detect shorter GAA•TTC tracts and cannot capture the full extent of GAA•TTC repeat expansions in any given tissue. Furthermore, our analyses did not permit the detection of rare, very large expansion events that may be occurring in Purkinje cells, which are primarily affected in SCA27B. Purkinje cells make up only a small fraction of the total cerebellar cell population and, as such, do not contribute significantly to the instability readout measured in bulk tissue. Additional techniques, such as single-cell sequencing, will be necessary to dissect cell-specific somatic instability in SCA27B. Moreover, in post-mortem tissues from patients with neurodegenerative ataxia, it is unknown whether the genotype of degenerated cerebellar neurons is similar to that of the remaining cells. However, it is possible that neurons carrying longer expansions degenerate faster, leading to an underrepresentation of longer allele sizes in the instability readout.

In conclusion, our analysis of the *FGF14* GAA•TTC repeat somatic instability in post-mortem brain samples from individuals with SCA27B revealed a marked expansion bias in the cerebellar hemispheres and vermis. Our study offers new insights into the biology of SCA27B by providing a potential explanation for the significant phenotypic variability and the incomplete penetrance of certain expansions. It also opens up important new lines of inquiry for understanding the mechanisms driving somatic instability of tandem repeats, the cell-specific patterns of instability, and their relationship to neurodegeneration.

## Acknowledgments

We thank the patients and their families for participating in this study. Part of this work was carried out in the DNA and cell bank of the Paris Brain Institute - ICM. We gratefully acknowledge Ludmila Jornea and Philippe Martin Hardy for their help as well as Sabrina Leclère-Turbant from Neuro-CEB. We are indebted to the Biobanc-Hospital Clinic-FRCB-IDIBAPS for samples and data procurement.

## Author contributions

Design or conceptualization of the study: DP, JLM, MCD, AB, SZ, AD, BB.

Acquisition of data: DP, JLM, SB, MCD, MJD, CSD, DG, GS, CA, JMH, BJG, VC, PUL, MS, CY, HL, JSN, HH, IWD, MN, AB, LMP, DS, SZ, AD, BB.

Analysis or interpretation of the data: DP, JLM, SB, MCD, MJD, CSD, DG, GS, CA, JMH, BJG, VC, PUL, MS, CY, HL, SKN, GMR, ZJ, KP, SA, AN, KU, MR, CB, GR, MAS, JSN, HH, IWD, MN, AB, LMP, DS, SZ, AD, BB.

Drafting or revising the manuscript for intellectual content: DP, JLM, SB, MCD, MJD, CSD, DG, GS, CA, JMH, BJG, VC, PUL, MS, CY, HL, SKN, GMR, ZJ, KP, SA, AN, KU, MR, CB, GR, MAS, JSN, HH, IWD, MN, AB, LMP, DS, SZ, AD, BB.

All authors read and approved the final version of the manuscript.

