## Supporting Information for "Somatic instability of the *FGF14*-SCA27B GAA•TTC repeat reveals a marked expansion bias in the cerebellum"

### Supplementary Methods

*Human sample collection*

DNA samples from 24 individuals attending the ataxia clinic at the Montreal Neurological Hospital, Montreal, and 45 individuals attending the neurogenetic clinic at the Hôpital Pitié-Salpêtrière, Paris, who had multiple blood samples collected during follow up ('longitudinal samples') were analyzed to determine the stability of the *FGF14* GAA•TTC repeat over time. We also analyzed the stability profile of the *FGF14* repeat in 31 additional individuals (*n* = 29 from Montreal, *n* = 2 from Paris), including two pairs of monozygotic twins with SCA27B. Genomic DNA was isolated from peripheral blood using the Puregene Blood kit (catalog no. 158026, Qiagen) or the Tecan Freedom EVO-HSM workstation with ReliaPrep Large Volume HT gDNA Isolation System kit (catalog no. A2751, Promega) as per the manufacturer's instructions.

Fibroblast lines (*n* = 3 patients with SCA27B) and induced pluripotent stem cells (iPSCs; *n* = 3 controls and *n* = 3 patients with SCA27B) were serially passaged 10 times and harvested after each passage to assess the stability of the *FGF14* repeat locus. We also compared the repeat length and instability profile of different iPSC clones generated from fibroblasts of three patients with SCA27B who respectively carried *FGF14* alleles of 292 / 304 (patient P1), 9 / 508 (patient P8), and 16 / 389 (patient P9) repeat units.

Human post-mortem brain tissue from six non-ataxic control individuals and six persons with SCA27B were obtained from the Douglas-Bell Canada Brain Bank, Montreal, QC, Canada (controls C1-C2, patient P1), the Neurological Tissue Brain Bank, Biobanc-Hospital Clínic- Fundació de Recerca Clínic Barcelona-Institut d'Investigacions Biomediques August Pi i Sunyer , Barcelona, Spain (patients P2-P3), and the NeuroCEB Brain Bank of the Hôpital Pitié-Salpêtrière, Paris, France (controls C3-C6, patients P4-P6) for the study of the somatic stability of the *FGF14* GAA•TTC repeat tract. None of the six controls had evidence of significant cerebellar pathology on neuropathological examination. When possible and based on tissue availability, more than one sample from any given central nervous system (CNS) region was obtained and analyzed. Blood samples were available for five of the six SCA27B patients (not available for patient P5), but not for the controls. DNA was extracted from fresh frozen brain tissue using the MagAttract HMW DNA kit (catalog no. 67563, Qiagen) or the Maxwell RSC Blood DNA Kit (catalog no. AS1400, Promega) as per manufacturer's instructions. Fresh frozen tissue of the cerebellar hemispheres and blood samples from an additional eight control individuals (controls C7-C14) were obtained through the Queen Square Brain Bank for Neurological Disorders, London. DNA was extracted from frozen brain tissue using the QIAamp DNA Mini Kit (catalog no. 56304, Qiagen) as per manufacturer's instructions.

In addition to the post-mortem CNS tissues used for the somatic stability study described above, we also obtained fresh frozen tissue of the cerebellar hemispheres from four additional controls, respectively carrying *FGF14* alleles of

8 / 17 (control C15), 9 / 96 (control C16), 16 / 51 (control C17), and 9 / 44 (control C18) repeat units, and one additional SCA27B patient (patient P7), carrying alleles of 9 / 432 repeat units, for methylation profiling analysis of *FGF14*. DNA was extracted from fresh frozen brain tissue using the MagAttract HMW DNA kit as per manufacturer's instructions (catalog no. 67563, Qiagen).

*Repeat length genotyping*

We genotyped the *FGF14* repeat locus by capillary electrophoresis or agarose gel electrophoresis of long-range PCR amplification products, as described previously.[1] Alleles longer than ≈400-450 triplets could not be sized by capillary electrophoresis as they fell beyond the limit of detection of this technique,[1] and were genotyped by agarose gel electrophoresis. For capillary electrophoresis, amplification products were analyzed on an ABI 3730*xl* DNA Analyzer (Applied Biosystems) with a 50-cm POP-7 capillary using the GeneScan 1200 Liz Dye Size Standard (catalog no. 4379950, Applied Biosystems). Results were analyzed with the GeneMapper software v6.0 (Applied Biosystems) using the built-in microsatellite default settings or the Peak Scanner software v1.0 (Applied Biosystems). For agarose gel electrophoresis, amplification products were resolved on a 1% agarose E-Gel 48 (catalog no. G800801, Invitrogen) and migrated for 25 min using the E-base system (catalog no. EB-M03, Invitrogen). Agarose gels were imaged with the ChemiDoc Touch Imaging System (Bio-Rad) and analyzed with Image Lab v6.1.0 (Bio-Rad). The GAA•TTC purity of large alleles was confirmed by bidirectional repeat-primed PCR.[1] Genotyping of each individual CNS region was performed in triplicate for samples from Montreal and Barcelona and in duplicate for samples from Paris, and technical variation across replicate PCR was assessed.

*Fibroblast cell culture*

Fibroblasts obtained from skin biopsy of three patients with SCA27B, respectively carrying *FGF14* alleles of 292 / 304, 9 / 508, 16 / 389 repeat units, as measured in whole-blood-extracted DNA, were cultured in DMEM, high glucose with 10% (v/v) fetal bovine serum, 100U/ml penicillin-streptomycin, 2.5µg/ml amphotericin B and 100µg/ml L-glutamine. Cells were grown at 37°C, 5% $CO_2$ in 95% humidified air.

Fibroblasts were serially passaged 10 times and harvested after each passage to genotype the *FGF14* repeat locus by long-range PCR.[1] DNA was extracted using the DNeasy Blood and Tissue kit (catalog no. 69504, Qiagen).

*Generation of induced pluripotent stem cells*

Induced pluripotent stem cells (iPSCs) were generated in Miami, Florida, using the CytoTtune-iPS 2.0 Sendai Virus Reprogramming kit (catalog no. A16518, Thermo Fisher Scientific) using the feeder-free reprogramming method for fibroblasts. These fibroblast lines originated from the same three patients with SCA27B (see *Fibroblast cell culture* above). Candidate iPSC colonies were isolated based on general morphology for further characterization. iPSCs underwent immunocytochemistry (ICC) for NANOG, SSEA3, and OCT4 to demonstrate their stem-cell like and pluripotent state. iPSC clones were sent to Cell Line Genetics for karyotype analysis to demonstrate no chromosomal anomalies that may affect downstream experiments and cell line health. iPSC colonies were cultured in mTeSR Plus media, with cell culture plates being coated with Geltrex. iPSCs were serially passaged 10 times and harvested after each passage to genotype the *FGF14* repeat locus by long-range PCR.[1] DNA was extracted using the DNeasy Blood and Tissue kit (catalog no. 69504, Qiagen).

iPSCs were also obtained in Dallas, Texas, using Sendai virus transduction (CytoTune-iPS 2.0 Sendai Reprogramming Kit, Thermo Fisher Scientific) and characterized as previously described.[2] For instability assay, iPSC were cultured on Matrigel (Corning) in mTeSR™1 (StemCell Technologies) as recommended by the manufacturer. Confluent cells were enzymatically dissociated and passaged to the fresh well of a six-well plate every ~5 days. Each passage was initiated with ~ 20% of dissociated cells from the previous passage and the remaining cells were pelleted for genomic DNA isolation. The iPSCs were passaged 10 times in the span of ~50 days. Subsequently, genomic DNA was isolated from each pellet using the DNeasy kit. Amplification of GAA•TTC repeats in the *FGF14* gene was performed using primers: FGF14_F1: 5'-AGCAATCGTCAGTCAGTGTAAGC and FGF14_R1: 5'-CAGTTCCTGCCCACATAGAGC.[3] Reactions were performed in 50 µL volume with 50 ng of genomic DNA using FailSafe PCR System with mix D (Epicentre). To reduce heteroduplex formation, PCR products containing varying lengths of GAA•TTC repeats were cooled down from 94°C to 4°C at a rate of 1°C/min after PCR amplification reactions before agarose gel electrophoresis. The length of the *FGF14* GAA•TTC repeats was determined using the base pair size called by Image Lab 6.1, with the total number of GAA•TTC repeats calculated by subtracting the length of the sequences flanking the GAA•TTC repeats (165 bp) from the total number of base pairs of the PCR product and dividing by three [Number of GAA•TTC repeats = (length of base pairs of a PCR product – 165)/3].

*Analysis of somatic instability*

Somatic stability was studied by capillary electrophoresis or by agarose gel electrophoresis for samples carrying alleles >400-450 triplets. For samples analyzed by capillary electrophoresis, the GeneMapper software v6.0 (Applied Biosystems) or Peak Scanner software v1.0 (Applied Biosystems) was used to establish the 'modal peak' of each of the two alleles in each sample. The modal peak was defined as the peak with the highest height, measured in relative fluorescence intensity units, within each of the two clusters of peaks that correspond to each allele. Peaks to the right of the modal allele represent somatically expanded GAA•TTC repeats, while peaks to the left may include both contracted GAA•TTC repeats and PCR stutter products. Expansion, contraction, and instability indices were calculated using peak height data extracted from GeneMapper or Peak Scanner (**Figure 1** and **Supplementary Figure 1**), as described previously.[4] Each chromatogram was inspected and individual peaks were manually called if not automatically detected by the software. Expansion indices were calculated by only taking into account peaks to the right of the 'modal allele' and using a 1% relative peak threshold (**Figure 1**). Contraction indices were calculated by only taking into account peaks to the left of the 'modal allele' and using a 10% relative peak threshold, while instability indices were calculated by taking into account peaks on both sides of the 'modal allele' and using a 10% relative peak threshold (**Supplementary Figure 1**). To prevent overestimation of contraction and instability indices from the excessive inclusion of PCR stutter products, a higher threshold was applied for their calculation. PCR stutter products increased incrementally and spanned the entire baseline in presence of longer alleles. Each index was obtained by first dividing the height of each individual peak by the sum of the heights of all peaks above the set threshold. The resulting normalized peak heights were then multiplied by the position of the peak and these normalized peak values were summed to generate the index. Analysis of blood and CNS tissues revealed that the modal length of both wild type and expanded alleles was remarkably stable across all regions examined, except in the cerebellar hemispheres and vermis where it showed a tendency to expand. We therefore considered the modal allele length as measured in the blood and most CNS regions as the constitutional allele length, which we set as the main allele for calculation of the indices in tissues with an expansion shift of the modal repeat,[5] tolerating a discrepancy of one triplet due to the relative imprecision of our genotyping technique.[1] The calculation of certain indices for alleles of SCA27B patients P1 and P3 was not possible because of overlapping peak clusters resulting from insufficient size difference between alleles. Furthermore, contraction and instability indices of longer alleles were not calculated in the vermis and cerebellar hemispheres as the marked somatic expansion resulted in insufficient height difference between the main allele and the adjacent peaks to its left.

*Methylation profiling of FGF14 in control and patient cerebellum*

We performed methylation analysis of *FGF14* in the post-mortem cerebellar hemispheres of four controls (C7-C10) and four SCA27B patients (P1, P2, P3, P7) using programmable targeted long-read sequencing[6] with Oxford Nanopore Technologies (ONT). High molecular weight (HMW) DNA samples extracted from post-mortem cerebellar hemispheres were transferred to the Garvan Institute's Sequencing Platform (Sydney), for programmable targeted ONT sequencing of the *FGF14* locus. Prior to ONT library preparations, the DNA was sheared to ~20 kb fragment size using a Diagenode MegaRuptor3 and visualised, post-shearing, on an Agilent FemtoPulse. Sequencing libraries were prepared from ~3-5 ug of HMW DNA, using native library prep kit and barcoding kits SQK-LSK114 and NBD114.24. Barcoded libraries were pooled in groups of three, then each pool was loaded onto an ONT R10.4.1 flow cell and sequenced on a PromethION device with live target selection/rejection executed by the ReadFish software package.[7] Detailed descriptions of gene targets, software and hardware configurations used for ReadFish experiments are outlined in a previous publication.[6] Samples were run for a maximum duration of 72 hours, with nuclease flushes and library reloading performed at approximately 24- and 48-hour timepoints for targeted sequencing runs, to maximize sequencing yield.

Raw ONT sequencing data was converted to BLOW5 format[8] using slow5tools v0.8.0.[9] Basecalling and 5'-methylcytosine (5mC) detection was performed using the Buttery-eel v0.4.2[10] SLOW5 wrapper for Guppy v6.5.7, with the model *dna_r10.4.1_e8.2_400bps_5khz_modbases_5mc_cg_sup_prom.cfg*. The resulting reads were aligned to the T2T-CHM13v2.0 reference genome using minimap2 v2.22[11] and converted to bedmethyl format expressing CpG methylation frequencies using mod_kit 0.2.3. Bedmethyl files were converted to bigwig format for visualization in the IGV browser, and CpG methylation frequency distributions were computed for each allele (haplotype-specific) and for all unphased reads for each sample within the overall *FGF14* locus (chr13:100923763-101619864), as well as its two putative promoter regions (P1 chr13:101612311-101618760 and P2 chr13:101131991-101134495). Distributions were plotted in GraphPad Prism v10.

*Statistics*

We assessed differences between groups with the non-parametric Mann-Whitney U test for continuous variables. We used the Sign test to evaluate differences between matched groups. Correlations were calculated using the Pearson's correlation coefficient. Somatic instability was analyzed by multiple regression as a function of the allele length, levels of *FGF14* expression, age at death, and sex. To assess the change in repeat size or instability index over time, we used a linear mixed-effects model fitted using the restricted maximum likelihood method (R packages: lme4 and lmerTest).[12] We analyzed the data in R (version 4.3). *P* value of <0.05 was considered significant. All analyses were two-sided.

### Supplementary Results

*Longitudinal analysis of FGF14 GAA•TTC repeat somatic instability and contraction indices in peripheral blood samples*

We performed a longitudinal analysis of the *FGF14* GAA•TTC repeat size in DNA extracted from peripheral blood to determine whether the length of the repeat tracts changes over time in serial blood samples from 69 individuals, 57 of whom had two serial samples, seven of whom had three serial samples, four of whom had four serial samples, and one of whom had five serial samples. The median interval between the first and last blood sample was 8.9 years (interquartile range [IQR]: 2.9 to 13.9). We observed no significant change of the modal *FGF14* repeat size between the first and last blood samples (**Figure 2a**; median difference [size of last sample – size of first sample], 0 triplet; IQR: 0 to 0; Sign test, $p$=0.22). Of the 120 alleles examined, 12 (10%) expanded by 1 triplet over time and 20 (17%) contracted by 1 to 3 triplets. Out of a total of 37 alleles longer than 200 triplets, the modal size remained stable in 15 alleles (40%), increased in 5 alleles (14%), and decreased in 17 alleles (46%). Similarly, we found no significant change over time in the size of the maximum GAA•TTC repeat using the 1% relative peak height threshold (median difference, 0 triplet [IQR: -1 to 0]; Sign test, $p$=0.43), including in alleles longer than 200 triplets (median difference, 0 triplet [IQR: -1 to 1]; Sign test, $p$=0.72). More than half of maximum alleles remained stable over time (63/120, 52%), while 32 (27%) contracted by 1 to 34 triplets (median difference, -1 triplet [IQR: -2 to -1]) and 25 (21%) expanded by 1 to 18 triplets (median difference, 1 triplet [IQR: 1 to 3]). In maximum alleles longer than 200 triplets, six remained stable over time (6/37, 16%), 17 (46%) contracted (median difference, -2 triplets [IQR: -2 to -1]), and 14 (38%) expanded (median difference, 2 triplets [IQR: 1 to 3]). We found no significant effect of elapsed time between blood samples on the change in modal and maximal repeat size as assessed by a linear mixed-effects model (estimate = -0.441, standard error = 0.678, $p$=0.52 and estimate = -0.374, standard error = 0.688, $p$=0.59, respectively).

We next sought to determine if the expansion index (EI) changed over time in serial blood samples from the same 69 individuals (**Figure 2b-c**). As for repeat sizes, EI did not significantly change over time in repeated blood samples (median of differences [EI of last sample – EI of first sample], 0 [IQR: -0.02 to 0.01]; Sign test, $p$=0.22), nor in the subset of alleles longer than 200 triplets (median of differences, 0 [IQR: -0.28 to 0.43]), Sign test, $p$=1). The EI remained unchanged in 25 of 120 alleles (21%), decreased in 54 alleles (45%) (median of difference, -0.02 [IQR: -0.24 to -0.01]), and increased in 41 alleles (34%) (median of difference, 0.08 [IQR: 0.01 to 0.48]) (**Figure 2c**). In the 37 alleles longer than 200, the EI remained stable in one (3%), decreased in 18 (49%) (median of difference, -0.30 [IQR: -0.37 to -0.18]), and increased in 18 (49%) (median of difference, 0.45 [IQR: 0.28 to 0.58]) (**Figure 2c**).

Similar to previous studies of peripheral blood samples from patients with Friedreich ataxia that have shown a contraction bias of long GAA•TTC repeat expansions in *FXN*,[13, 14] we observed that the somatic instability of *FGF14* alleles was generally skewed toward contractions (indicated by a negative instability index). This estimate may, however, be biased by the inclusion of PCR stutter products in the calculation of the index due to the impossibility of distinguishing those from genuine contraction events. Somatic instability increased proportionately with increasing allele length (Pearson's r = -0.59 [95% confidence interval (CI): -0.68 to -0.48], slope = -0.004, $p<0.0001$; **Supplementary Figure 2a**). Of the 173 alleles analyzed, 160 (92%) showed a contraction bias, five (3%) showed an expansion bias, and eight (5%) showed no somatic instability. In alleles longer than 200 triplets, only three of the 55 alleles (5%) showed an expansion bias, while the rest (52/55, 95%) showed a contraction bias. We next examined whether the instability index changed over time in serial blood samples. We found that the instability index did not significantly change over time in repeated blood samples (**Supplementary Figure 2b**, median of differences [instability index of last sample – instability index of first sample], 0.01 [IQR: -0.22 to 0.27]; Sign test, $p$=0.46), including in the subset of alleles longer than 200 triplets (median of differences, -0.32 [IQR: -0.77 to 0.79]), Sign test, $p$=0.74). In the 37 alleles longer than 200, the instability index remained stable in one (3%), decreased in 20 (54%) (median of difference, -0.77 [IQR: -1.33 to -0.38]), and increased in 16 (43%) (median of difference, 1.00 [IQR: 0.63 to 1.79]) (**Supplementary Figure 2c**). As observed with the expansion index, we found that allele size (estimate = -5.675e-03, SE = 5.455e-04, $p<0.0001$), but not elapsed time (estimate = 8.753e-03, SE = 8.806e-03, $p$=0.32), the interaction between allele size and elapsed time (estimate = -5.977e-05, SE = 6.171e-05, $p$=0.33), and sex (estimate = 1.859e-02, SE = 1.164e-01, $p$=0.87) had a significant effect on the instability index in a linear mixed-effects model.

We next sought to determine the relationship between somatic contraction, as measured by the contraction index, and the size of the *FGF14* GAA•TTC repeat tract. We found that somatic contractions increased linearly with repeat size (Pearson's r = -0.88 [95% CI: -0.91 to -0.84], slope = -0.013, $p<0.0001$; **Supplementary Figure 3a**). We also found that the contraction index did not significantly change over time in repeated blood samples from 69 individuals (**Supplementary Figure 3b**, median of differences [contraction index of last sample – contraction index of first sample], 0.01 [IQR: -0.12 to 0.22]; Sign test, $p$=0.26), including in the subset of alleles longer than 200 triplets (median of differences, -0.05 [IQR: -0.72 to 0.55]), Sign test, $p$=1). In the 37 alleles longer than 200, the contraction index remained stable in one (3%), decreased (indicating an increase of contraction events) in 19 (51%) (median of difference, -0.72 [IQR: -2.06 to -0.29]), and increased in 17 (46%) (median of difference, 0.63 [IQR: 0.08 to 1.37]) (**Supplementary Figure 3c**). Lastly, allele size (estimate = -1.401-02, SE = 6.730e-04, $p<0.0001$) and the interaction between allele size and elapsed time (estimate = -2.117e-04, SE = 7.350e-05, $p$=0.0044), but not elapsed time (estimate = 1.793e-02, SE = 1.053e-02, $p$=0.090), and sex (estimate = 4.252e-02, SE = 1.499e-01, $p$=0.78) had a significant effect on the contraction index in a linear mixed-effects model.

*Analysis of somatic FGF14 GAA•TTC repeat instability in post-mortem brain regions of patients with SCA27B*

In addition the modal shift and high expansion index, the pronounced somatic instability in the cerebellum of patients with SCA27B was further evidenced by the complete rightward shift and "flattened" appearance of the instability profiles on capillary electrophoresis, as opposed to the typical bell shape seen in other tissues, indicative of significant somatic expansion (**Supplementary Figures 33-35**). This resulted in the complete blending of the profiles for the short and long alleles of patient P1, making it impossible to distinguish the contribution of each allele to the overall profile. This likely led to an underestimation of the modal shift and expansion index in the cerebellum, since these were calculated relative to the longer allele. The flattened and widened appearance of the instability profile of expanded *FGF14* alleles in the cerebellum further suggests the absence of a single predominant allele subtype, but instead indicates the presence of multiple populations of alleles in similar proportions. This observation also implies a notably broader spread and a more homogeneous distribution of allele sizes within the cerebellum. However, the inability of capillary electrophoresis to resolve low-abundance alleles and alleles longer than ≈400-450 triplets does not permit the analysis of the full spectrum of variability in the cerebellum. This methodological limitation likely resulted in the underestimation of the maximum allele size measured in each brain region, which, for alleles longer than ≈90 GAA•TTC repeats.

We also observed variation of the modal allele and instability profile within some of the brain regions for which multiple tissue sub-pieces were analyzed (**Supplementary Figures 17, 18, 23-25, 27, 28, 33-35**). This was particularly notable in the cerebellum and likely reflects intra-tissue heterogeneity in the distribution of allele sizes rather than technical variation between PCR replicates, which were highly reproducible (**Supplementary Figures 36-44**). The intra-tissue variability may result from sampling tissue blocks that vary in their content of neurons or glial cells, since neurons exhibit substantially higher *FGF14* expression levels than glial cells.[13]

As with blood samples, we observed a significant correlation between the degree of somatic instability and the *FGF14* GAA•TTC allele length across multiple CNS regions (**Supplementary Figure 26**). In the 12 post-mortem samples analyzed, alleles shorter than ≈90 GAA•TTC repeat units showed very low to undetectable levels of somatic expansion in all examined regions, including the cerebellum (**Supplementary Figures 45-46**). Although signal abnormality of the superior cerebellar peduncles (SCPs) has been described on imaging in a large proportion of patients with SCA27B,[14] we found no evidence of significant somatic expansion in this tissue, which had the second lowest expansion index among all central nervous system tissues studied (**Supplementary Figures 45-46**). Furthermore, we observed that some, but not all, longer alleles exhibited a pattern of somatic expansion in the dentate nucleus that was intermediate between the cerebellar cortex (vermis and hemispheres) and other central nervous system brain regions, as evidenced by a shift in the modal allele, a higher expansion index, and a broader instability profile (**Supplementary Figures 27, 29, 31, 33, 34**). The inconsistency of these findings across samples, coupled with the fact that repeated sampling of the dentate nucleus in some individuals yielded instability profiles with no evidence of significant somatic expansion, suggests that some of the analyzed tissue blocks may have been 'contaminated' with tissue from the cerebellar hemispheres due to their close anatomical proximity.

We next sought to determine if the somatic instability of the *FGF14* allele in the central nervous system was generally skewed toward contractions, as observed for peripheral blood samples. Aside from the vermis and the cerebellar hemispheres, we found that the large majority of studied tissues had a negative average instability index, indicative of an excess of contraction events, irrespective of the *FGF14* allele size (**Supplementary Figure 47**). Moreover, none of the non-cerebellar tissues consistently exhibited a positive instability index across the entire range of studied alleles, further supporting the observation that somatic instability in all tissues except the cerebellum is skewed toward contractions (**Supplementary Figures 48-49**). The overall excess of contraction in non-cerebellar regions was further qualitatively evidenced by the general appearance of the instability profiles, which showed an increased number of high-intensity peaks left to the modal alleles (**Supplementary Figures 27-35**).

Lastly, we observed that the contraction index increased linearly with repeat size in all tissues (**Supplementary Figures 50-52**). Using a multiple regression model, we found significant relationships between the contraction index and repeat size (estimate = -0.025, SE = 0.001, $p < 0.0001$), age at death (estimate = -0.027, SE = 0.013, $p = 0.036$), and tissue expression (estimate = -0.014, SE = 0.006, $p = 0.022$). In contrast, sex (estimate = 0.143, SE = 0.330, $p = 0.67$) did not have a significant effect on the contraction index. These variables together explained approximately 83% of the variance in the contraction index.

### Supplementary Figures

#### Supplementary Figure 1: Determination of the *FGF14* GAA•TTC repeat instability and contraction indices

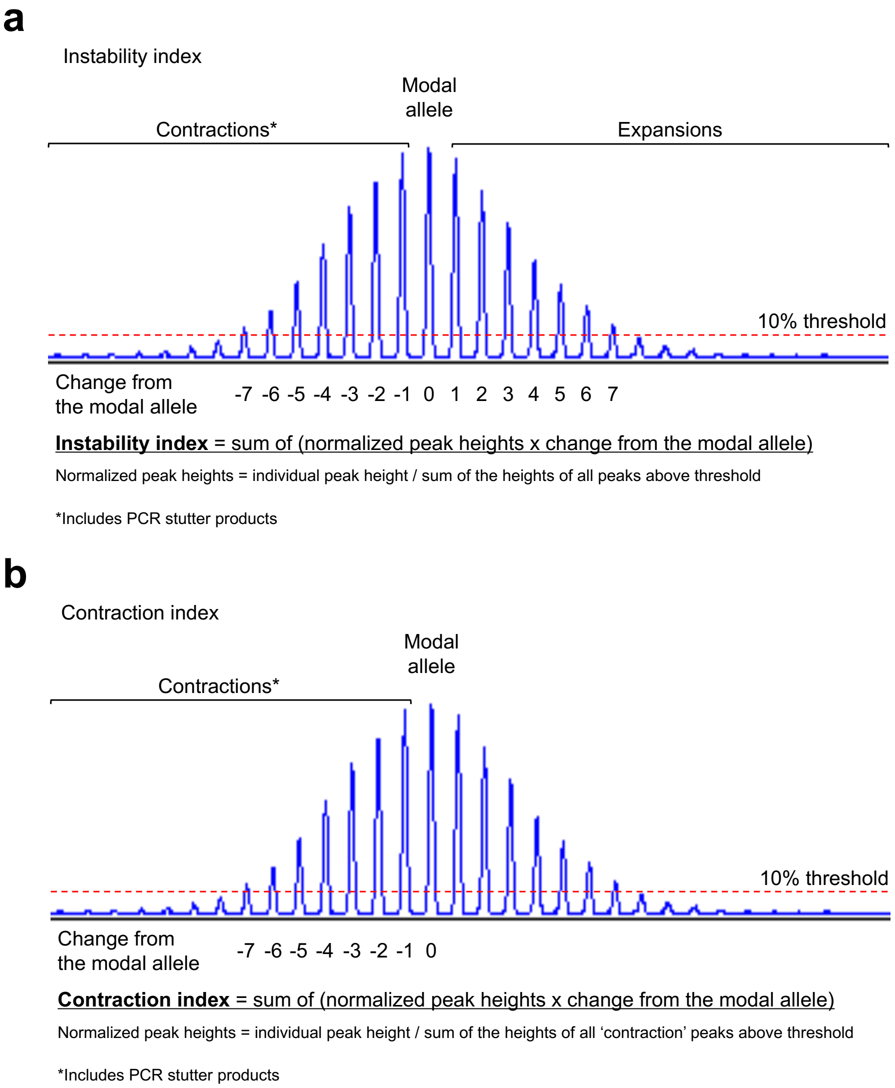


Method for calculating (**a**) the instability index and (**b**) the contraction index of the *FGF14* GAA•TTC repeat. The instability index is calculated by taking into account peaks on both sides of the modal allele and using a 10% relative peak threshold. The contraction index is calculated by only taking into account peaks to the left of the modal allele, which represent both somatically contracted GAA•TTC repeats and PCR stutter products, and using a 10% relative peak threshold. The modal allele corresponds to the peak with the highest intensity, as measured in relative fluorescence units in GeneMapper or Peak Scanner. The instability and contraction indices are calculated by first dividing the height of each individual peak by the sum of the heights of all peaks that are above the set 10% threshold. The resulting normalized peak heights are then multiplied by the position of the peak (change from the modal allele) and these values are summed to generate the index.

**Supplementary Figure 2: Analysis of the *FGF14* GAA•TTC repeat instability index in peripheral blood samples**

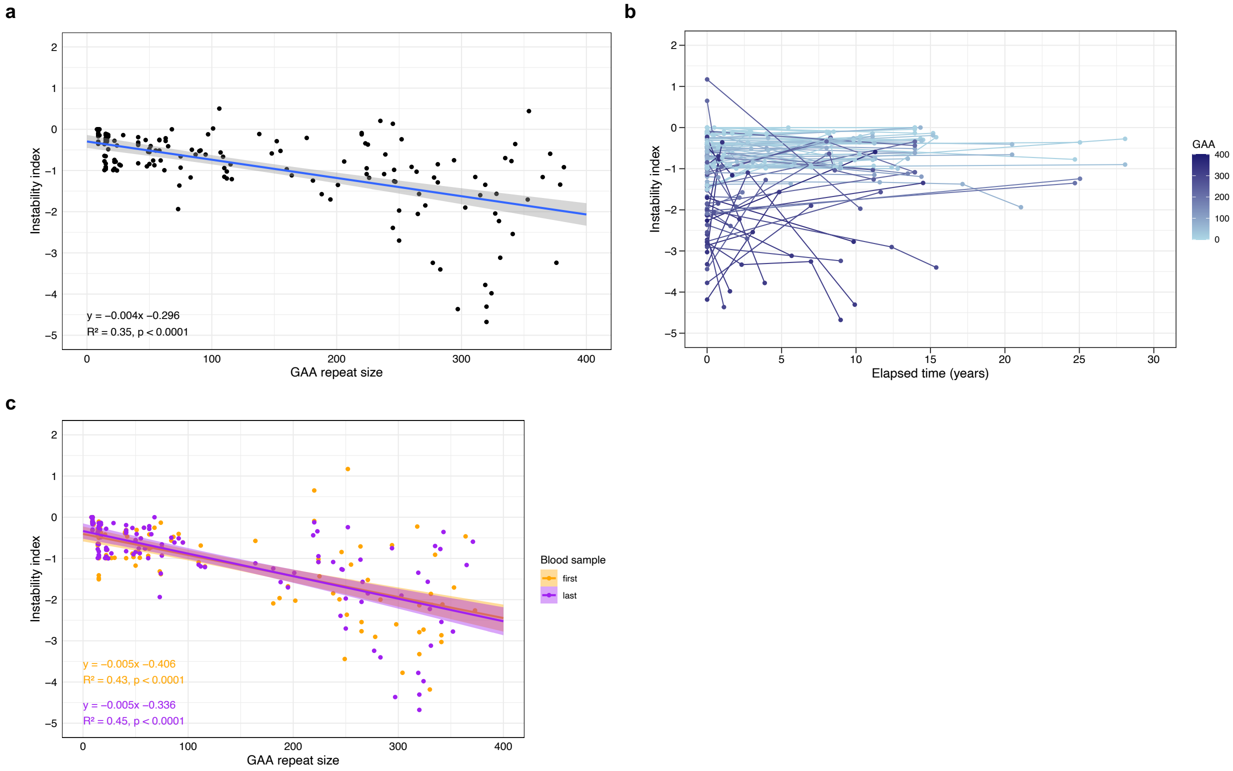


(a) Negative linear relationship between the *FGF14* GAA•TTC repeat tract size, expressed in triplet repeat counts, and instability index across 100 individuals (*n* = 173 alleles analyzed). The graph shows that 160 out of 173 alleles (92%) have a contraction bias (negative instability index), while only five alleles (3%), which are all longer than 100 triplets, have an expansion bias (positive instability index). Eight alleles (5%) showed no somatic instability in the peripheral blood. The Pearson's correlation coefficient is r = -0.59 (95% confidence interval [CI]: -0.68 to -0.48). The gray area displays the 95% confidence intervals. (b) Longitudinal analysis of the instability index across 69 individuals (*n* = 120 alleles analyzed) who underwent serial blood collections over a median period of 8.9 years (interquartile range [IQR]: 2.9 to 13.9). Observations from the same person are connected by a line. The color gradient shows the GAA•TTC allele size of each data point. (c) Negative linear relationship between the *FGF14* GAA•TTC repeat tract size and instability index in each of the first (orange) and last (purple) longitudinal blood samples across 69 individuals (*n* = 120 alleles). The Pearson's correlation coefficient is r = -0.66 (95% CI: -0.75 to -0.54) for the first blood samples and r = -0.67 (95% CI: -0.76 to -0.56) for the last blood samples. Both regression lines did not differ significantly (mixed-effect analysis, *p*=0.56), indicating relative stability of the instability index over time. The shaded areas display the 95% confidence intervals.

**Supplementary Figure 3: Analysis of the *FGF14* GAA•TTC repeat contraction index in peripheral blood samples**

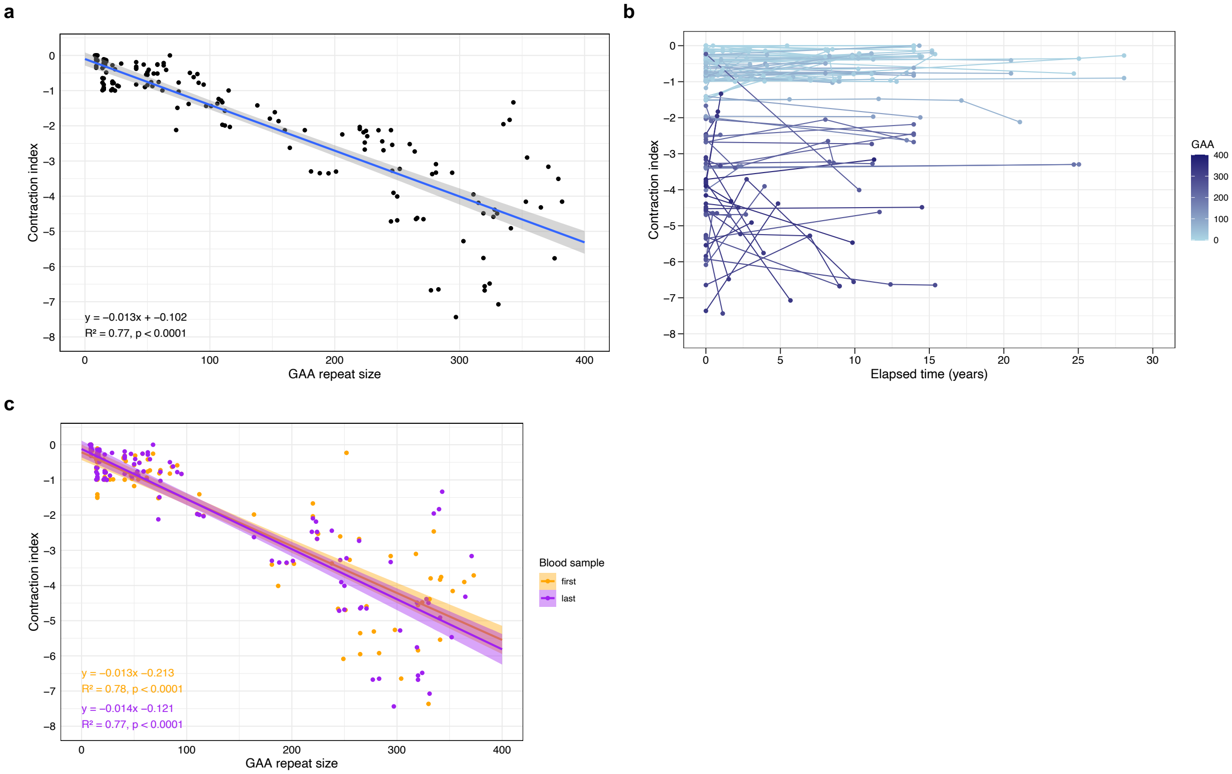


(a) Negative linear relationship between the *FGF14* GAA•TTC repeat tract size, expressed in triplet repeat counts, and contraction index across 100 individuals (*n* = 173 alleles analyzed). The Pearson's correlation coefficient is r = -0.88 (95% confidence interval [CI]: -0.91 to -0.84). The gray area displays the 95% confidence intervals. (b) Longitudinal analysis of the contraction index across 69 individuals (*n* = 120 alleles analyzed) who underwent serial blood collections over a median period of 8.9 years (interquartile range [IQR]: 2.9 to 13.9). Observations from the same person are connected by a line. The color gradient shows the GAA•TTC allele size of each data point. (c) Negative linear relationship between the *FGF14* GAA•TTC repeat tract size and instability index in each of the first (orange) and last (purple) longitudinal blood samples across 69 individuals (*n* = 120 alleles). The Pearson's correlation coefficient is r = -0.88 (95% CI: -0.92 to -0.83) for the first blood samples and r = -0.88 (95% CI: -0.91 to -0.83) for the last blood samples. Both regression lines did not differ significantly (mixed-effect analysis, *p*=0.25), indicating relative stability of the contraction index over time. The shaded areas display the 95% confidence intervals.

**Supplementary Figure 4: Modal and maximal *FGF14* GAA•TTC repeat length in peripheral blood, fibroblasts, and induced pluripotent stem cells from three patients with SCA27B**

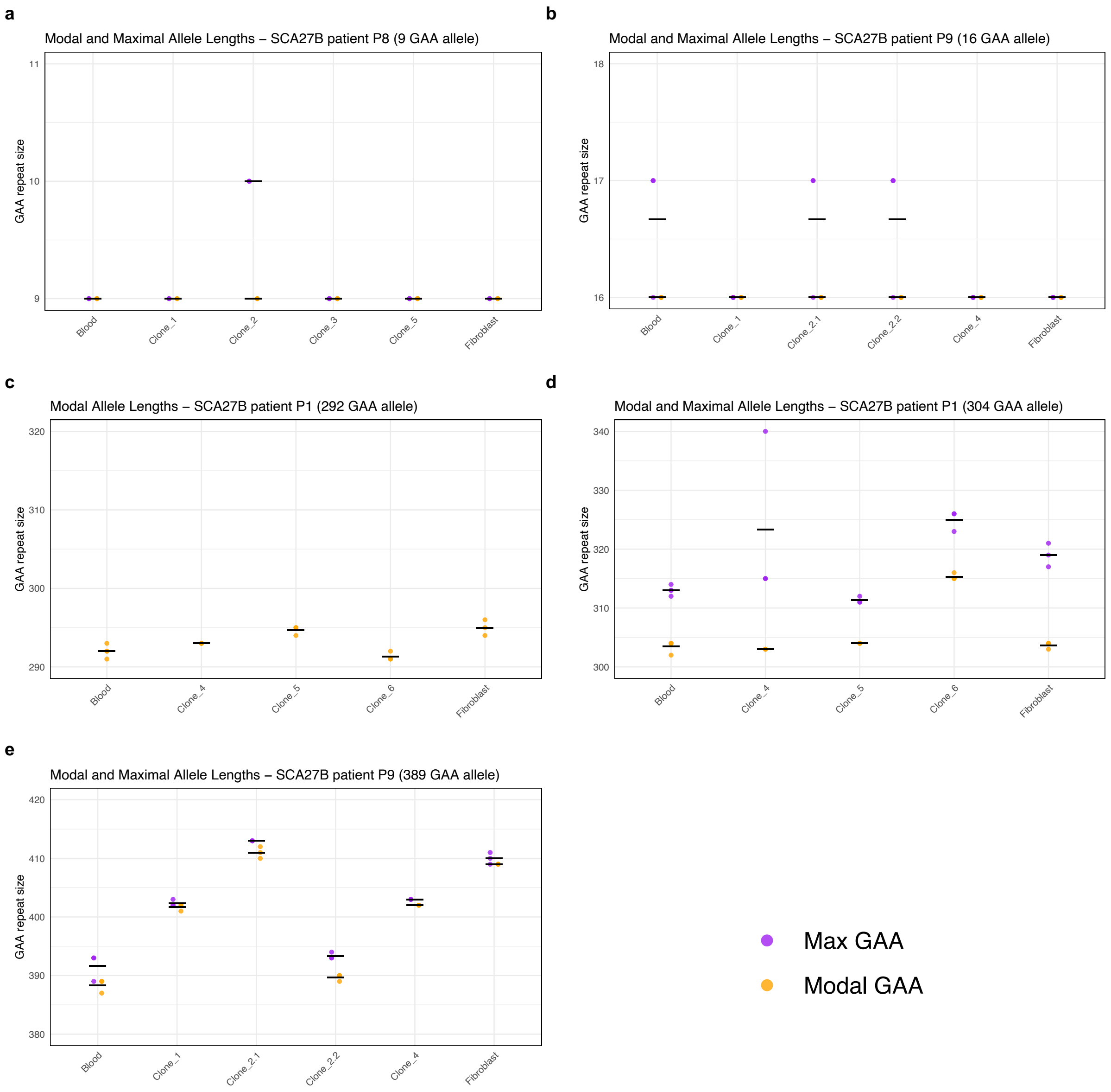


Modal and maximal GAA•TTC repeat sizes (using a 1% relative peak threshold) measured in triplicate by capillary electrophoresis of long-range PCR amplification products in peripheral blood, fibroblasts, and induced pluripotent stem cell (iPSC) clones from three patients with SCA27B. The results are shown for each individual allele, except for the longer allele of case P8, which was too long to be measured by capillary electrophoresis (see **Supplementary Figure 5**). Maximum size could not be calculated for the shorter allele of SCA27B patient P1 (292 triplets) due to the insufficient size difference with the longer allele. Modal sizes are represented by orange points, while maximum sizes are represented by purple points. Individual data points correspond to repeated measurements. Horizontal black lines show mean of replicates.

**Supplementary Figure 5: *FGF14* GAA•TTC repeat length in peripheral blood, fibroblasts, and induced pluripotent stem cell clones from SCA27B patient P8**

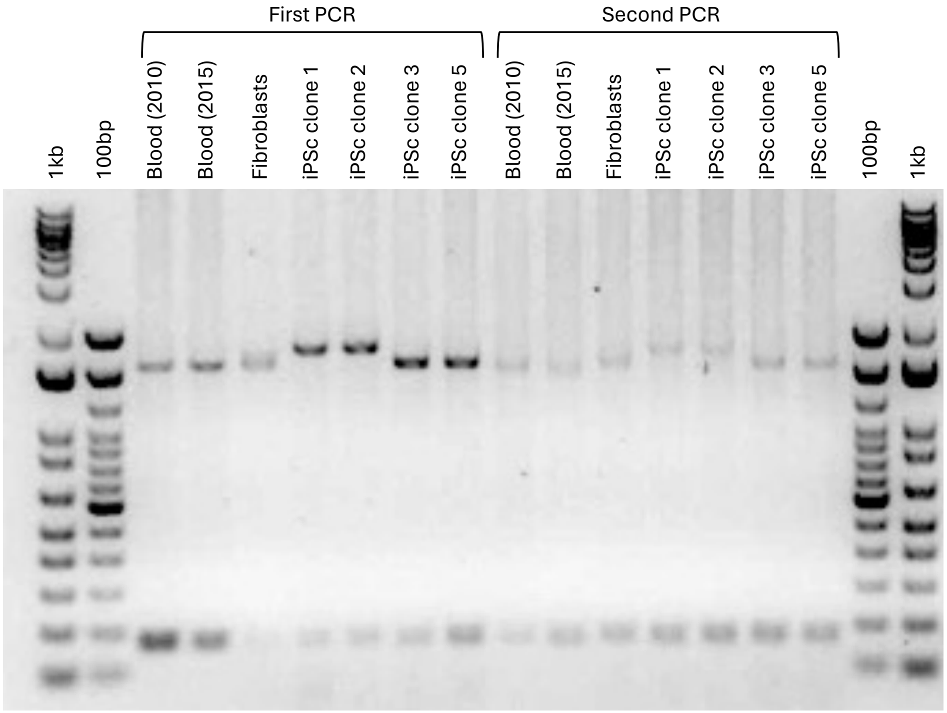


Agarose gel analysis of *FGF14* GAA•TTC repeat sizes in peripheral blood, fibroblasts, and induced pluripotent stem cells (iPSC) clones from SCA27B patient P8. PCR reactions were performed in duplicates.

**Supplementary Figure 6: Capillary electrophoresis profiles of the *FGF14* GAA•TTC repeat in blood, fibroblasts and induced pluripotent stem cell (iPSC) clones of SCA27B patient P1**

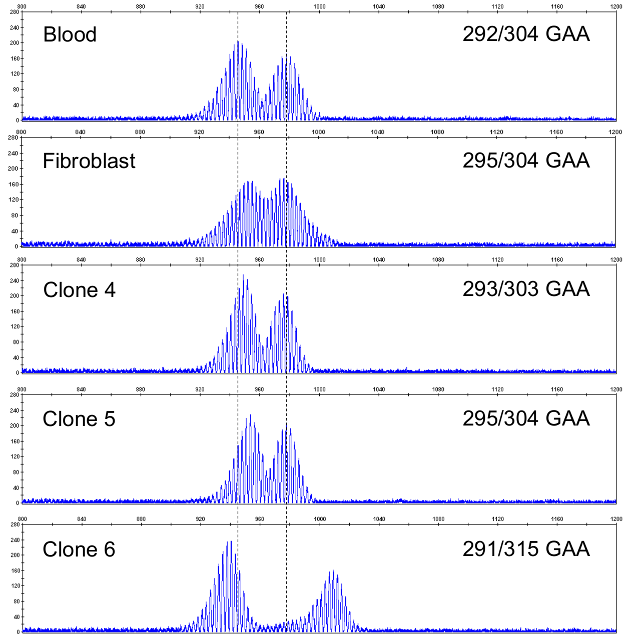


Representative capillary electrophoresis traces (visualized with the GeneMapper software) of the *FGF14* GAA•TTC repeat in blood, fibroblasts, and induced pluripotent stem cell (iPSC) clones from SCA27B patient P1. The vertical dashed black lines indicate the modal GAA•TTC alleles of 292 and 304 repeat units measured in the blood.

**Supplementary Figure 7: Stability of the *FGF14* GAA•TTC repeat during serial passages of induced pluripotent stem cells**

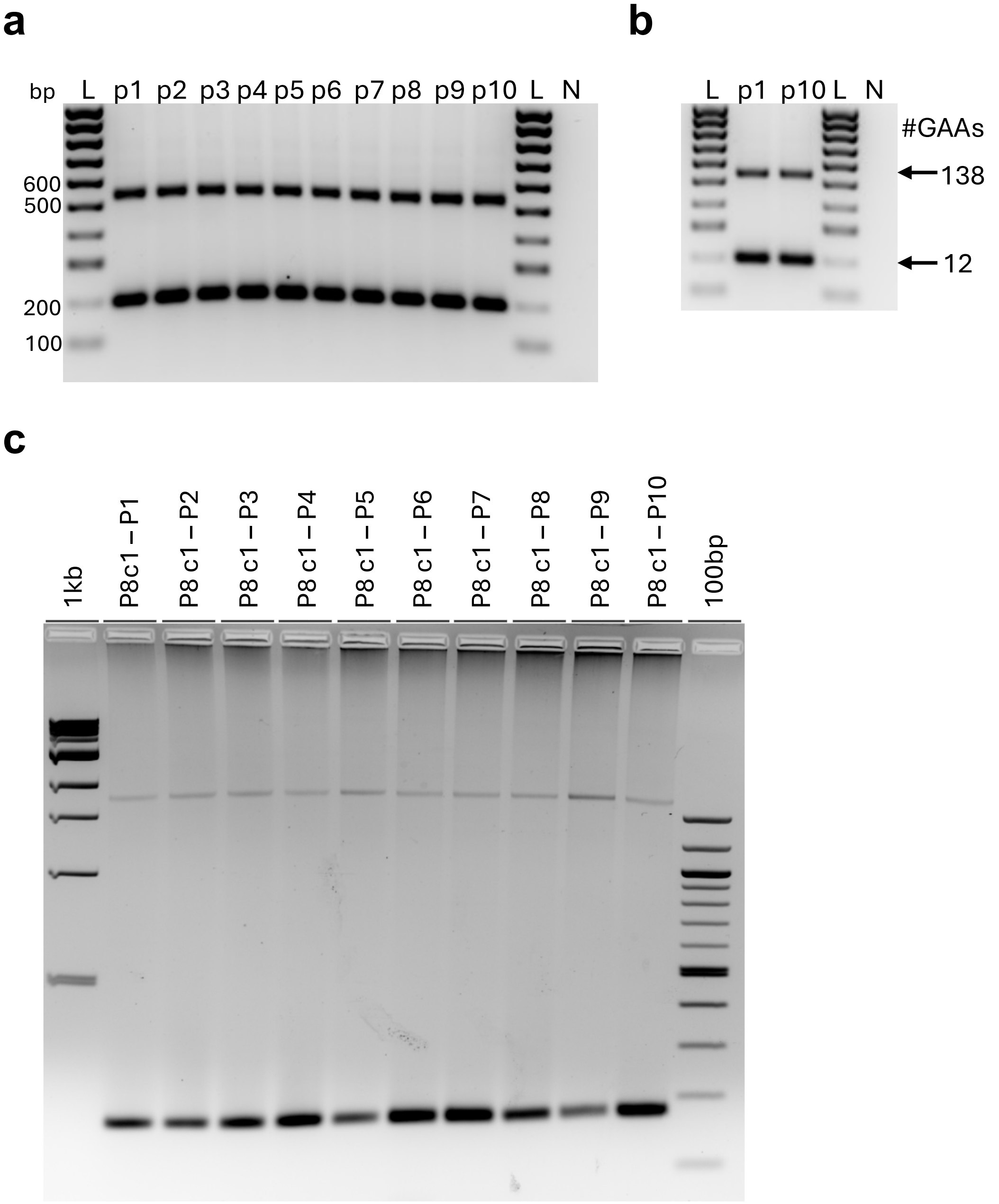


Analysis of FGF14 GAA•TTC repeat instability in induced pluripotent stem cell (iPSCs). Agarose gel analysis shows stability of *FGF14* GAA•TTC repeat sizes in serially passaged iPSC clones from (a, b) cells harboring alleles of 12 / 138 GAA•TTC repeats and (c) SCA27B patient P8 carrying 9 / 597 GAA•TTC repeats. In panel b, PCR product from passage 1 and 10 of cells shown in panel a were analyzed next to each other to verify no change of the GAA•TTC repeat number during prolonged culturing. In panel c, the stability of the shorter allele of patient Q4 was confirmed on capillary electrophoresis of long-range PCR amplification products. The length of the FGF14 GAA•TTC repeat was determined for each passage by PCR and 1.3% agarose gel electrophoresis.

**Supplementary Figure 8: Average somatic instability profiles from post-mortem central nervous system samples of control brain C1**

**a**

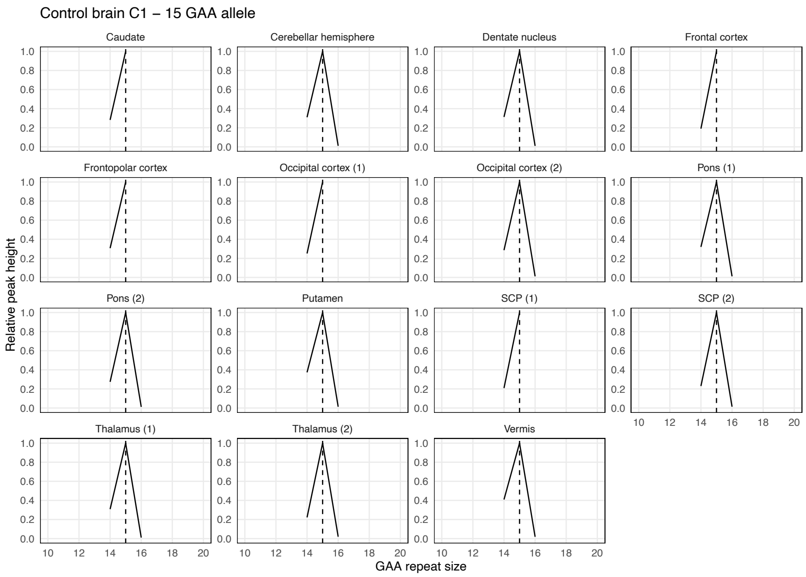


**b**

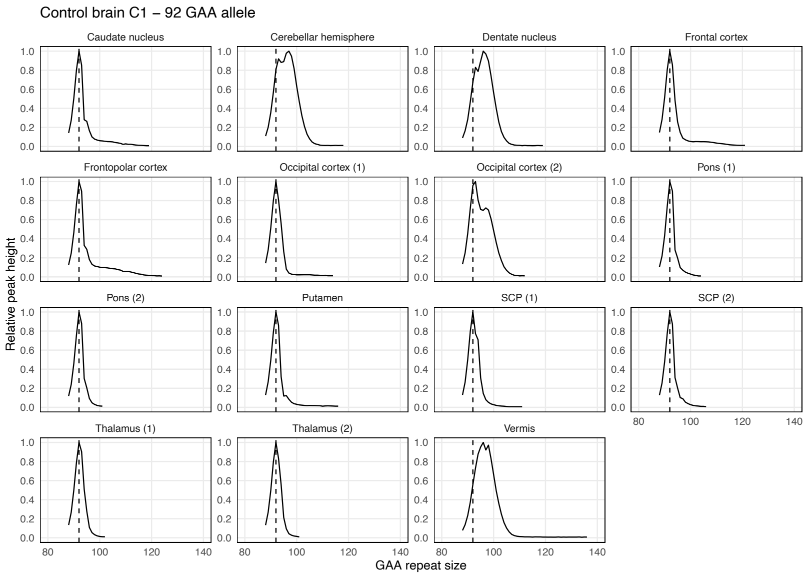


Somatic instability profiles of (a) the *FGF14* $(GAA)_{15}$ repeat allele and (b) the *FGF14* $(GAA)_{92}$ repeat allele in different brain regions, derived from post-mortem samples of control brain C1. Each plot shows the average instability profile within a given brain region, calculated from triplicate PCR reactions. For regions where multiple tissue sub-pieces were analyzed, results for each sub-piece are shown individually. Profiles were plotted by normalizing individual peak height data (extracted from the GeneMapper software) to the height of the modal allele within each brain region. Peaks left of the modal allele above a 10% threshold and those right of the modal allele above a 1% threshold were plotted. Vertical dashed black lines indicate the size of the modal alleles measured in non-cerebellar regions. SCP: superior cerebellar peduncles.

**Supplementary Figure 9: Average somatic instability profiles from post-mortem central nervous system samples of control brain C2**

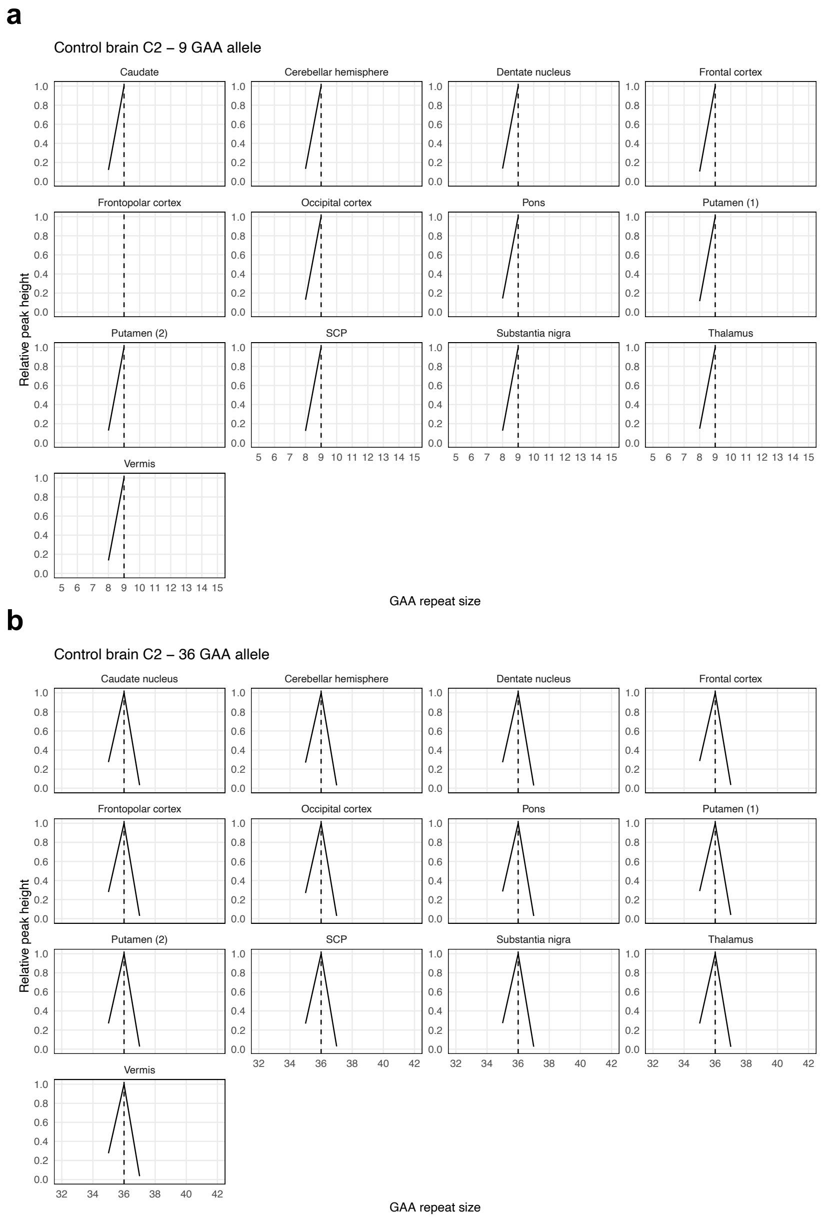


Somatic instability profiles of (a) the *FGF14* $(GAA)_9$ repeat allele and (b) the *FGF14* $(GAA)_{36}$ repeat allele in different brain regions, derived from post-mortem samples of control brain C2. Each plot shows the average instability profile within a given brain region, calculated from triplicate PCR reactions. For regions where multiple tissue sub-pieces were analyzed, results for each sub-piece are shown individually. Profiles were plotted by normalizing individual peak height data (extracted from the GeneMapper software) to the height of the modal allele within each brain region. Peaks left of the modal allele above a 10% threshold and those right of the modal allele above a 1% threshold were plotted. Vertical dashed black lines indicate the size of the modal alleles measured in non-cerebellar regions. SCP: superior cerebellar peduncles.

**Supplementary Figure 10: Average somatic instability profiles from post-mortem central nervous system samples of control brain C3**

**a**

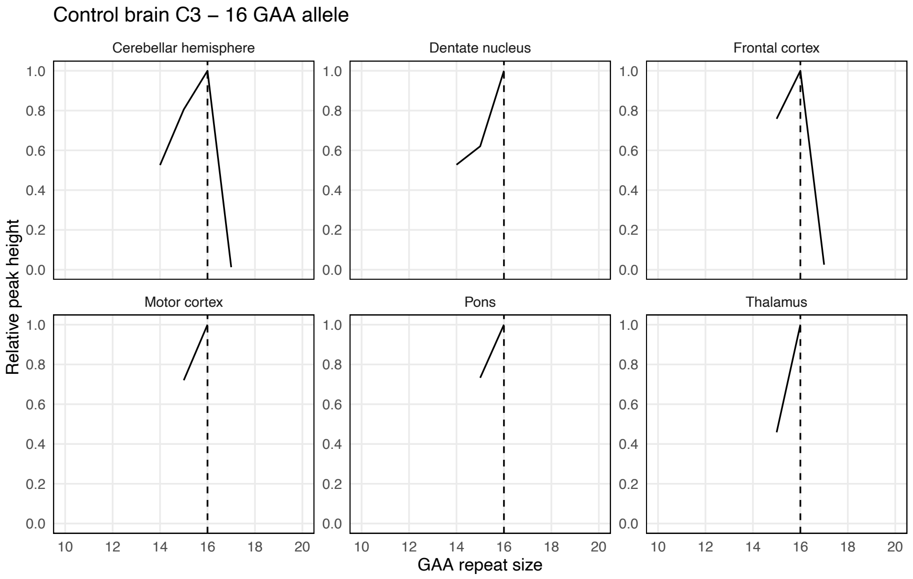


**b**

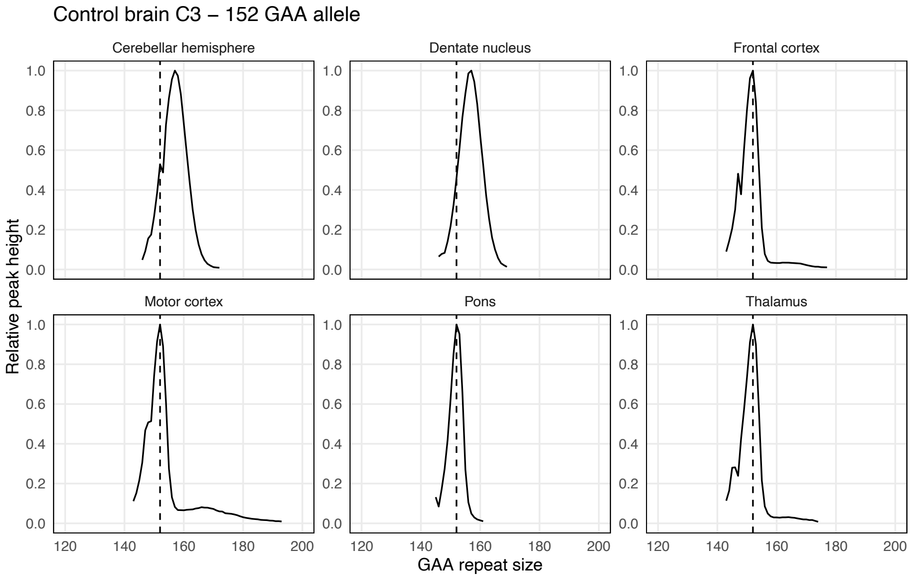


Somatic instability profiles of (a) the *FGF14* $(GAA)_{16}$ repeat allele and (b) the *FGF14* $(GAA)_{152}$ repeat allele in different brain regions, derived from post-mortem samples of control brain C3. Each plot shows the average instability profile within a given brain region, calculated from duplicate PCR reactions. Profiles were plotted by normalizing individual peak height data (extracted from the Peak Scanner software) to the height of the modal allele within each brain region. Peaks left of the modal allele above a 10% threshold and those right of the modal allele above a 1% threshold were plotted. Vertical dashed black lines indicate the size of the modal alleles measured in non-cerebellar regions.

**Supplementary Figure 11: Average somatic instability profiles from post-mortem central nervous system samples of control brain C4**

**a**

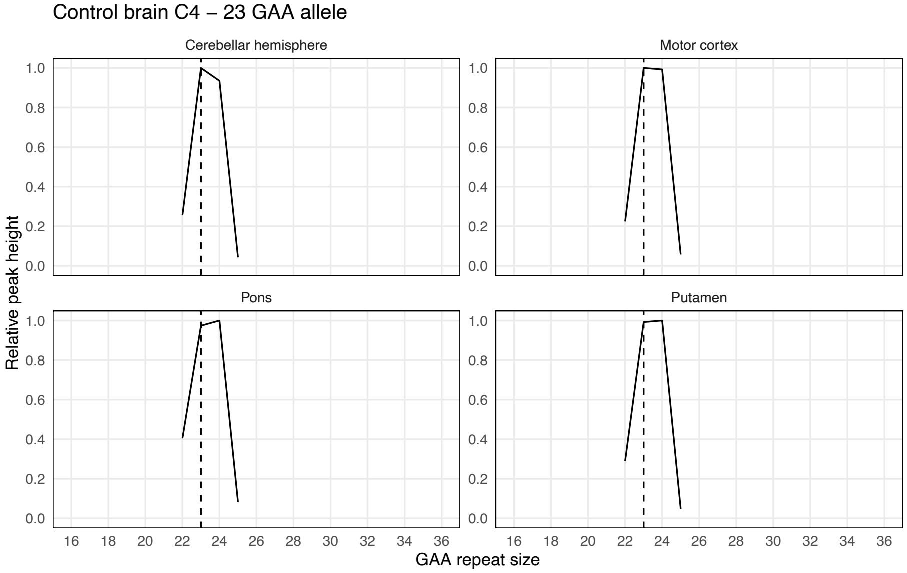


**b**

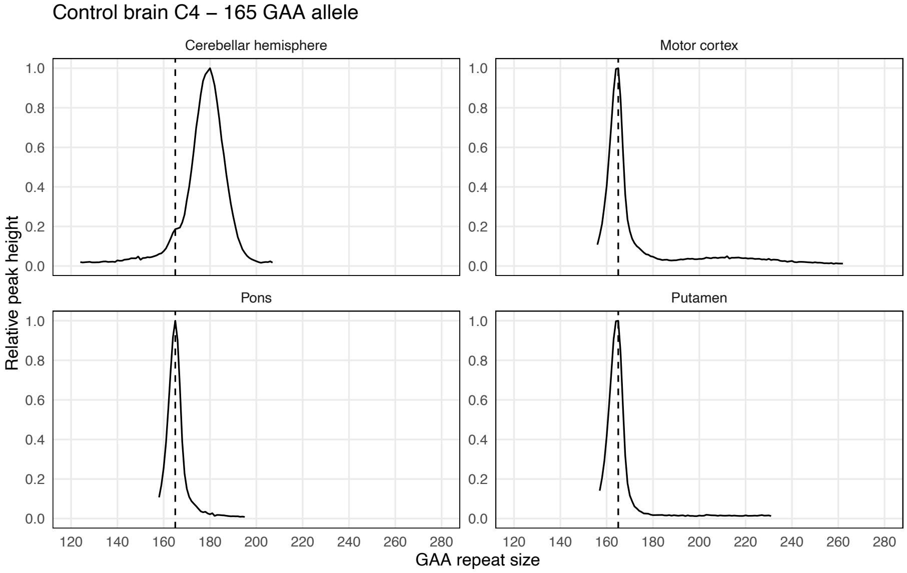


Somatic instability profiles of (a) the *FGF14* $(GAA)_{23}$ repeat allele and (b) the *FGF14* $(GAA)_{165}$ repeat allele in different brain regions, derived from post-mortem samples of control brain C4. Each plot shows the average instability profile within a given brain region, calculated from duplicate PCR reactions. Profiles were plotted by normalizing individual peak height data (extracted from the Peak Scanner software) to the height of the modal allele within each brain region. Peaks left of the modal allele above a 10% threshold and those right of the modal allele above a 1% threshold were plotted. Vertical dashed black lines indicate the size of the modal alleles measured in non-cerebellar regions.

**Supplementary Figure 12: Average somatic instability profiles from post-mortem central nervous system samples of control brain C5**

**a**

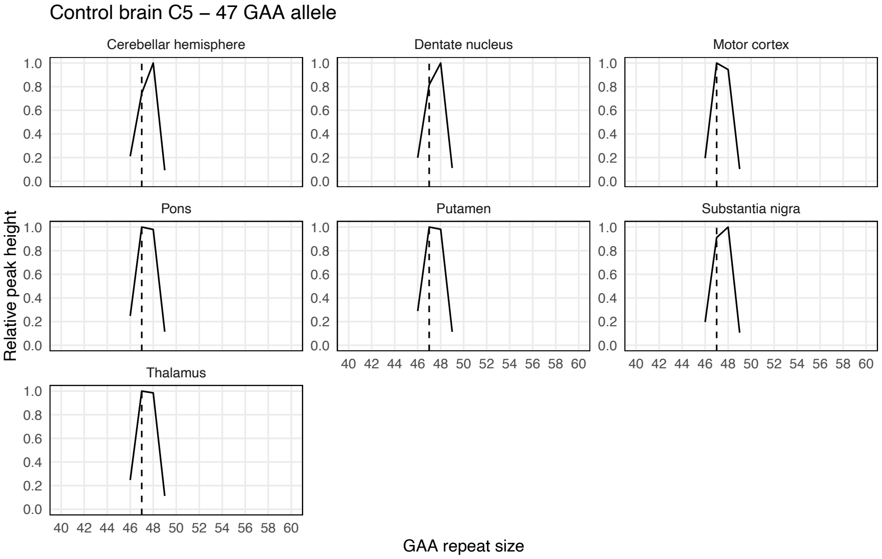


**b**

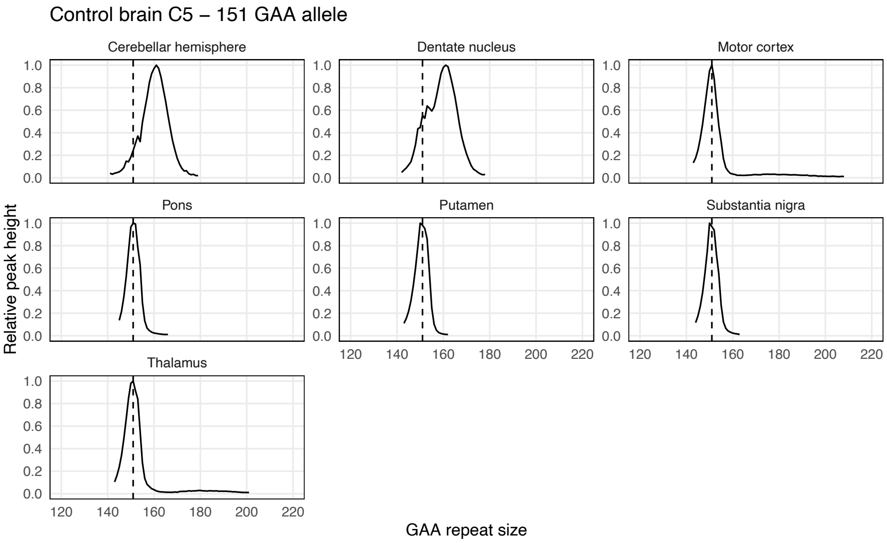


Somatic instability profiles of (a) the *FGF14* $(GAA)_{47}$ repeat allele and (b) the *FGF14* $(GAA)_{151}$ repeat allele in different brain regions, derived from post-mortem samples of control brain C5. Each plot shows the average instability profile within a given brain region, calculated from duplicate PCR reactions. Profiles were plotted by normalizing individual peak height data (extracted from the Peak Scanner software) to the height of the modal allele within each brain region. Peaks left of the modal allele above a 10% threshold and those right of the modal allele above a 1% threshold were plotted. Vertical dashed black lines indicate the size of the modal alleles measured in non-cerebellar regions.

**Supplementary Figure 13: Average somatic instability profiles from post-mortem central nervous system samples of control brain C6**

**a**

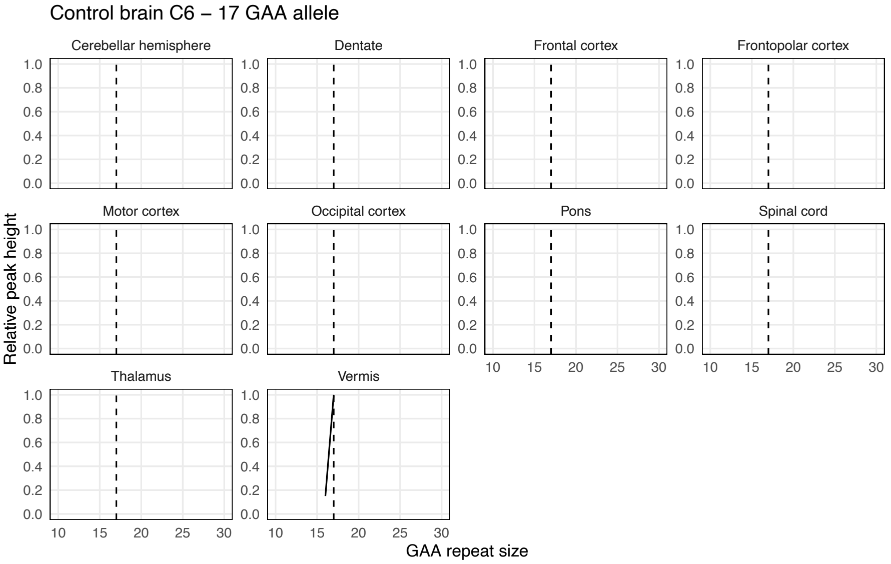


**b**

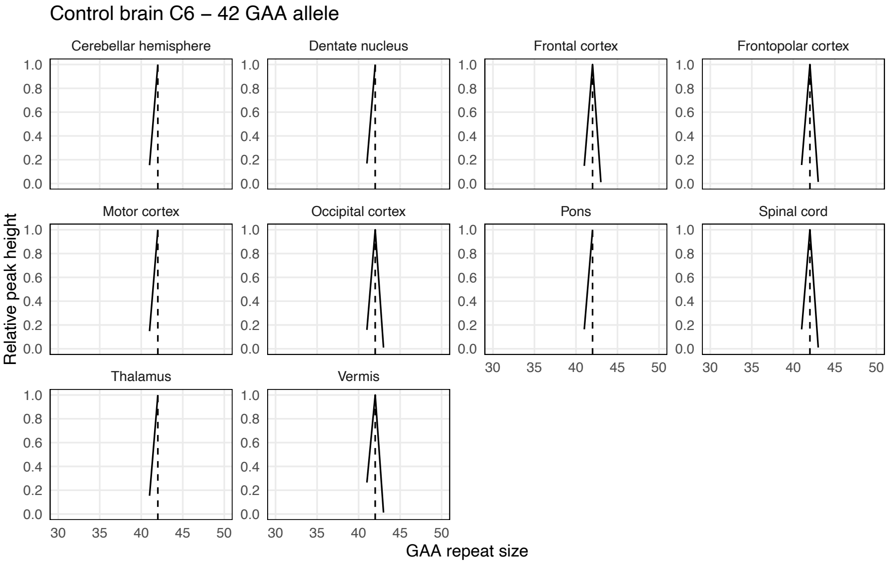


Somatic instability profiles of (a) the *FGF14* $(GAA)_{17}$ repeat allele and (b) the *FGF14* $(GAA)_{42}$ repeat allele in different brain regions, derived from post-mortem samples of control brain C6. Each plot shows the average instability profile within a given brain region. Profiles were plotted by normalizing individual peak height data (extracted from the Peak Scanner software) to the height of the modal allele within each brain region. Peaks left of the modal allele above a 10% threshold and those right of the modal allele above a 1% threshold were plotted. Vertical dashed black lines indicate the size of the modal alleles measured in non-cerebellar regions.

**Supplementary Figure 14: Average somatic instability profiles from post-mortem central nervous system samples of SCA27B patient brain P1**

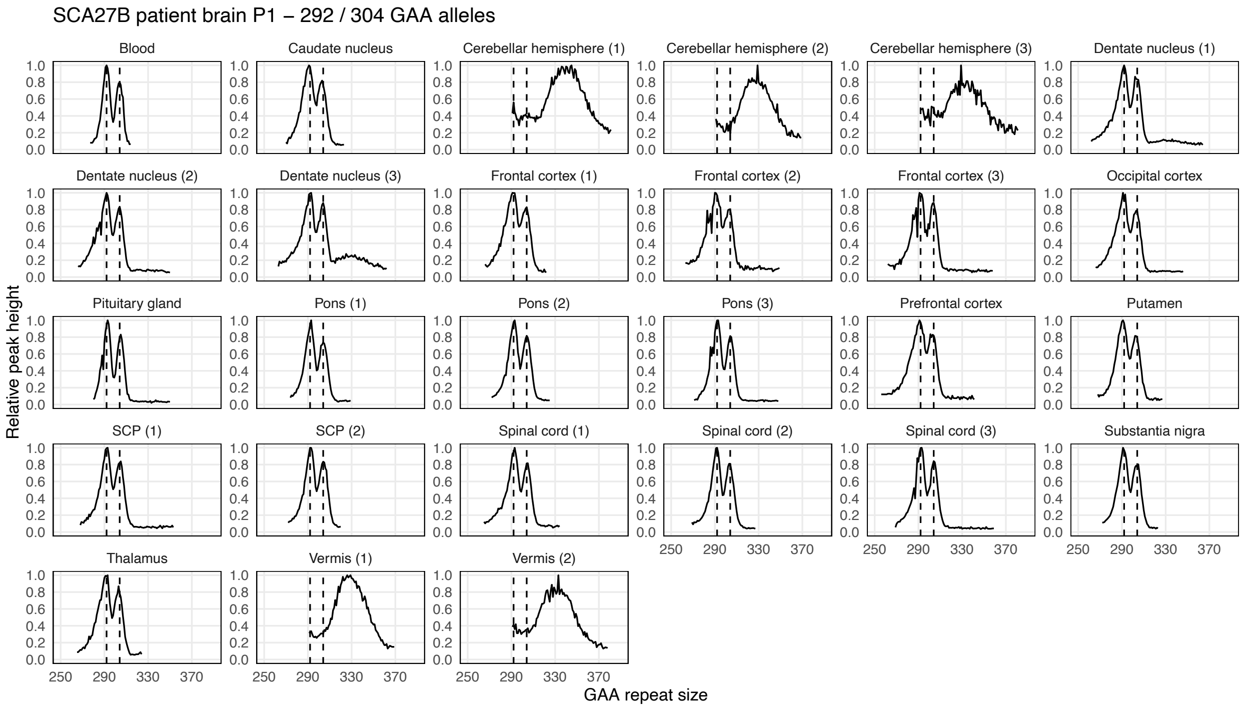


Somatic instability profiles of the *FGF14* $(GAA)_{292}$ and $(GAA)_{304}$ repeat alleles in different brain regions, derived from post-mortem samples of SCA27B patient brain P1. Each plot shows the average instability profile within a given brain region, calculated from triplicate PCR reactions. For regions where multiple tissue sub-pieces were analyzed, results for each sub-piece are shown individually. Profiles were plotted by normalizing individual peak height data (extracted from the GeneMapper software) to the height of the modal peak of the shorter allele within each brain region. Peaks left of the modal allele above a 10% threshold and those right of the modal allele above a 1% threshold were plotted. Vertical dashed black lines indicate the size of the modal alleles measured in the blood. SCP: superior cerebellar peduncles.

**Supplementary Figure 15: Average somatic instability profiles from post-mortem central nervous system samples of SCA27B patient brain P2**

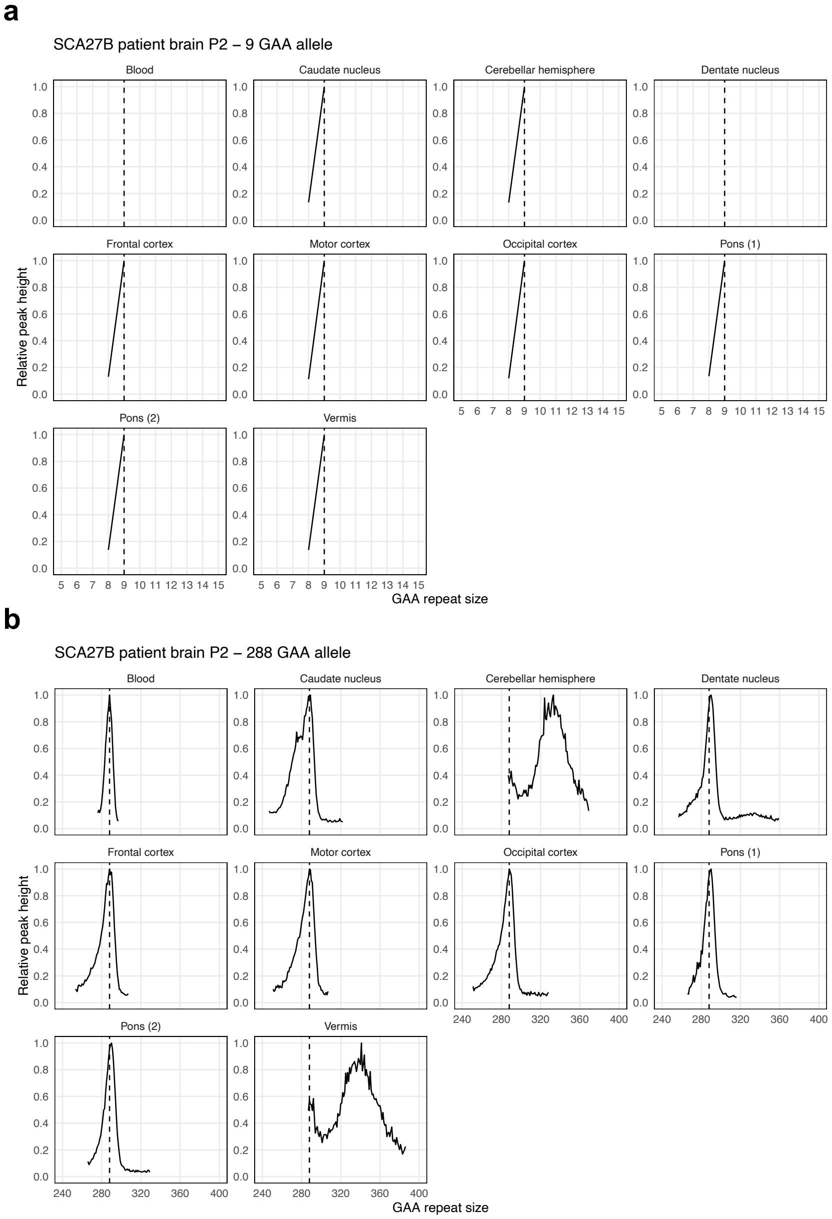


Somatic instability profiles of (a) the *FGF14* $(GAA)_9$ repeat allele and (b) the *FGF14* $(GAA)_{288}$ repeat allele in different brain regions, derived from post-mortem samples of SCA27B patient brain P2. Each plot shows the average instability profile within a given brain region, calculated from triplicate PCR reactions. For regions where multiple tissue sub-pieces were analyzed, results for each sub-piece are shown individually. Profiles were plotted by normalizing individual peak height data (extracted from the GeneMapper software) to the height of the modal allele within each brain region. Peaks left of the modal allele above a 10% threshold and those right of the modal allele above a 1% threshold were plotted. Vertical dashed black lines indicate the size of the modal alleles measured in the blood.

**Supplementary Figure 16: Average somatic instability profiles from post-mortem central nervous system samples of SCA27B patient brain P3**

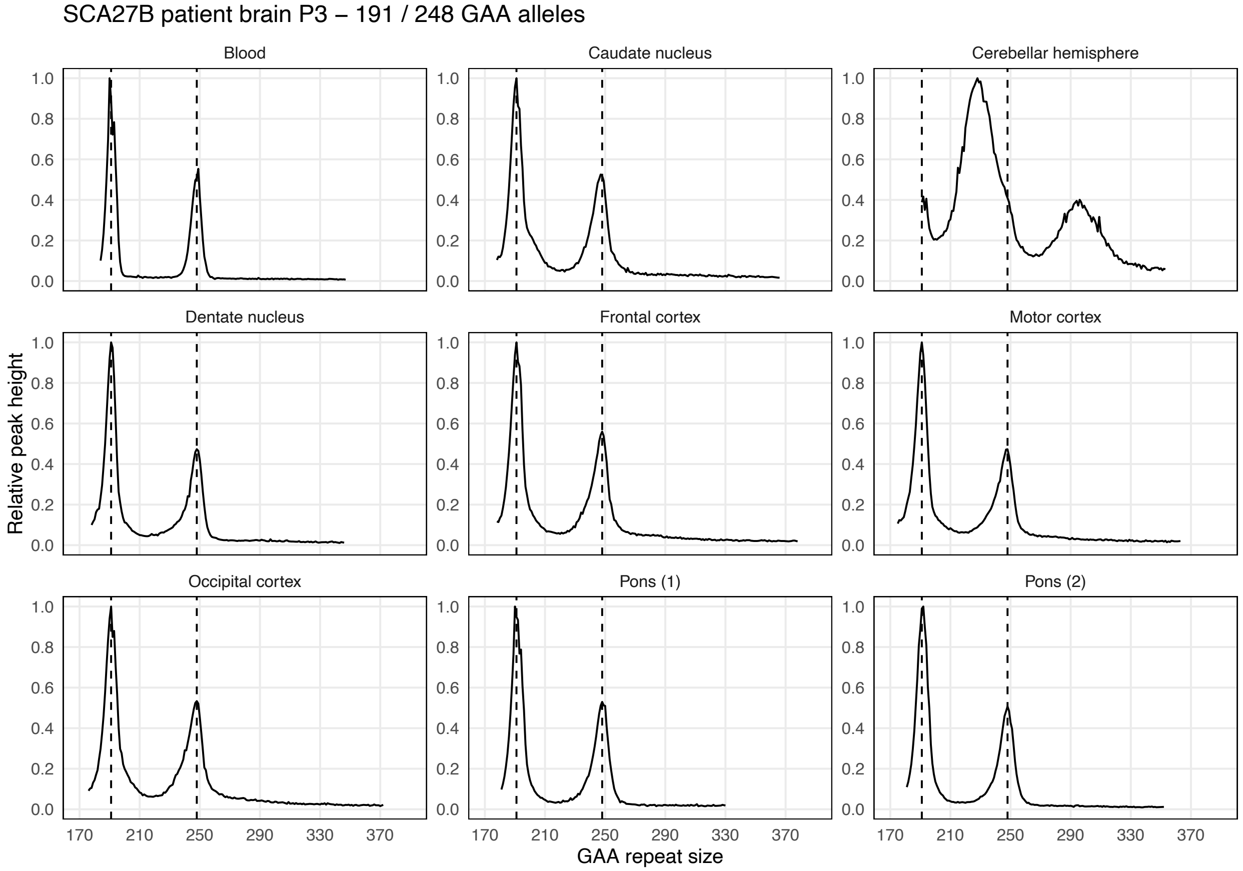


Somatic instability profiles of the *FGF14* $(GAA)_{191}$ and $(GAA)_{248}$ repeat alleles in different brain regions, derived from post-mortem samples of SCA27B patient brain P3. Each plot shows the average instability profile within a given brain region, calculated from triplicate PCR reactions. For regions where multiple tissue sub-pieces were analyzed, results for each sub-piece are shown individually. Profiles were plotted by normalizing individual peak height data (extracted from the GeneMapper software) to the height of the modal peak of the shorter allele within each brain region. Peaks left of the modal allele above a 10% threshold and those right of the modal allele above a 1% threshold were plotted. Vertical dashed black lines indicate the size of the modal alleles measured in the blood.

**Supplementary Figure 17: Modal and maximal *FGF14* GAA•TTC repeat length in post-mortem central nervous system tissues from control brain C1**

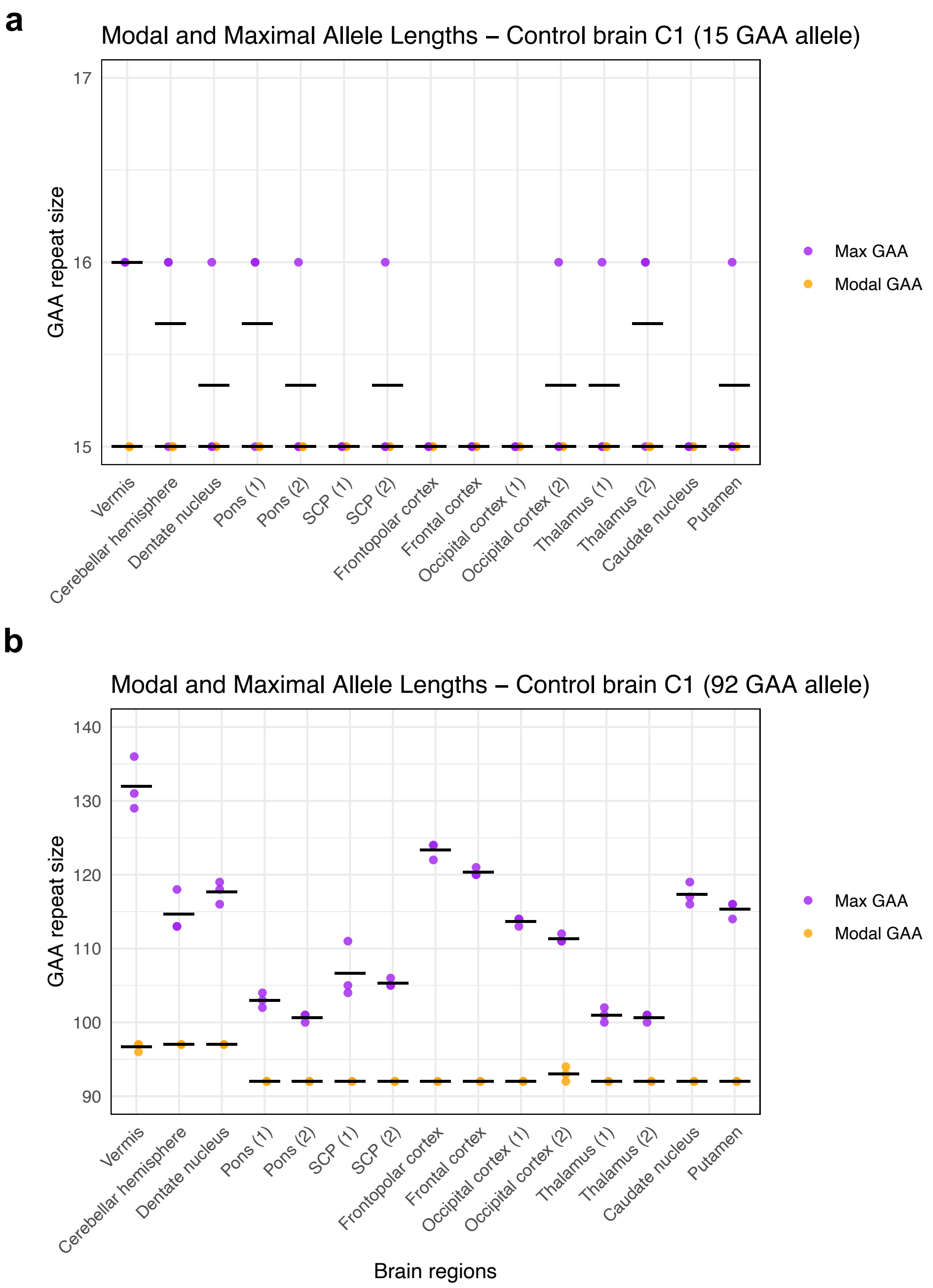


Modal and maximal GAA•TTC repeat sizes (using a 1% relative peak threshold) measured in triplicate by capillary electrophoresis of long-range PCR amplification products in post-mortem central nervous system regions from control brain C1. Results for the short and long allele are shown in panels a and b, respectively. Modal sizes are represented by orange points, while maximum sizes are represented by purple points. Individual data points correspond to repeated measurements. Horizontal black lines show mean of replicates. SCP: superior cerebellar peduncles.

**Supplementary Figure 18: Modal and maximal *FGF14* GAA•TTC repeat length in post-mortem central nervous system tissues from control brain C2**

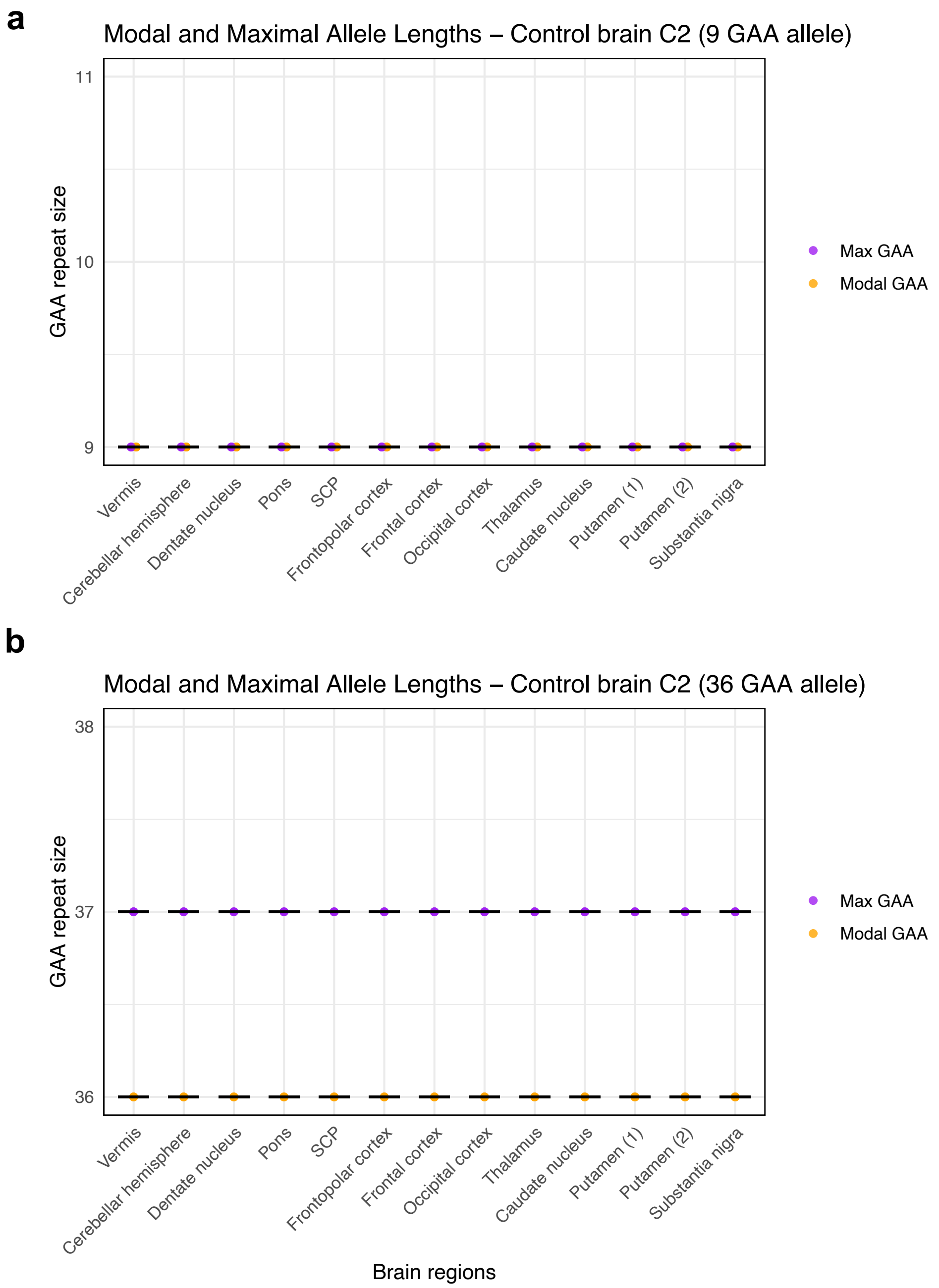


Modal and maximal GAA•TTC repeat sizes (using a 1% relative peak threshold) measured in triplicate by capillary electrophoresis of long-range PCR amplification products in post-mortem central nervous system regions from control brain C2. Results for the short and long allele are shown in panels a and b, respectively. Modal sizes are represented by orange points, while maximum sizes are represented by purple points. Individual data points correspond to repeated measurements. Horizontal black lines show mean of replicates. SCP: superior cerebellar peduncles.

**Supplementary Figure 19: Modal and maximal *FGF14* GAA•TTC repeat length in post-mortem central nervous system tissues from control brain C3**

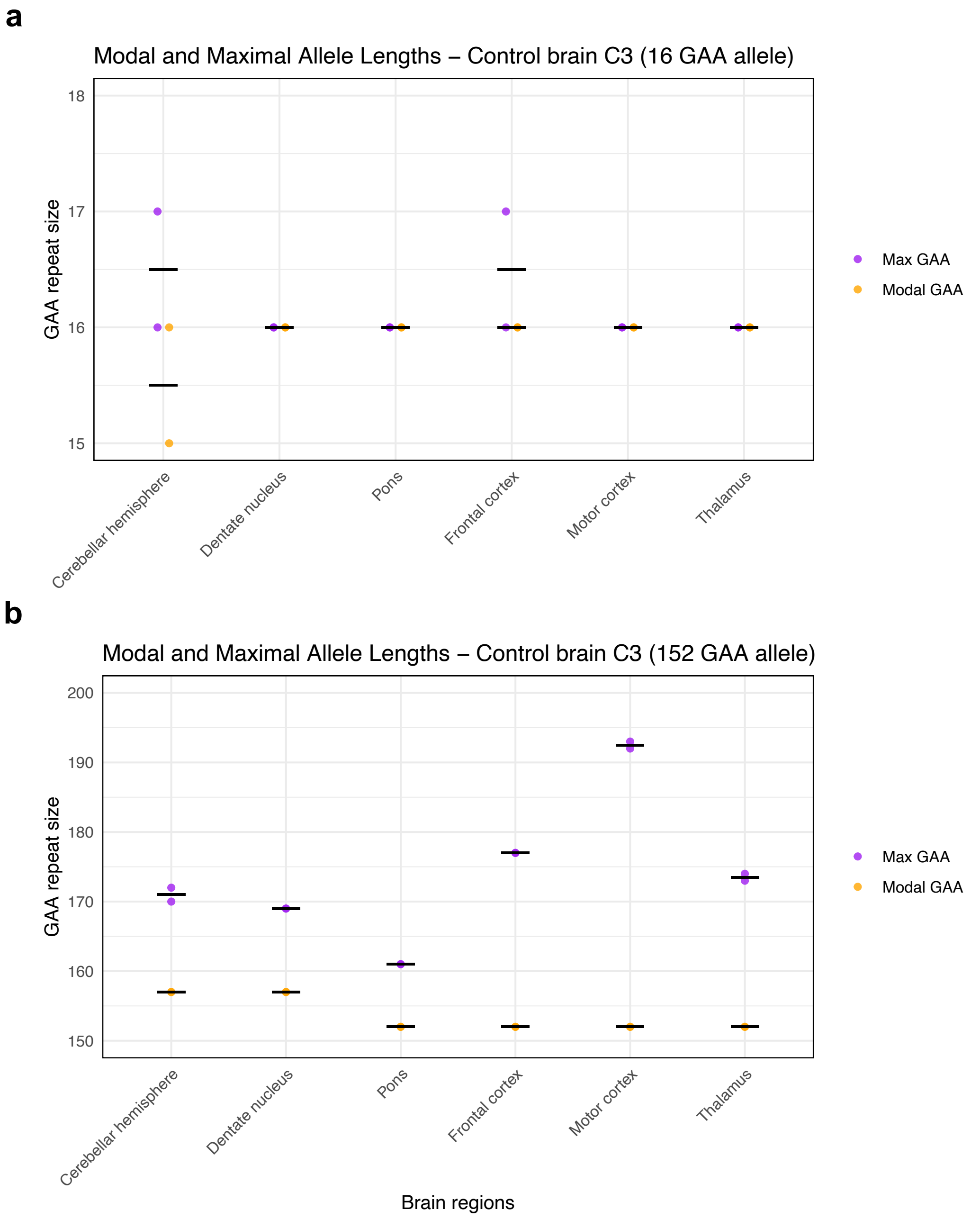


Modal and maximal GAA•TTC repeat sizes (using a 1% relative peak threshold) measured in duplicate by capillary electrophoresis of long-range PCR amplification products in post-mortem central nervous system regions from control brain C3. Results for the short and long allele are shown in panels a and b, respectively. Modal sizes are represented by orange points, while maximum sizes are represented by purple points. Individual data points correspond to repeated measurements. Horizontal black lines show mean of replicates.

**Supplementary Figure 20: Modal and maximal *FGF14* GAA•TTC repeat length in post-mortem central nervous system tissues from control brain C4**

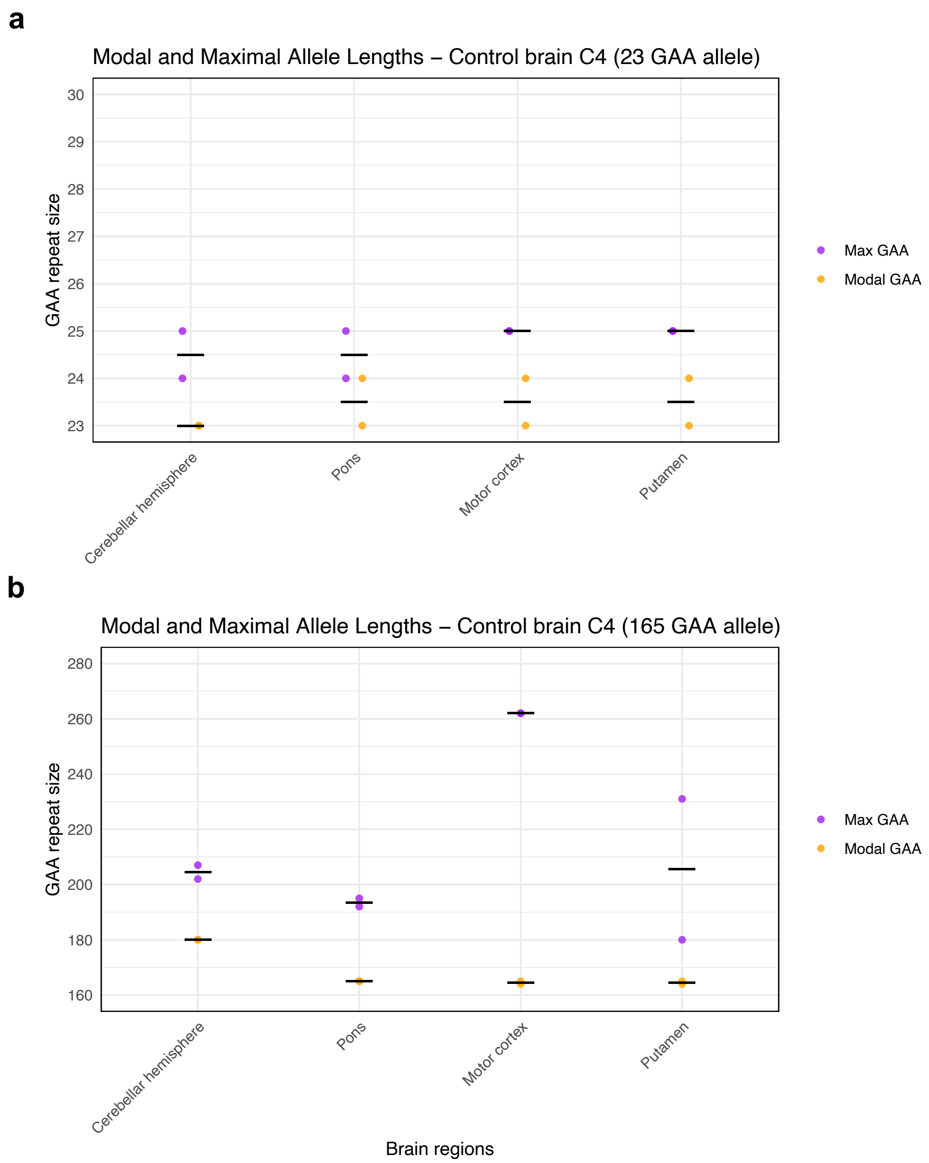


Modal and maximal GAA•TTC repeat sizes (using a 1% relative peak threshold) measured in duplicate by capillary electrophoresis of long-range PCR amplification products in post-mortem central nervous system regions from control brain C4. Results for the short and long allele are shown in panels a and b, respectively. Modal sizes are represented by orange points, while maximum sizes are represented by purple points. Individual data points correspond to repeated measurements. Horizontal black lines show mean of replicates.

**Supplementary Figure 21: Modal and maximal *FGF14* GAA•TTC repeat length in post-mortem central nervous system tissues from control brain C5**

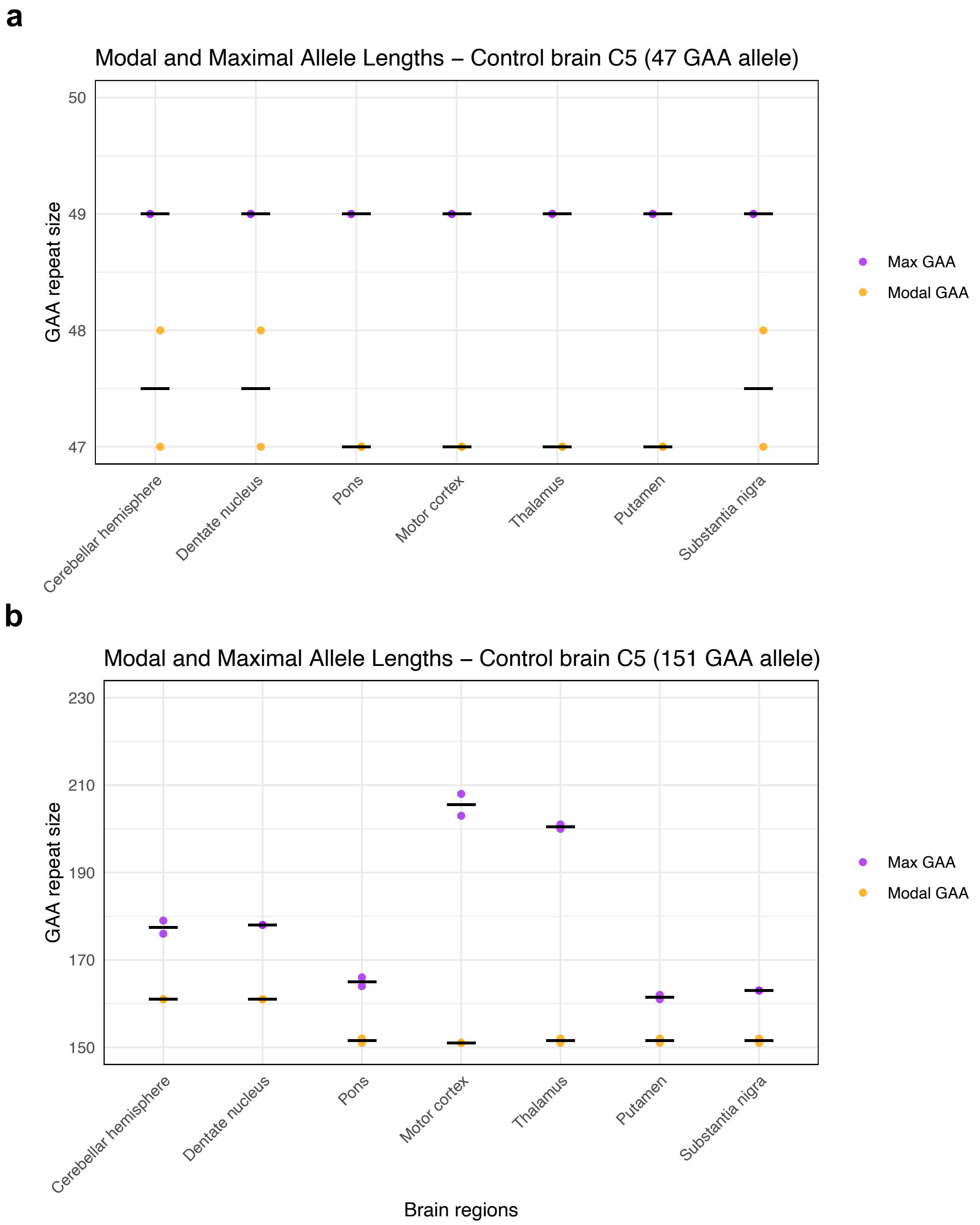


Modal and maximal GAA•TTC repeat sizes (using a 1% relative peak threshold) measured in duplicate by capillary electrophoresis of long-range PCR amplification products in post-mortem central nervous system regions from control brain C5. Results for the short and long allele are shown in panels a and b, respectively. Modal sizes are represented by orange points, while maximum sizes are represented by purple points. Individual data points correspond to repeated measurements. Horizontal black lines show mean of replicates.

**Supplementary Figure 22: Modal and maximal *FGF14* GAA•TTC repeat length in post-mortem central nervous system tissues from control brain C6**

a

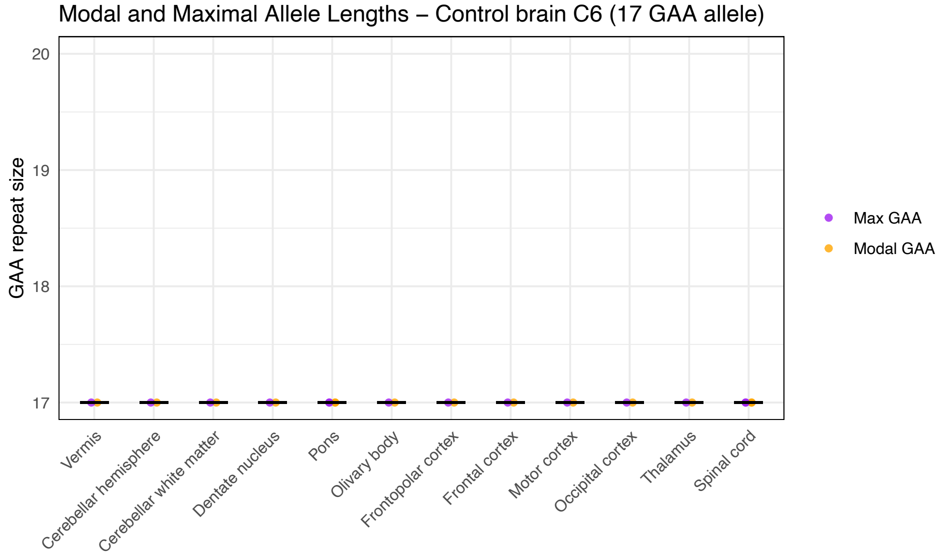


b

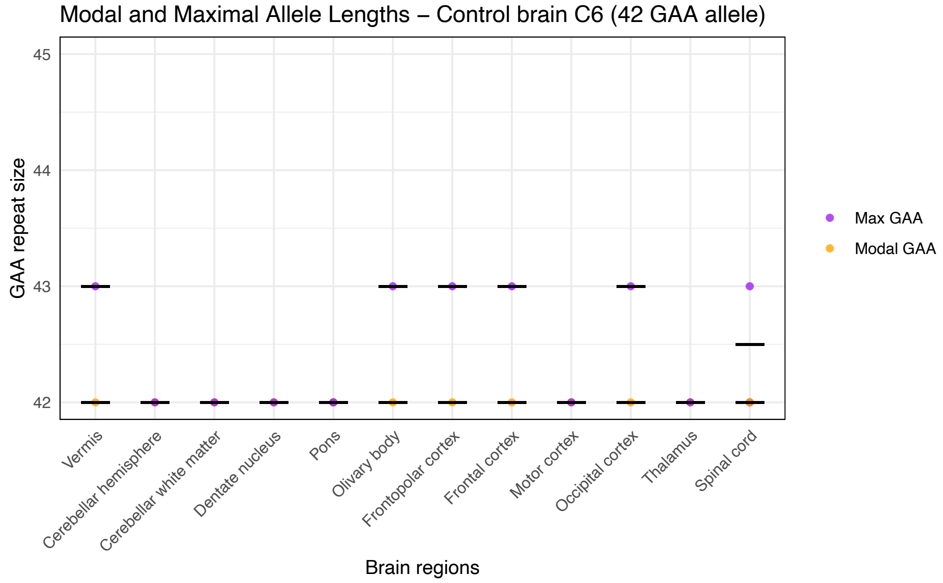


Modal and maximal GAA•TTC repeat sizes (using a 1% relative peak threshold) measured by capillary electrophoresis of long-range PCR amplification products in post-mortem central nervous system regions from control brain C6. Results for the short and long allele are shown in panels a and b, respectively. Modal sizes are represented by orange points, while maximum sizes are represented by purple points. Individual data points correspond to repeated measurements. Horizontal black lines show mean of replicates.

**Supplementary Figure 23: Modal and maximal *FGF14* GAA•TTC repeat length in post-mortem central nervous system tissues from SCA27B patient brain P1**

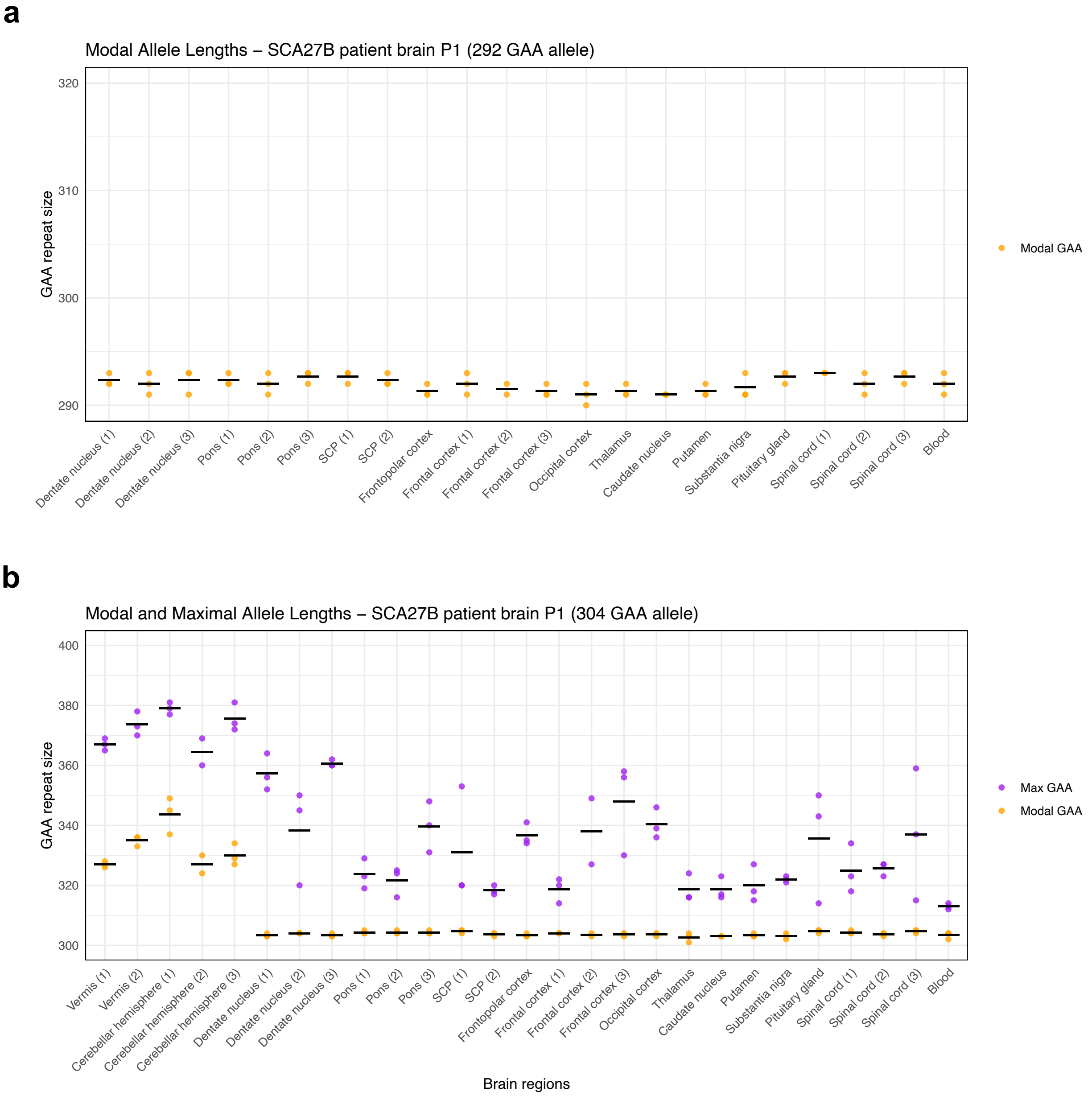


Modal and maximal GAA•TTC repeat sizes (using a 1% relative peak threshold) measured in triplicate by capillary electrophoresis of long-range PCR amplification products in post-mortem central nervous system regions from SCA27B patient brain P1. Results for the short and long allele are shown in panels a and b, respectively. Maximum size could not be calculated for the short allele due to the insufficient size difference with the long allele. Modal sizes are represented by orange points, while maximum sizes are represented by purple points. Individual data points correspond to repeated measurements. Horizontal black lines show mean of replicates. SCP: superior cerebellar peduncles.

**Supplementary Figure 24: Modal and maximal *FGF14* GAA•TTC repeat length in post-mortem central nervous system tissues from SCA27B patient brain P2**

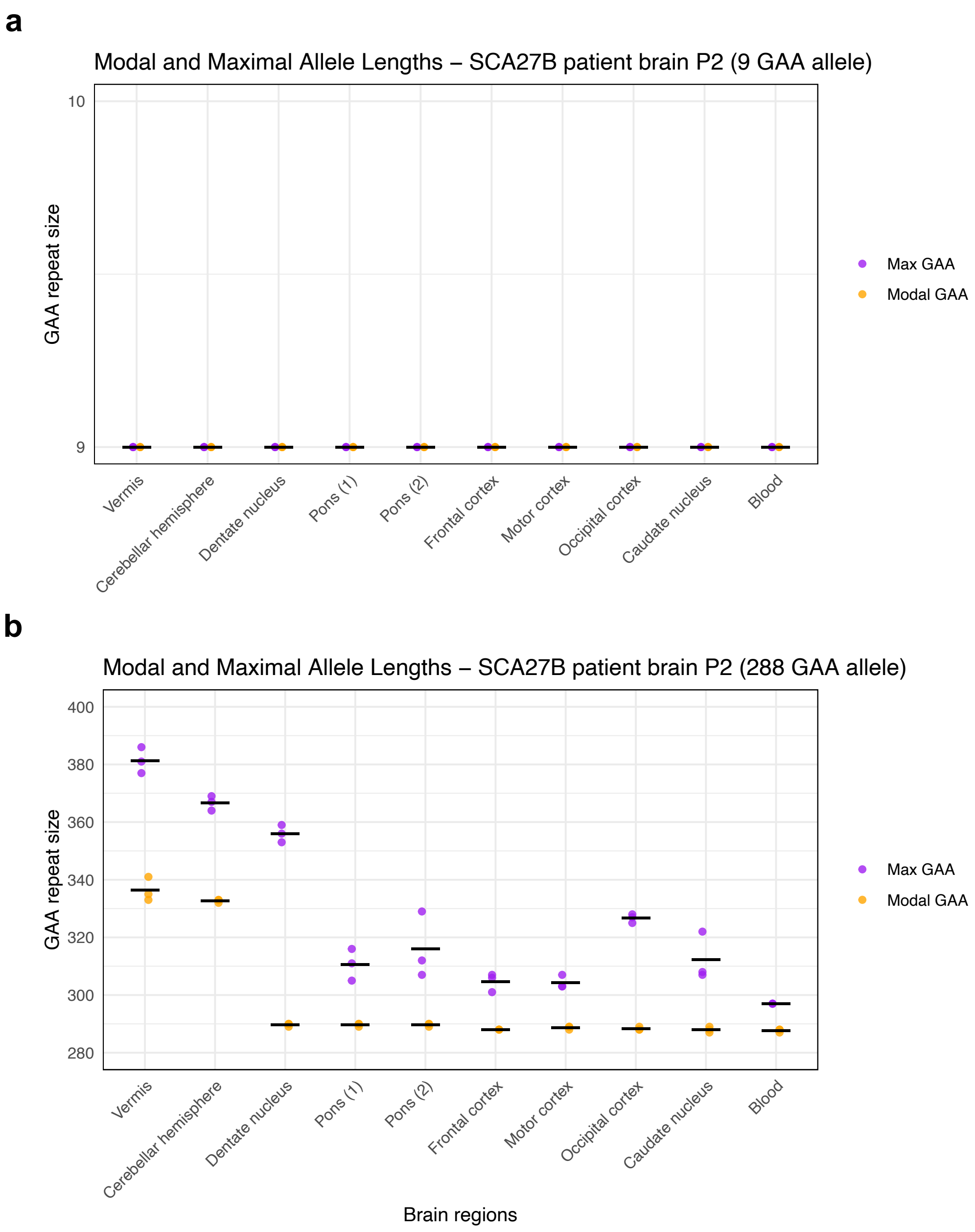


Modal and maximal GAA•TTC repeat sizes (using a 1% relative peak threshold) measured in triplicate by capillary electrophoresis of long-range PCR amplification products in post-mortem central nervous system regions from SCA27B patient brain P2. Results for the short and long allele are shown in panels a and b, respectively. Modal sizes are represented by orange points, while maximum sizes are represented by purple points. Individual data points correspond to repeated measurements. Horizontal black lines show mean of replicates.

**Supplementary Figure 25: Modal and maximal *FGF14* GAA•TTC repeat length in post-mortem central nervous system tissues from SCA27B patient brain P3**

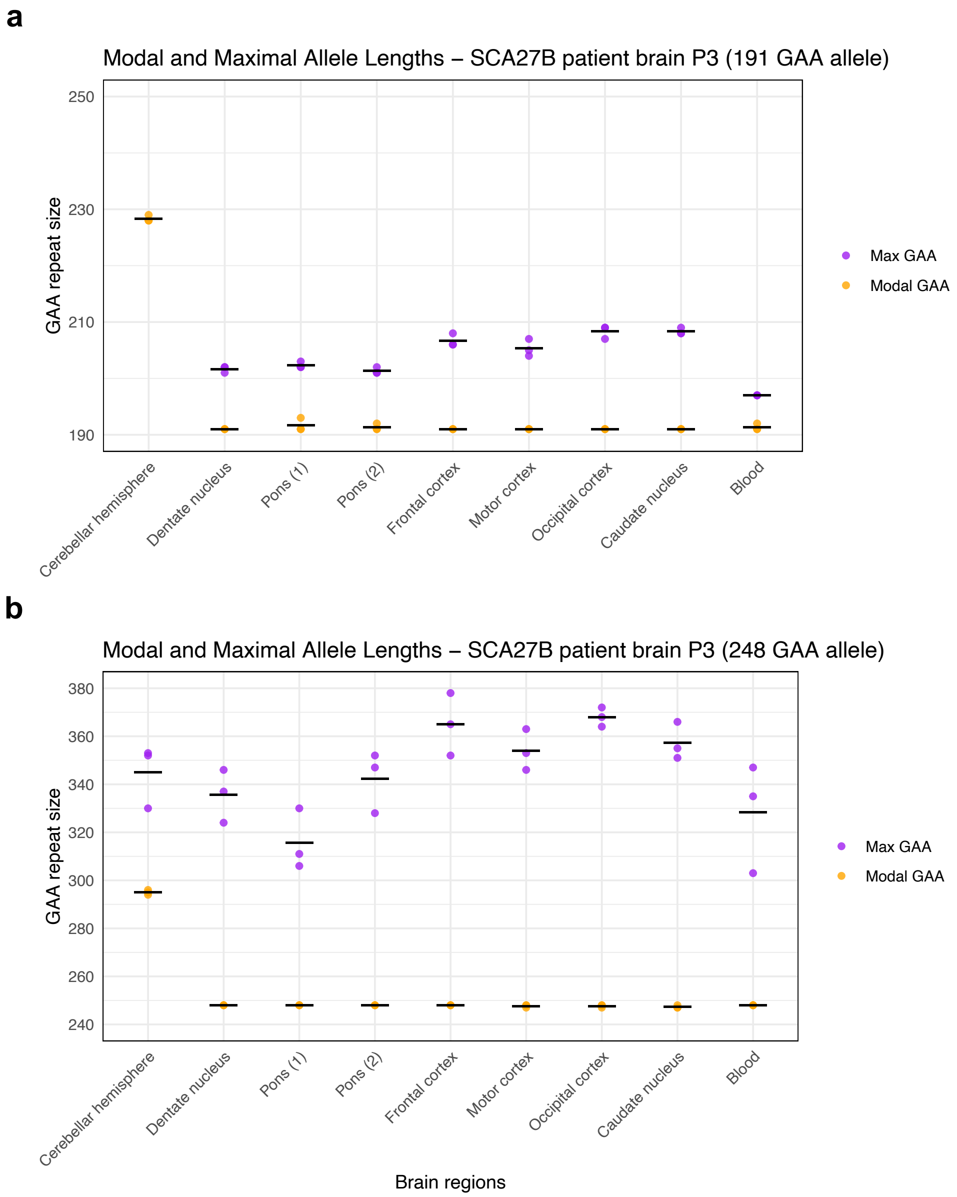


Modal and maximal GAA•TTC repeat sizes (using a 1% relative peak threshold) measured in triplicate by capillary electrophoresis of long-range PCR amplification products in post-mortem central nervous system regions from SCA27B patient brain P3. Results for the short and long allele are shown in panels a and b, respectively. Maximum size of the short allele could not be calculated in the cerebellar hemispheres due to the extreme instability of this tissue, which prevented the distinction of the short and long allele beyond their modal peak. Modal sizes are represented by orange points, while maximum sizes are represented by purple points. Individual data points correspond to repeated measurements. Horizontal black lines show mean of replicates.

**Supplementary Figure 26: Correlation of GAA•TTC repeat length with expansion index across multiple central nervous system regions**

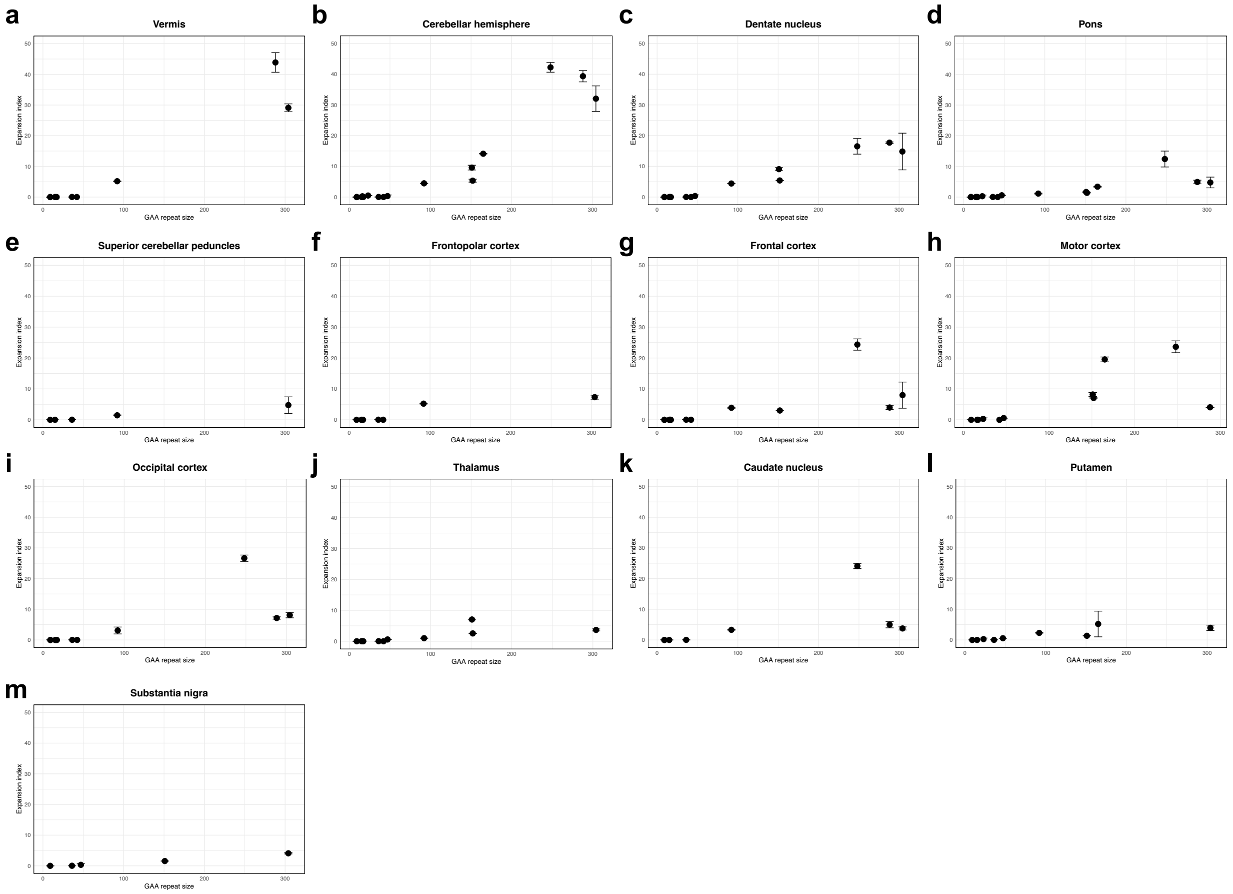


Correlation between *FGF14* GAA•TTC repeat length and expansion index across multiple central nervous system regions. Points show the mean expansion index values measured across replicate PCR reactions and error bars the standard deviation. For regions where multiple tissue sub-pieces were analyzed, the mean expansion index across all sub-pieces is shown. Regression analysis and Pearson's correlation showed a significant linear relationship between repeat length and expansion index in (a) the vermis (r = 0.96 [95% CI: 0.81 to 0.99], slope = 0.130, $p$<0.0001), (b) the cerebellar hemispheres (r = 0.93 [95% CI: 0.81 to 0.98], slope = 0.133, $p$<0.0001), (c) the dentate nucleus (r = 0.98 [95% CI: 0.92 to 0.99], slope = 0.062, $p$<0.0001), (d) the pons (r = 0.79 [95% CI: 0.48 to 0.92], slope = 0.025, $p$=0.00029), (e) the superior cerebellar peduncles (r = 1.00 [95% CI: 0.93 to 1.00], slope = 0.017, $p$=0.00033), (f) the frontopolar cortex (r = 0.90 [95% CI: 0.47 to 0.99], slope = 0.027, $p$=0.0052), (g) the frontal cortex (r = 0.67 [95% CI: 0.16 to 0.90], slope = 0.041, $p$=0.016), (h) the motor cortex (r = 0.70 [95% CI: 0.17 to 0.92], slope = 0.059, $p$=0.016), (i) the occipital cortex (r = 0.71 [95% CI: 0.15 to 0.93], slope = 0.049, $p$=0.020), (j) the thalamus (r = 0.73 [95% CI: 0.23 to 0.93], slope = 0.018, $p$=0.011), (l) the putamen (r = 0.82 [95% CI: 0.34 to 0.96], slope = 0.016, $p$=0.0068), and (m) the substantia nigra (r = 0.99 [95% CI: 0.91 to 1.00], slope = 0.014, $p$=0.00051), but not in (k) the caudate nucleus (r = 0.57 [95% CI: -0.22 to 0.91], slope = 0.036, $p$=0.14).

**Supplementary Figure 27: Capillary electrophoresis profiles of the *FGF14* GAA•TTC repeat in post-mortem central nervous system tissues from control brain C1**

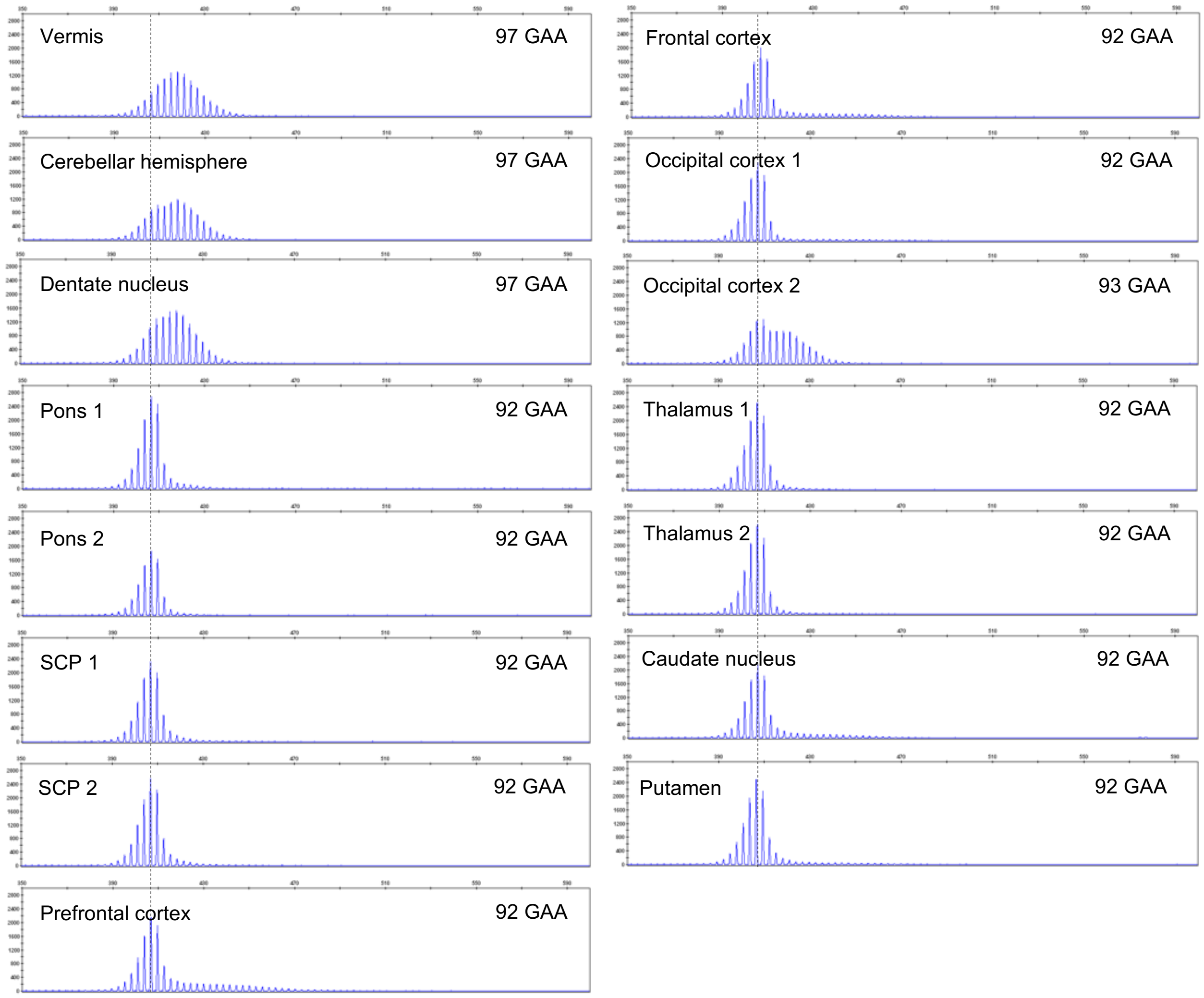


Representative capillary electrophoresis traces (visualized with the GeneMapper software) of the longer *FGF14* $(GAA)_{92}$ allele in post-mortem brain regions from control brain C1. The vertical dashed black lines indicate the modal GAA•TTC allele of 92 repeat units measured in all non-cerebellar brain regions. The small allele of 15 GAA•TTC repeat units is not shown due to the absence of significant instability of this allele. SCP: superior cerebellar peduncles.

**Supplementary Figure 28: Capillary electrophoresis profiles of the *FGF14* GAA•TTC repeat in post-mortem central nervous system tissues from control brain C2**

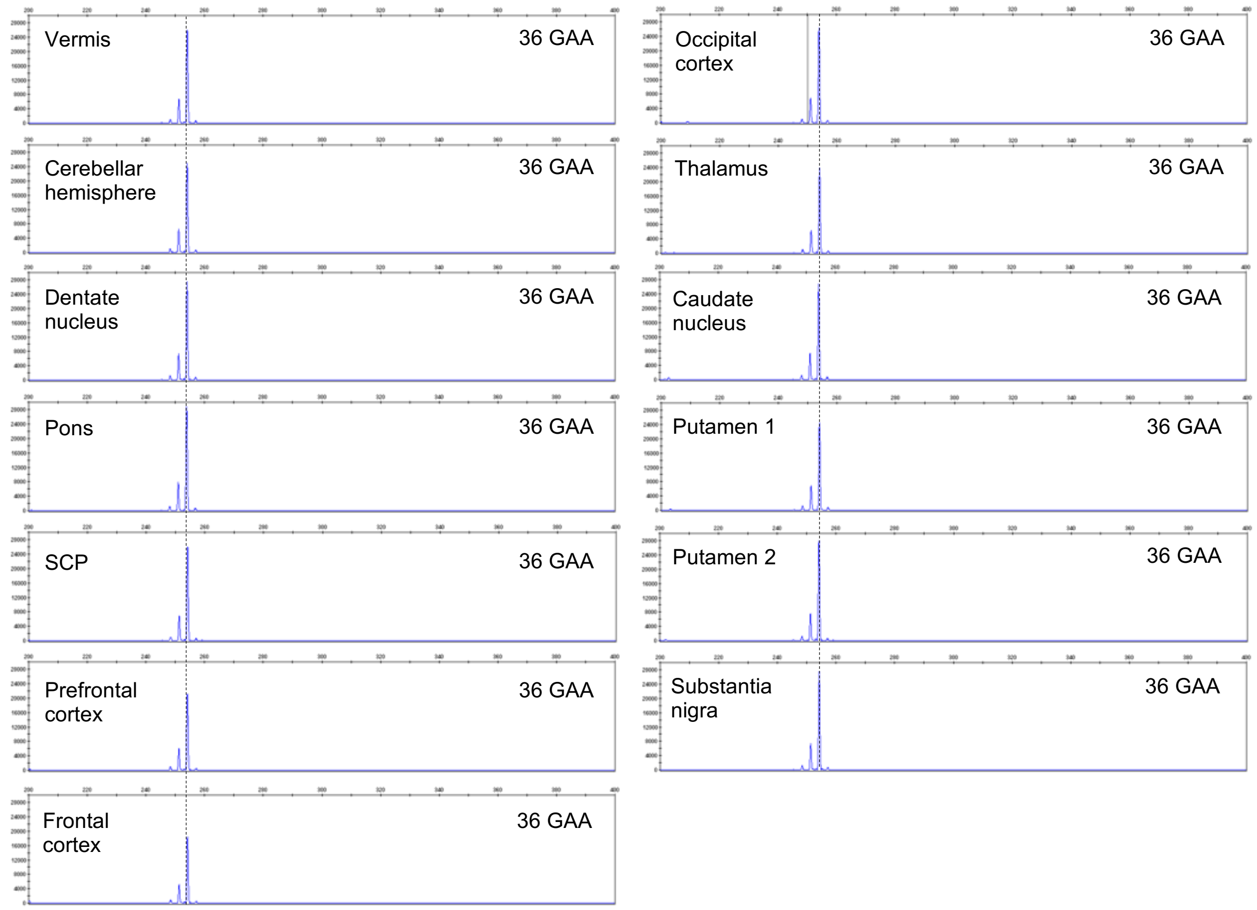


Representative capillary electrophoresis traces (visualized with the GeneMapper software) of the longer *FGF14* $(GAA)_{36}$ allele in post-mortem brain regions from control brain C2. The vertical dashed black lines indicate the modal GAA•TTC allele of 36 repeat units measured in all non-cerebellar brain regions. The small allele of 9 GAA•TTC repeat units is not shown due to the absence of significant instability of this allele. SCP: superior cerebellar peduncles.

**Supplementary Figure 29: Capillary electrophoresis profiles of the *FGF14* GAA•TTC repeat in post-mortem central nervous system tissues from control brain C3**

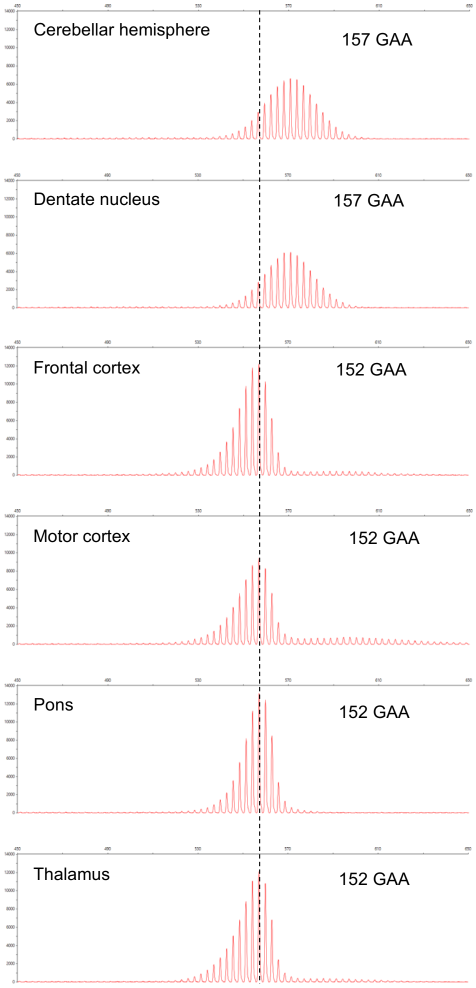


Representative capillary electrophoresis traces (visualized with the Peak Scanner software) of the longer *FGF14* $(GAA)_{152}$ allele in post-mortem brain regions from control brain C3. The vertical dashed black lines indicate the modal GAA•TTC allele of 152 repeat units measured in all non-cerebellar brain regions. The small allele of 16 GAA•TTC repeat units is not shown due to the absence of significant instability of this allele.

**Supplementary Figure 30: Capillary electrophoresis profiles of the *FGF14* GAA•TTC repeat in post-mortem central nervous system tissues from control brain C4**

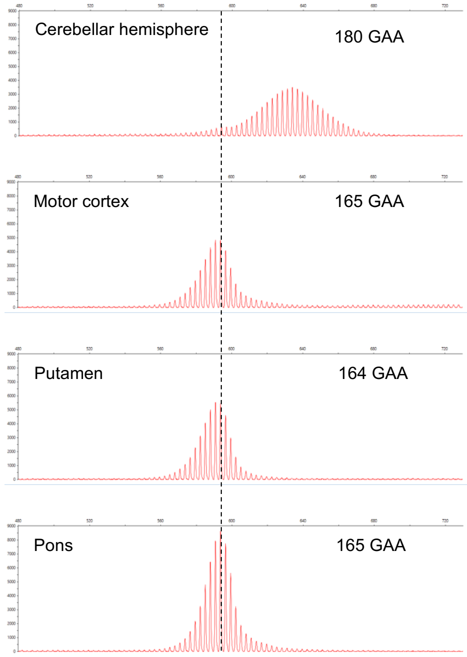


Representative capillary electrophoresis traces (visualized with the Peak Scanner software) of the longer *FGF14* $(GAA)_{165}$ allele in post-mortem brain regions from control brain C4. The vertical dashed black lines indicate the modal GAA•TTC allele of 165 repeat units measured in all non-cerebellar brain regions. The small allele of 23 GAA•TTC repeat units is not shown due to the absence of significant instability of this allele.

**Supplementary Figure 31: Capillary electrophoresis profiles of the *FGF14* GAA•TTC repeat in post-mortem central nervous system tissues from control brain C5**

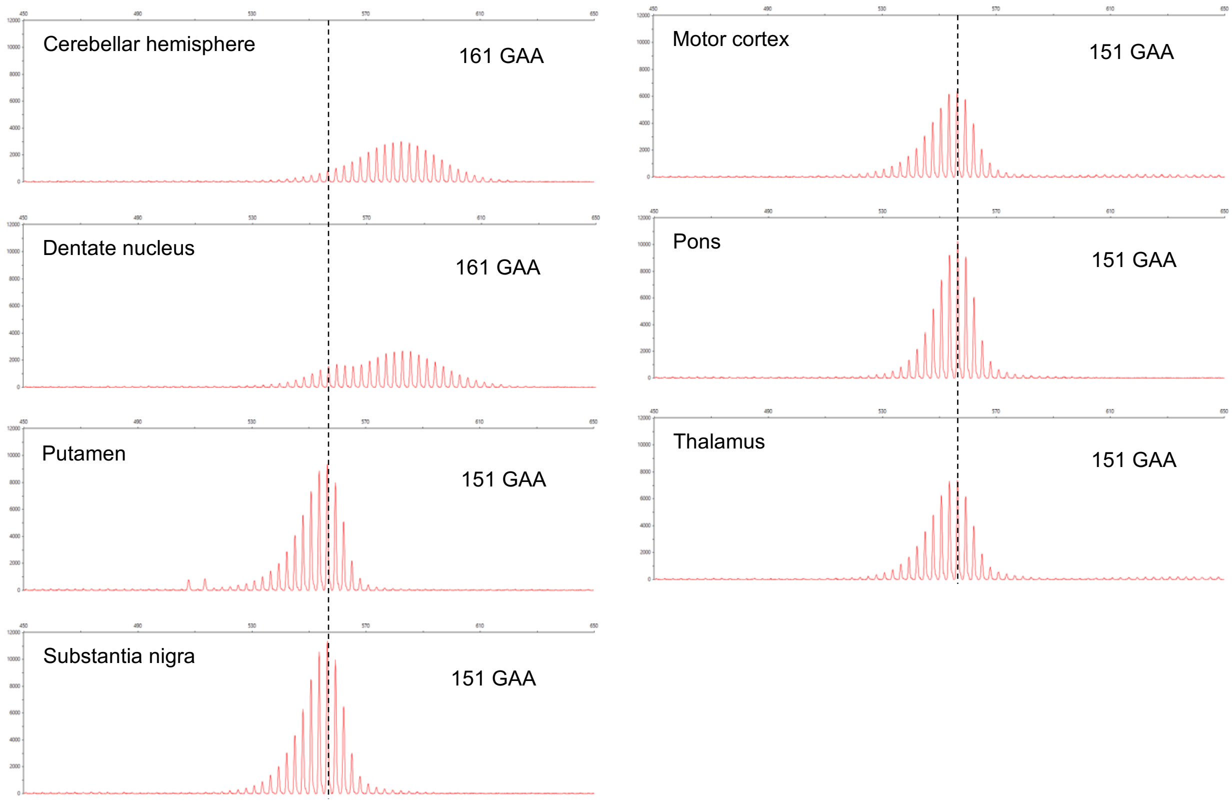


Representative capillary electrophoresis traces (visualized with the Peak Scanner software) of the longer *FGF14* $(GAA)_{151}$ allele in post-mortem brain regions from control brain C5. The vertical dashed black lines indicate the modal GAA•TTC allele of 151 repeat units measured in all non-cerebellar brain regions. The small allele of 47 GAA•TTC repeat units is not shown due to the absence of significant instability of this allele.

**Supplementary Figure 32: Capillary electrophoresis profiles of the *FGF14* GAA•TTC repeat in post-mortem central nervous system tissues from control brain C6**

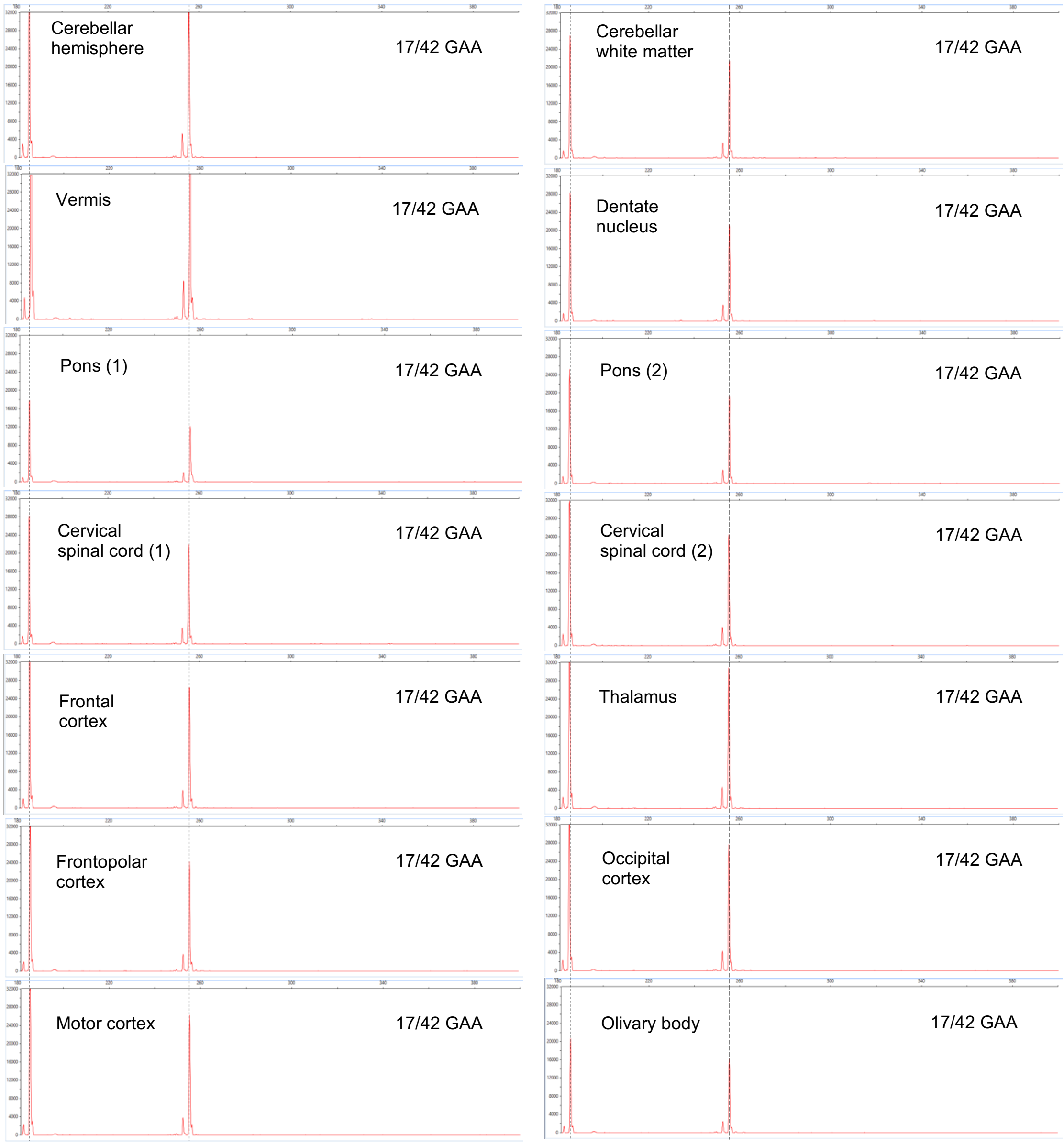


Representative capillary electrophoresis traces (visualized with the Peak Scanner software) of the *FGF14* $(GAA)_{17}$ and $(GAA)_{42}$ alleles in post-mortem brain regions from control brain C6. The vertical dashed black lines indicate the modal GAA•TTC alleles of 17 and 42 repeat units.

**Supplementary Figure 33: Capillary electrophoresis profiles of the *FGF14* GAA•TTC repeat in post-mortem central nervous system tissues from SCA27B patient brain P1**

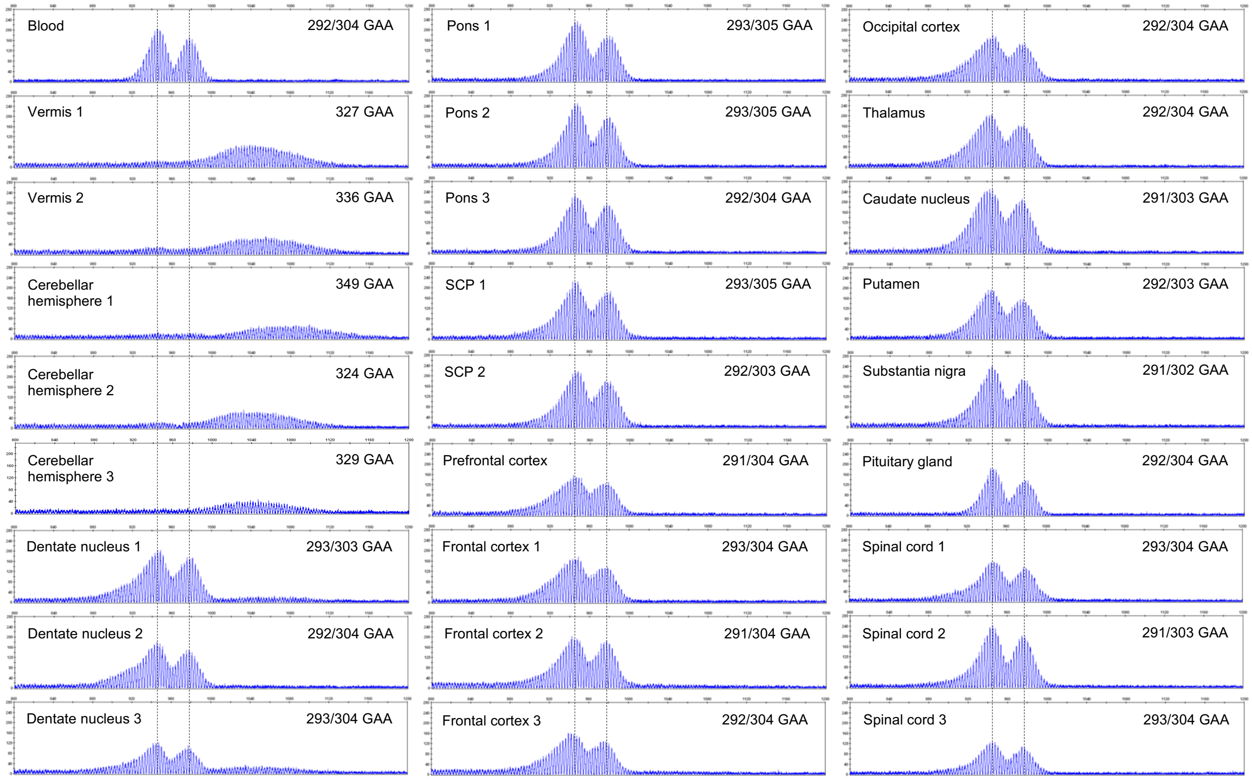


Representative capillary electrophoresis traces (visualized with the GeneMapper software) of the *FGF14* GAA•TTC alleles in post-mortem brain regions from SCA27B patient brain P1. The vertical dashed black lines indicate the modal GAA•TTC alleles of 292 and 304 repeat units measured in the blood. SCP: superior cerebellar peduncles.

**Supplementary Figure 34: Capillary electrophoresis profiles of the *FGF14* GAA•TTC repeat in post-mortem central nervous system tissues from SCA27B patient brain P2**

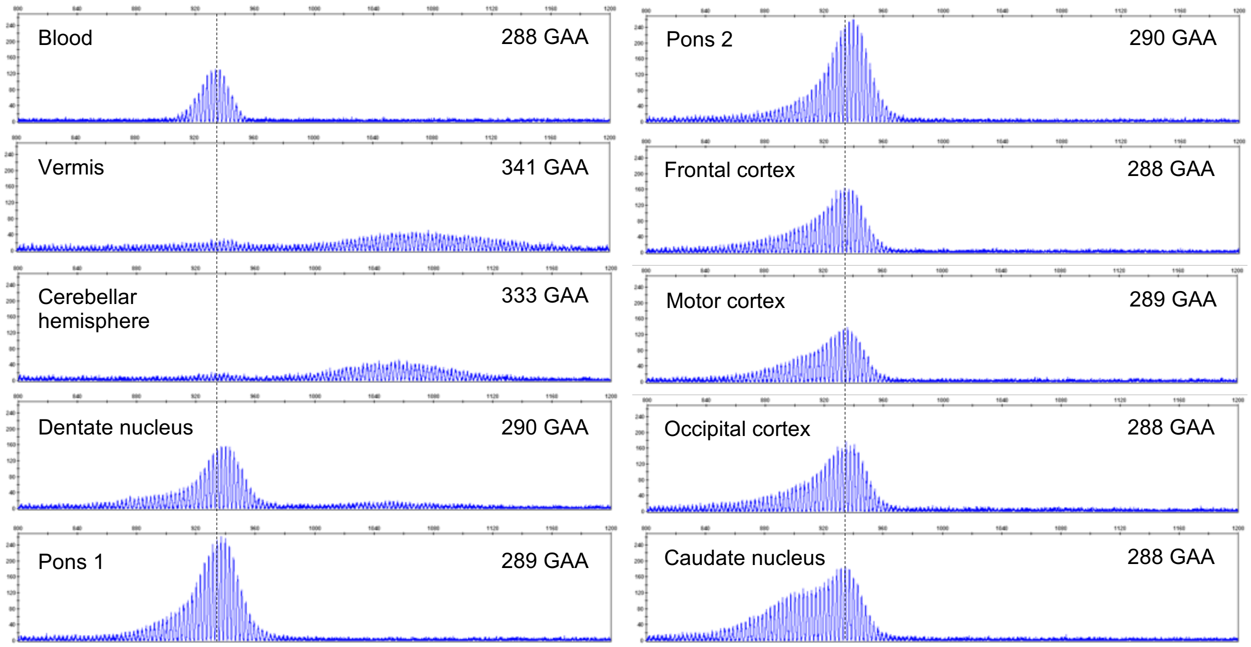


Representative capillary electrophoresis traces (visualized with the GeneMapper software) of the *FGF14* $(GAA)_{288}$ allele in post-mortem brain regions from SCA27B patient brain P2. The vertical dashed black lines indicate the modal GAA•TTC allele of 288 repeat units measured in the blood. The small allele of 9 GAA•TTC repeat units is not shown due to the absence of significant instability of this allele.

**Supplementary Figure 35: Capillary electrophoresis profiles of the *FGF14* GAA•TTC repeat in post-mortem central nervous system tissues from SCA27B patient brain P3**

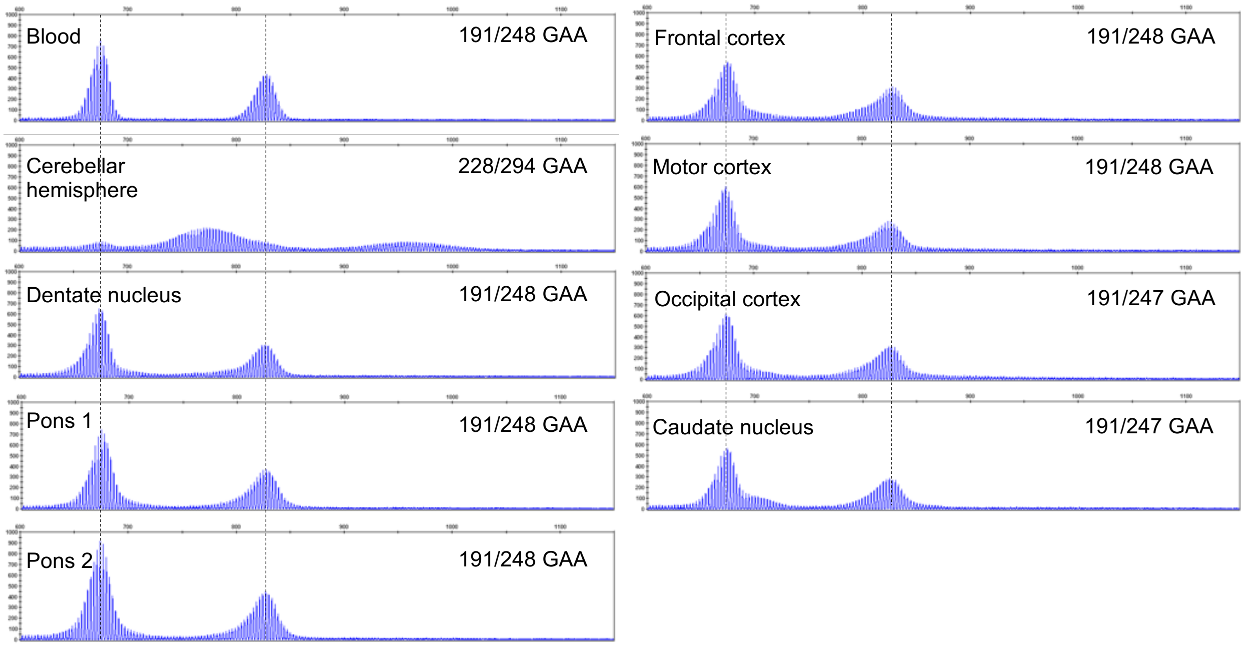


Representative capillary electrophoresis traces (visualized with the GeneMapper software) of the *FGF14* GAA•TTC alleles in post-mortem brain regions from SCA27B patient brain P3. The vertical dashed black lines indicate the modal GAA•TTC alleles of 191 and 248 repeat units measured in the blood.

**Supplementary Figure 36: Technical variation across replicate PCRs from post-mortem central nervous system samples of control brain C1**

**a**

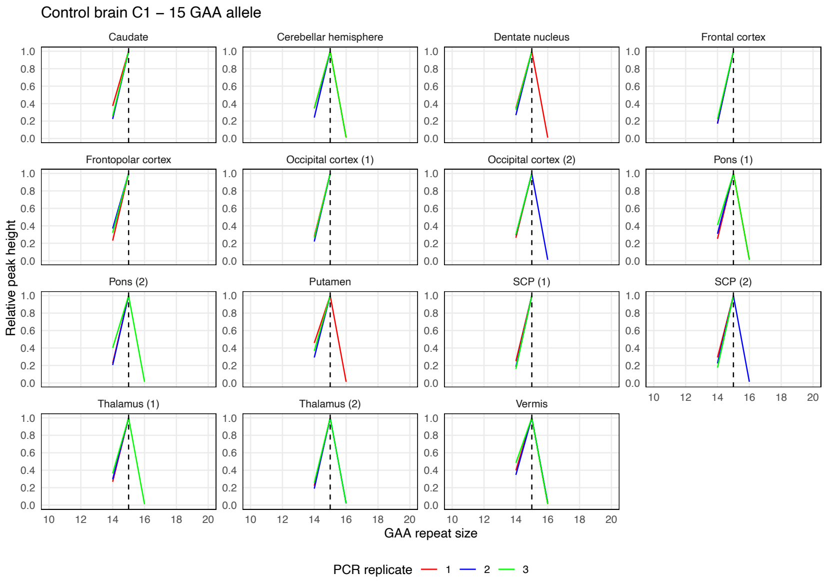


**b**

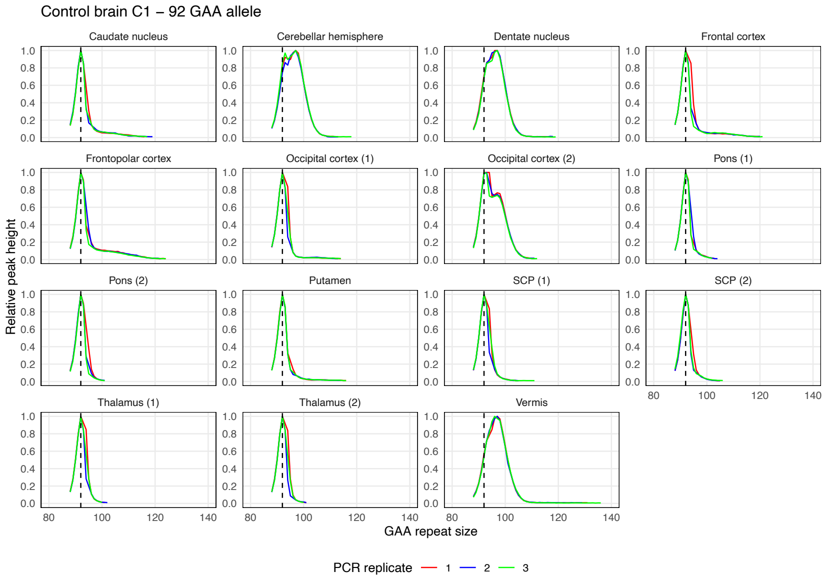


Somatic instability profiles of (a) the *FGF14* $(GAA)_{15}$ repeat allele and (b) the *FGF14* $(GAA)_{92}$ repeat allele in different brain regions, derived from post-mortem samples of control brain C1. Each plot shows the instability profile within a given brain region across triplicate PCR reactions. For regions where multiple tissue sub-pieces were analyzed, results for each sub-piece are shown individually. Profiles were plotted by normalizing individual peak height data (extracted from the GeneMapper software) to the height of the modal allele within each brain region. Peaks left of the modal allele above a 10% threshold and those right of the modal allele above a 1% threshold were plotted. Vertical dashed black lines indicate the size of the modal alleles measured in non-cerebellar regions. SCP: superior cerebellar peduncles.

**Supplementary Figure 37: Technical variation across replicate PCRs from post-mortem central nervous system samples of control brain C2**

**a**

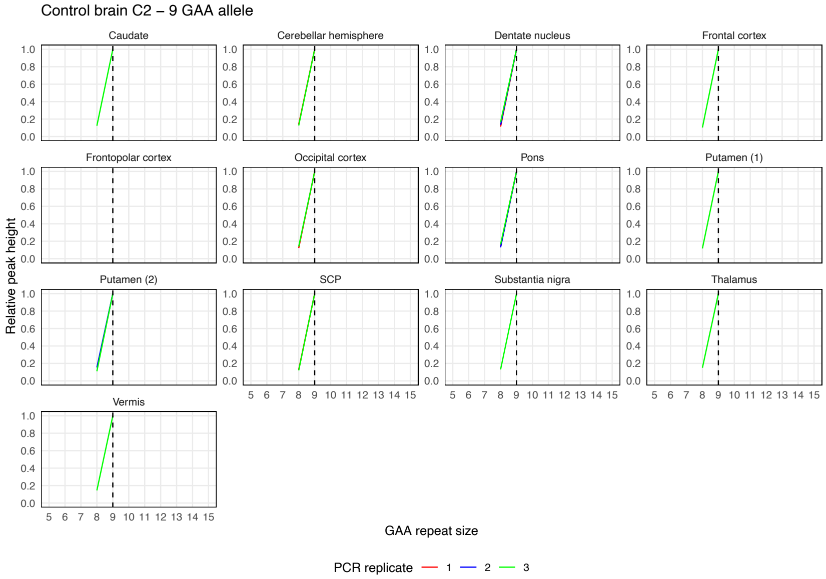


**b**

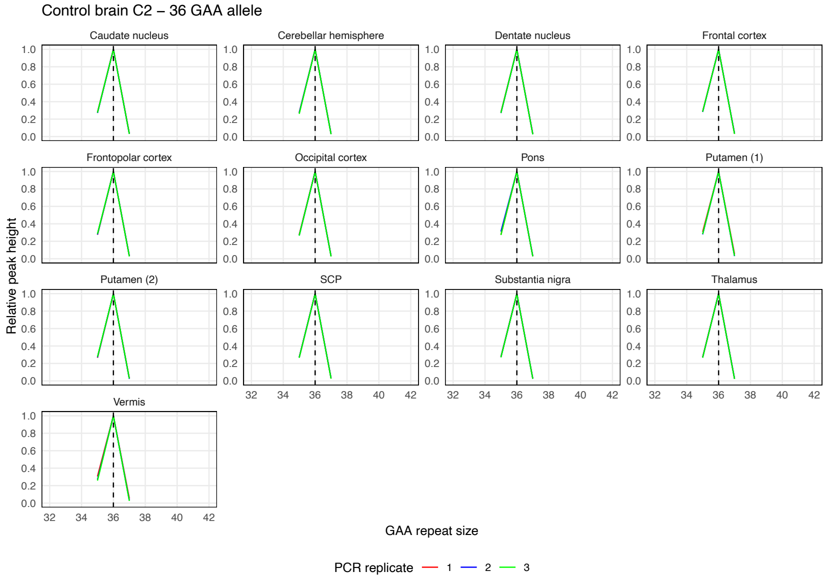


Somatic instability profiles of (a) the *FGF14* $(GAA)_9$ repeat allele and (b) the *FGF14* $(GAA)_{36}$ repeat allele in different brain regions, derived from post-mortem samples of control brain C2. Each plot shows the instability profile within a given brain region across triplicate PCR reactions. For regions where multiple tissue sub-pieces were analyzed, results for each sub-piece are shown individually. Profiles were plotted by normalizing individual peak height data (extracted from the GeneMapper software) to the height of the modal allele within each brain region. Peaks left of the modal allele above a 10% threshold and those right of the modal allele above a 1% threshold were plotted. Vertical dashed black lines indicate the size of the modal alleles measured in non-cerebellar regions. SCP: superior cerebellar peduncles.

**Supplementary Figure 38: Technical variation across replicate PCRs from post-mortem central nervous system samples of control brain C3**

a

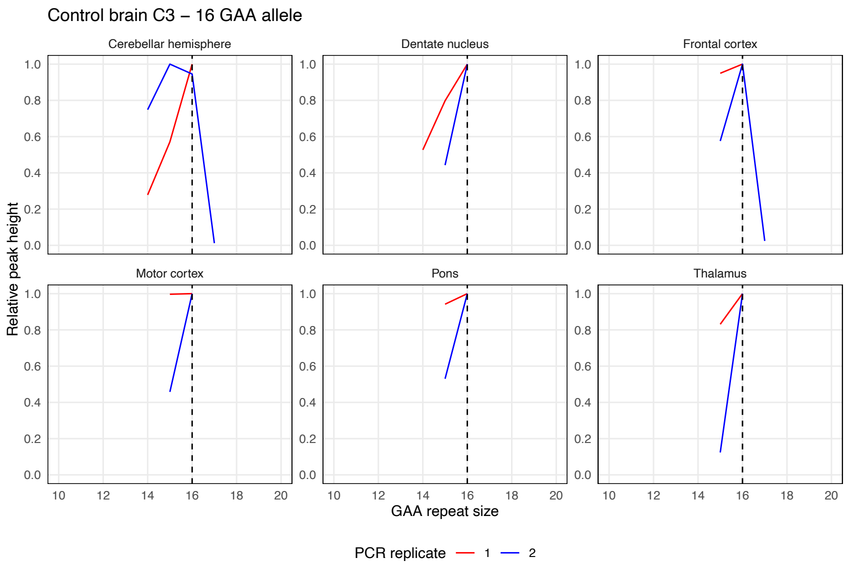


b

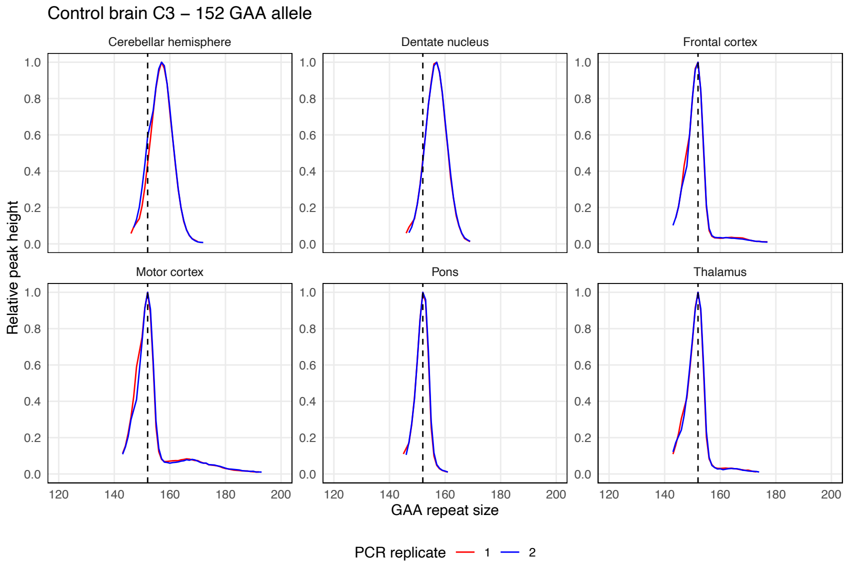


Somatic instability profiles of (a) the *FGF14* $(GAA)_{16}$ repeat allele and (b) the *FGF14* $(GAA)_{152}$ repeat allele in different brain regions, derived from post-mortem samples of control brain C3. Each plot shows the instability profile within a given brain region across duplicate PCR reactions. Profiles were plotted by normalizing individual peak height data (extracted from the Peak Scanner software) to the height of the modal allele within each brain region. Peaks left of the modal allele above a 10% threshold and those right of the modal allele above a 1% threshold were plotted. Vertical dashed black lines indicate the size of the modal alleles measured in non-cerebellar regions.

**Supplementary Figure 39: Technical variation across replicate PCRs from post-mortem central nervous system samples of control brain C4**

**a**

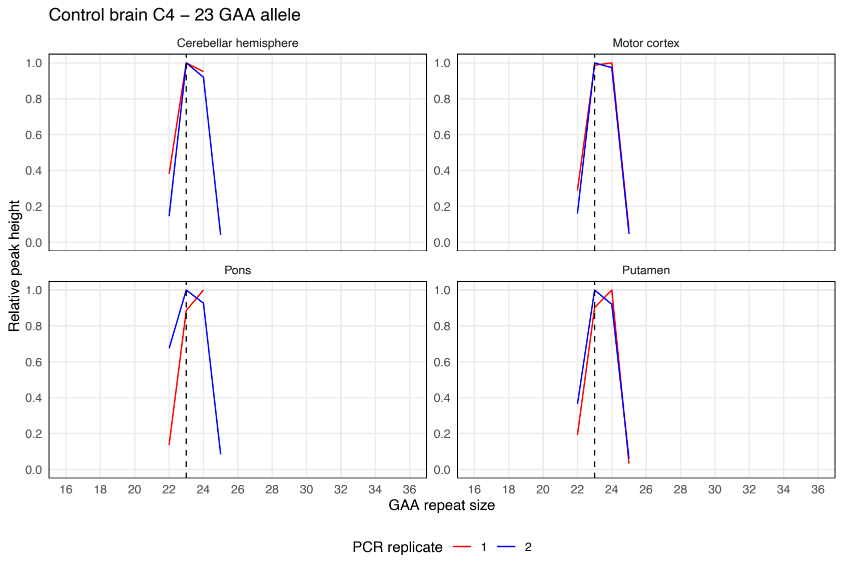


**b**

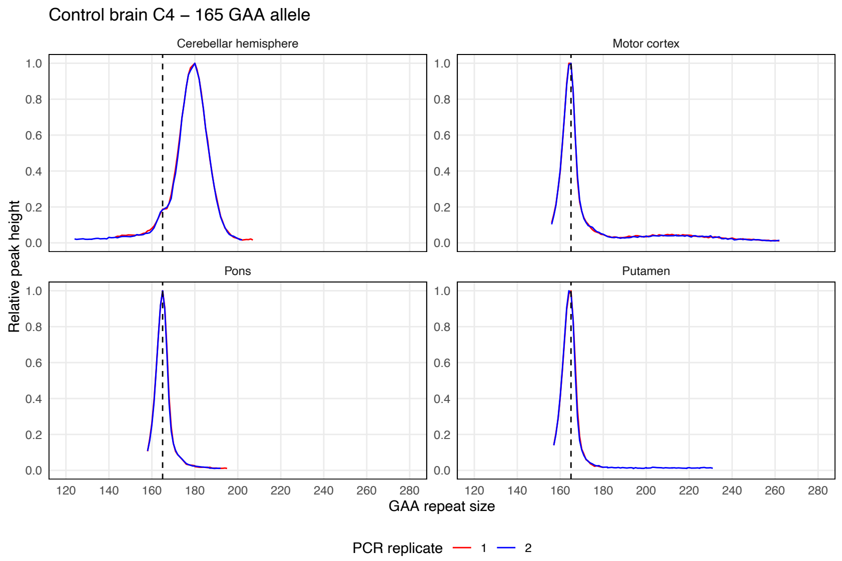


Somatic instability profiles of (a) the *FGF14* $(GAA)_{23}$ repeat allele and (b) the *FGF14* $(GAA)_{165}$ repeat allele in different brain regions, derived from post-mortem samples of control brain C4. Each plot shows the instability profile within a given brain region across duplicate PCR reactions. Profiles were plotted by normalizing individual peak height data (extracted from the Peak Scanner software) to the height of the modal allele within each brain region. Peaks left of the modal allele above a 10% threshold and those right of the modal allele above a 1% threshold were plotted. Vertical dashed black lines indicate the size of the modal alleles measured in non-cerebellar regions.

**Supplementary Figure 40: Technical variation across replicate PCRs from post-mortem central nervous system samples of control brain C5**

a

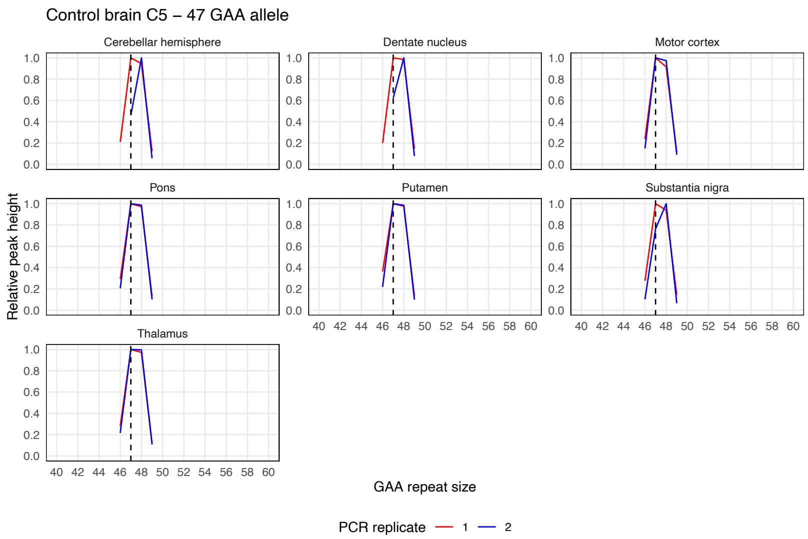


b

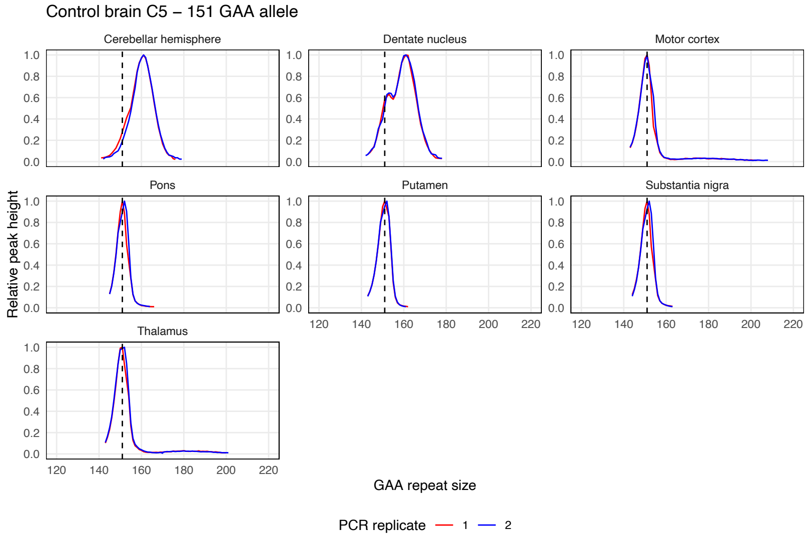


Somatic instability profiles of (a) the *FGF14* $(GAA)_{47}$ repeat allele and (b) the *FGF14* $(GAA)_{151}$ repeat allele in different brain regions, derived from post-mortem samples of control brain C5. Each plot shows the instability profile within a given brain region across duplicate PCR reactions. Profiles were plotted by normalizing individual peak height data (extracted from the Peak Scanner software) to the height of the modal allele within each brain region. Peaks left of the modal allele above a 10% threshold and those right of the modal allele above a 1% threshold were plotted. Vertical dashed black lines indicate the size of the modal alleles measured in non-cerebellar regions.

**Supplementary Figure 41: Technical variation across replicate PCRs from post-mortem central nervous system samples of control brain C6**

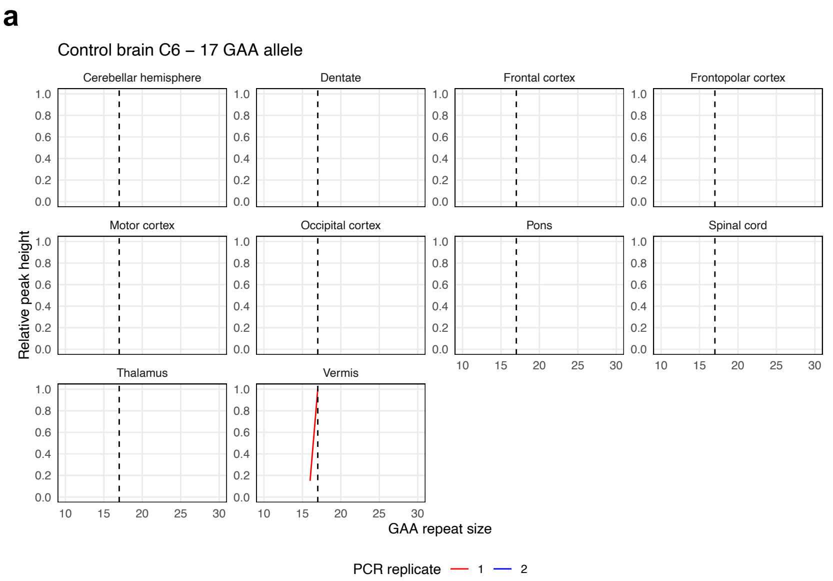


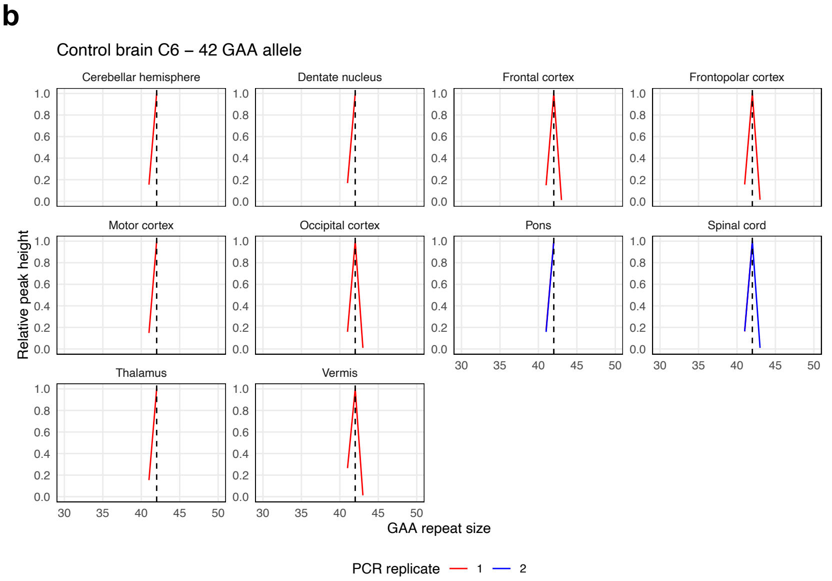


Somatic instability profiles of (a) the *FGF14* $(GAA)_{17}$ repeat allele and (b) the *FGF14* $(GAA)_{42}$ repeat allele in different brain regions, derived from post-mortem samples of control brain C6. Each plot shows the instability profile within a given brain region across replicate PCR reactions. For regions where multiple tissue sub-pieces were analyzed, results for each sub-piece are shown individually. Profiles were plotted by normalizing individual peak height data (extracted from the Peak Scanner software) to the height of the modal allele within each brain region. Peaks left of the modal allele above a 10% threshold and those right of the modal allele above a 1% threshold were plotted. Vertical dashed black lines indicate the size of the modal alleles measured in non-cerebellar regions.

**Supplementary Figure 42: Technical variation across replicate PCRs from post-mortem central nervous system samples of SCA27B patient brain P1**

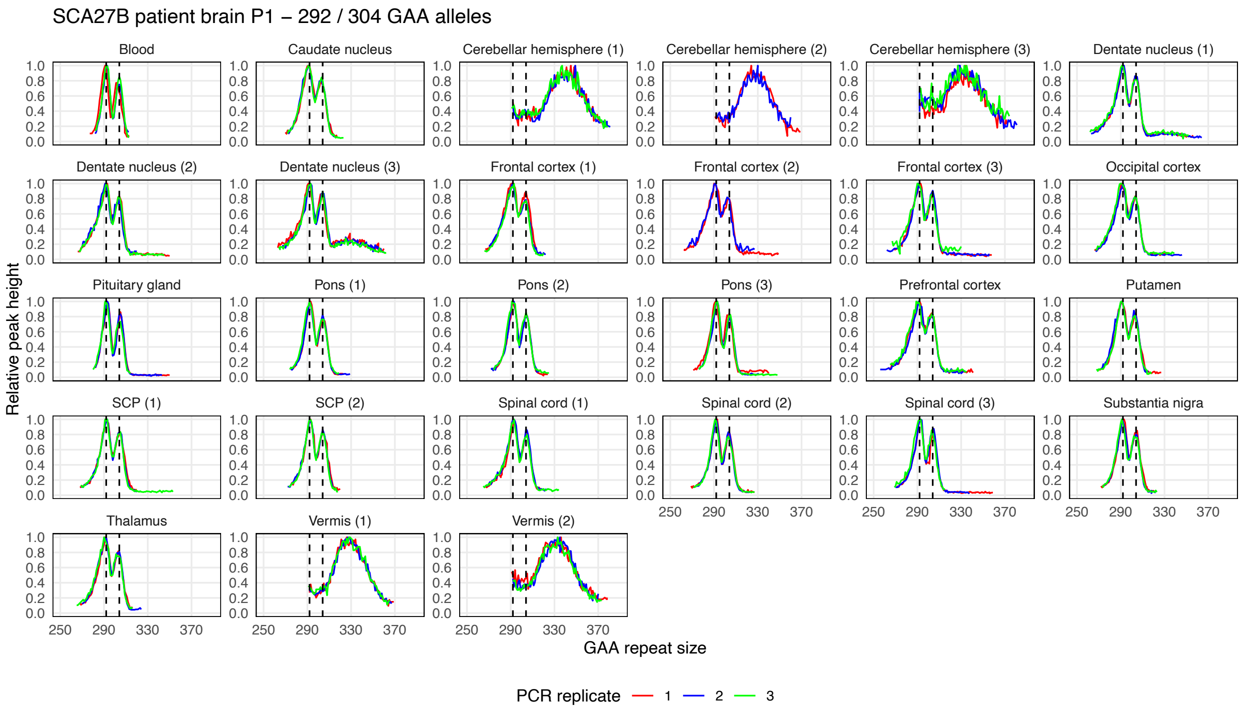


Somatic instability profiles of the *FGF14* $(GAA)_{292}$ and $(GAA)_{304}$ repeat alleles in different brain regions, derived from post-mortem samples of SCA27B patient brain P1. Each plot shows the instability profile within a given brain region across triplicate PCR reactions. For regions where multiple tissue sub-pieces were analyzed, results for each sub-piece are shown individually. Profiles were plotted by normalizing individual peak height data (extracted from the GeneMapper software) to the height of the modal peak of the shorter allele within each brain region. Peaks left of the modal allele above a 10% threshold and those right of the modal allele above a 1% threshold were plotted. Vertical dashed black lines indicate the size of the modal alleles measured in the blood. SCP: superior cerebellar peduncles.

**Supplementary Figure 43: Technical variation across replicate PCRs from post-mortem central nervous system samples of SCA27B patient brain P2**

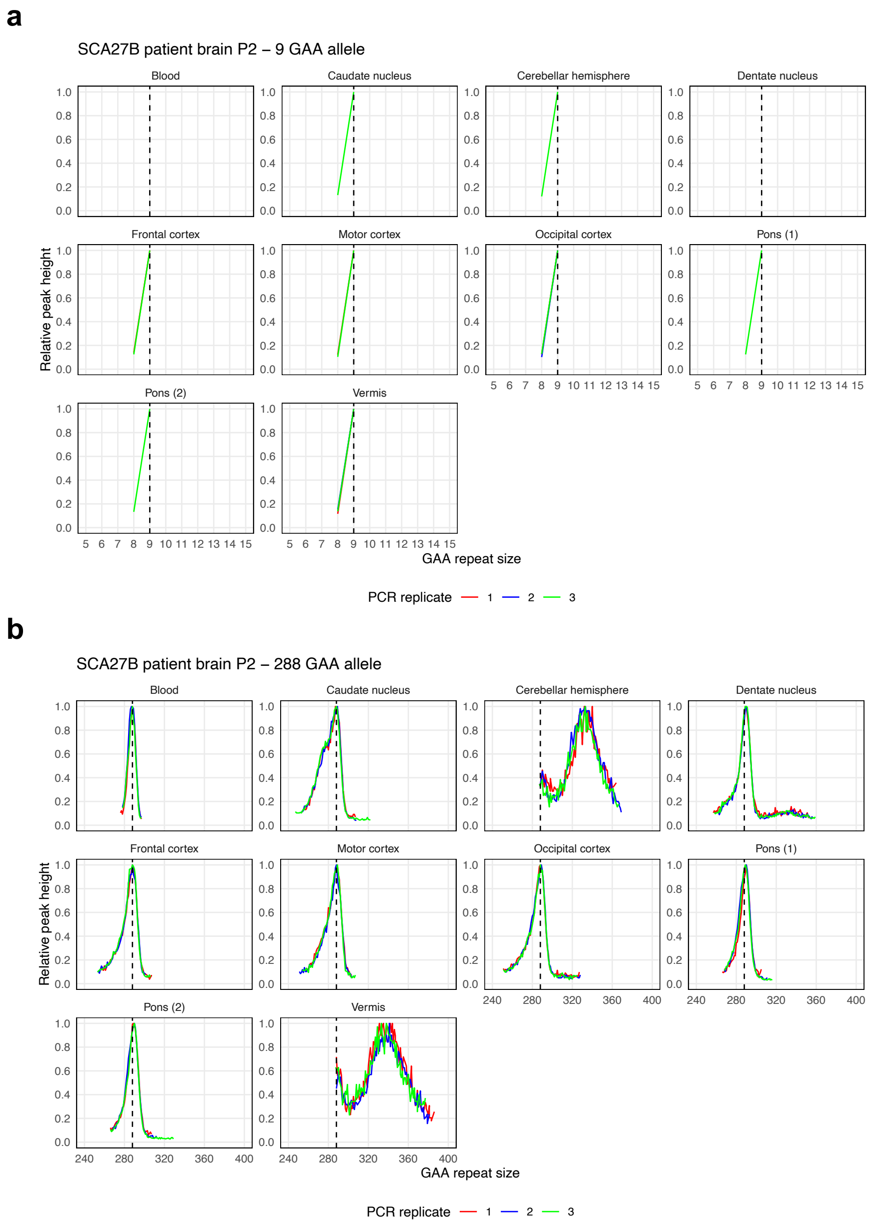


Somatic instability profiles of (a) the *FGF14* $(GAA)_9$ repeat allele and (b) the *FGF14* $(GAA)_{288}$ repeat allele in different brain regions, derived from post-mortem samples of SCA27B patient brain P2. Each plot shows the instability profile within a given brain region across triplicate PCR reactions. For regions where multiple tissue sub-pieces were analyzed, results for each sub-piece are shown individually. Profiles were plotted by normalizing individual peak height data (extracted from the GeneMapper software) to the height of the modal allele within each brain region. Peaks left of the modal allele above a 10% threshold and those right of the modal allele above a 1% threshold were plotted. Vertical dashed black lines indicate the size of the modal alleles measured in the blood.

**Supplementary Figure 44: Technical variation across replicate PCRs from post-mortem central nervous system samples of SCA27B patient brain P3**

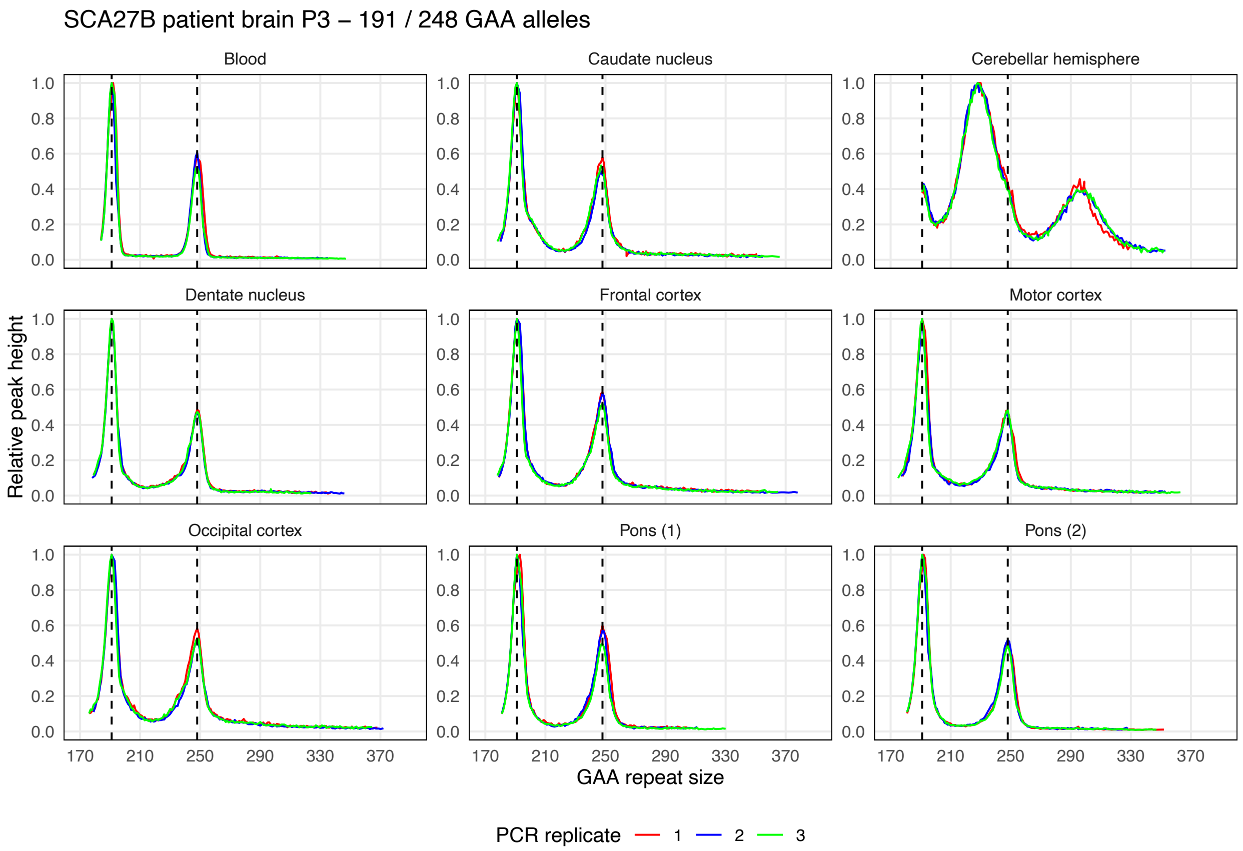


Somatic instability profiles of the *FGF14* $(GAA)_{191}$ and $(GAA)_{248}$ repeat alleles in different brain regions, derived from post-mortem samples of SCA27B patient brain P3. Each plot shows the instability profile within a given brain region across triplicate PCR reactions. For regions where multiple tissue sub-pieces were analyzed, results for each sub-piece are shown individually. Profiles were plotted by normalizing individual peak height data (extracted from the GeneMapper software) to the height of the modal peak of the shorter allele within each brain region. Peaks left of the modal allele above a 10% threshold and those right of the modal allele above a 1% threshold were plotted. Vertical dashed black lines indicate the size of the modal alleles measured in the blood.

**Supplementary Figure 45: Expansion index of individual *FGF14* allele across central nervous system regions**

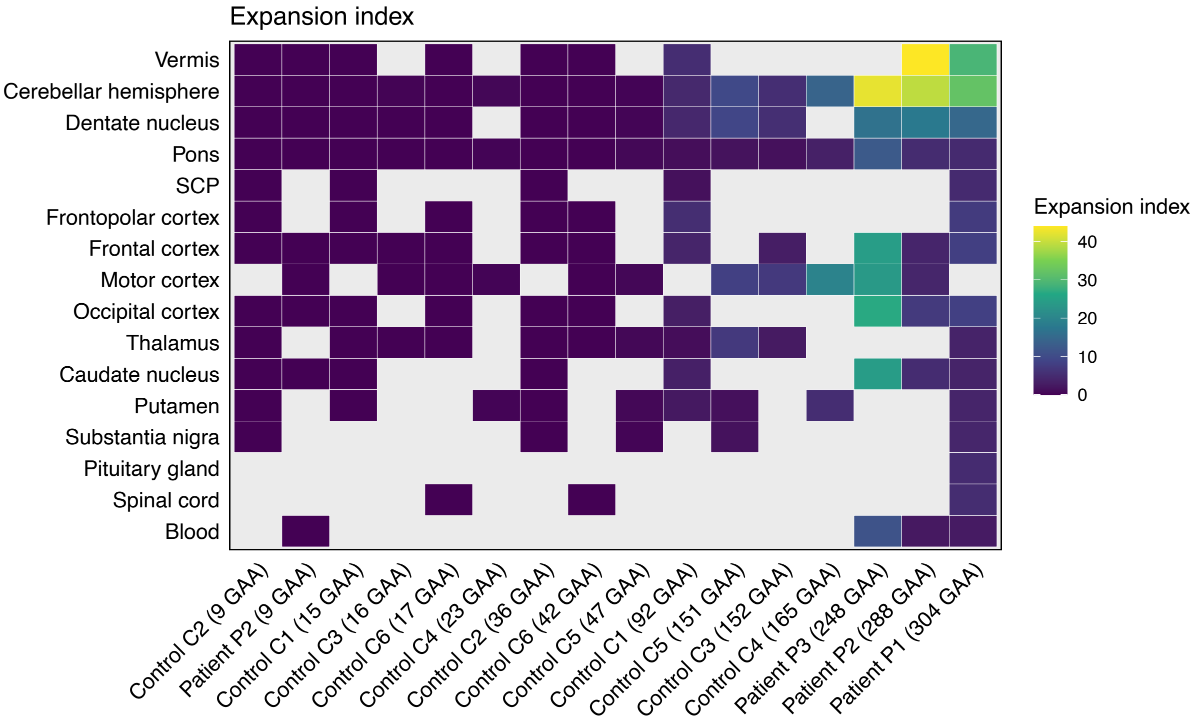


Heatmap of mean expansion indices for individual alleles across central nervous system regions and blood samples. Alleles are arranged in ascending order of length. The color of each tile represents the absolute mean expansion index, according to the color scale shown on the right side of the heatmap. Gray tiles indicate unavailable tissue samples for analysis. SCP: superior cerebellar peduncles.

**Supplementary Figure 46: Ranking of expansion indices in central nervous system regions**

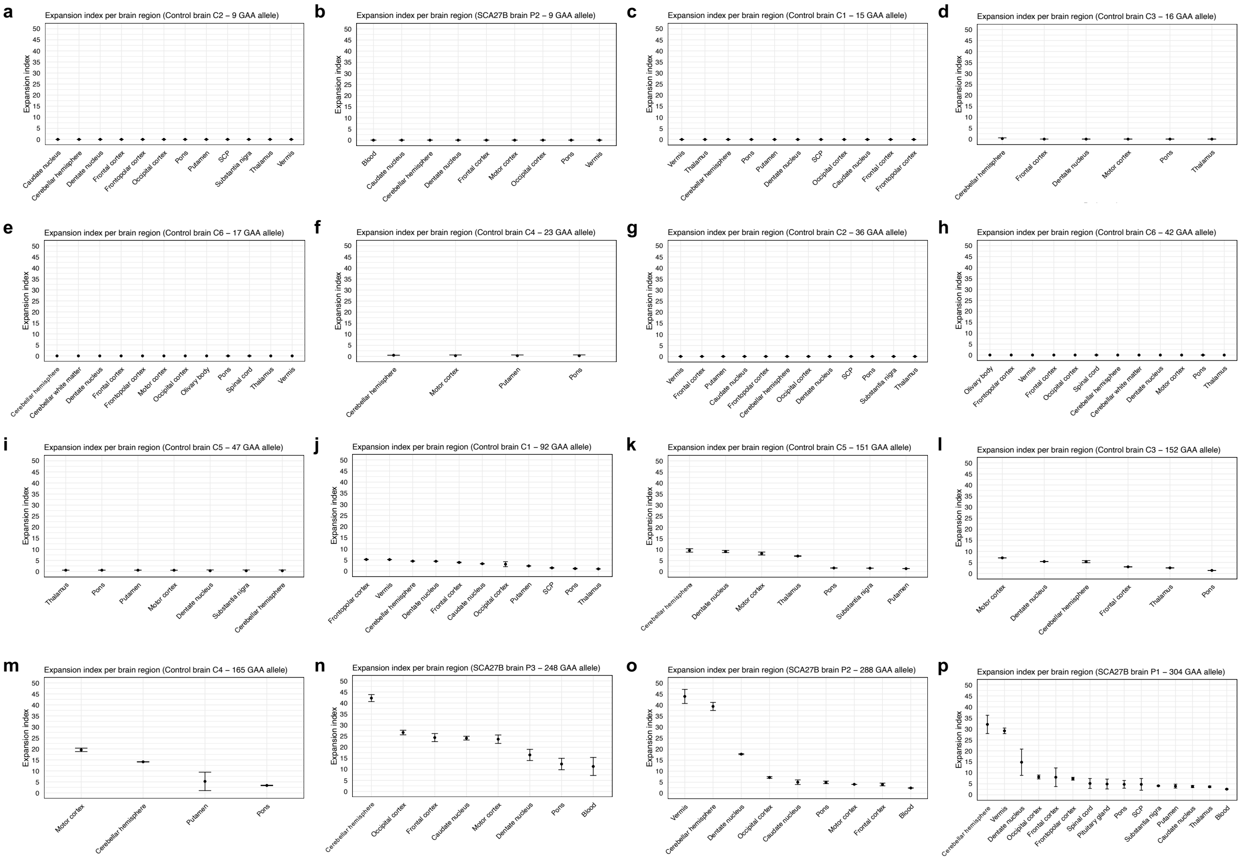


Mean expansion indices for individual alleles across central nervous system regions and blood samples. In each plot, tissues are displayed from left to right in descending order of expansion index value. Points show the mean expansion index values measured across replicate PCR reactions and error bars the standard deviation. For regions where multiple tissue sub-pieces were analyzed, the mean expansion index across all sub-pieces is shown. Results for the short alleles of patients P1 and P3 are not shown as expansion indices could not be calculated. (a, b) Somatic expansion was not detected for the short $(GAA)_9$ alleles (expansion index = 0) in any of the tissues. SCP: superior cerebellar peduncles.

**Supplementary Figure 47: Instability index of individual *FGF14* allele across central nervous system regions**

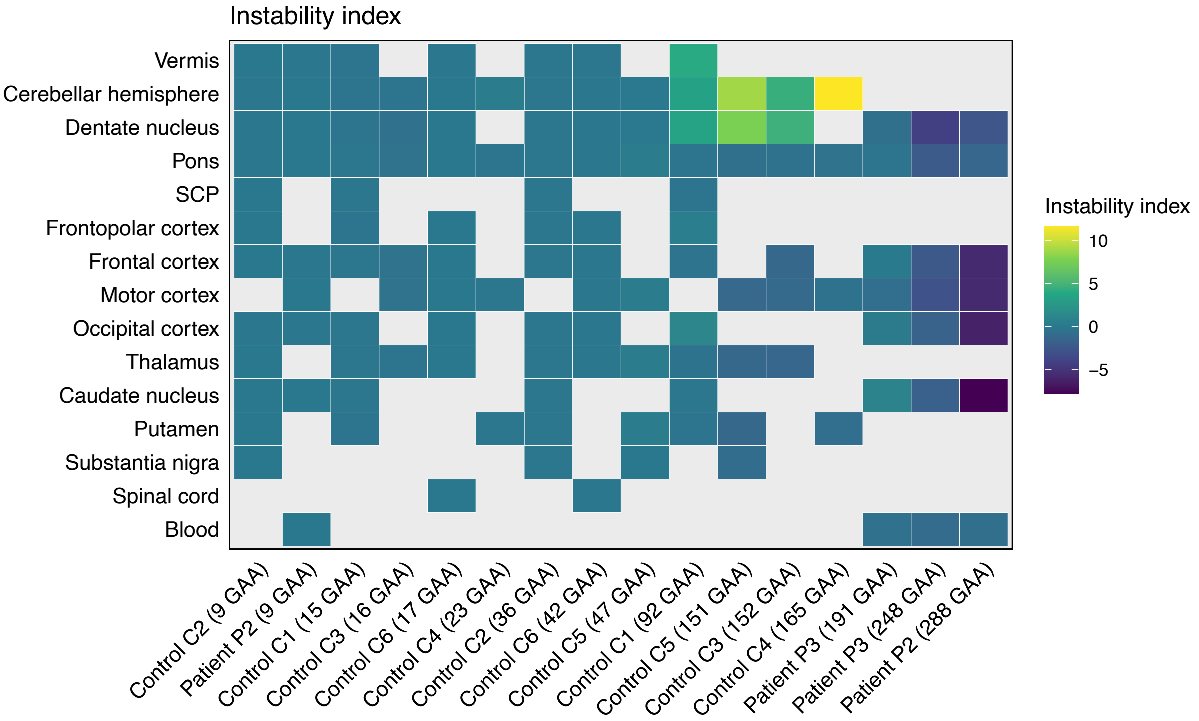


Heatmap of mean instability indices for individual alleles across central nervous system regions and blood samples. Alleles are arranged in ascending order of length. The color of each tile represents the mean instability index, according to the color scale shown on the right side of the heatmap. Gray tiles indicate either unavailable tissue samples for analysis or that instability index could not be calculated. SCP: superior cerebellar peduncles.

**Supplementary Figure 48: Ranking of instability indices in central nervous system regions**

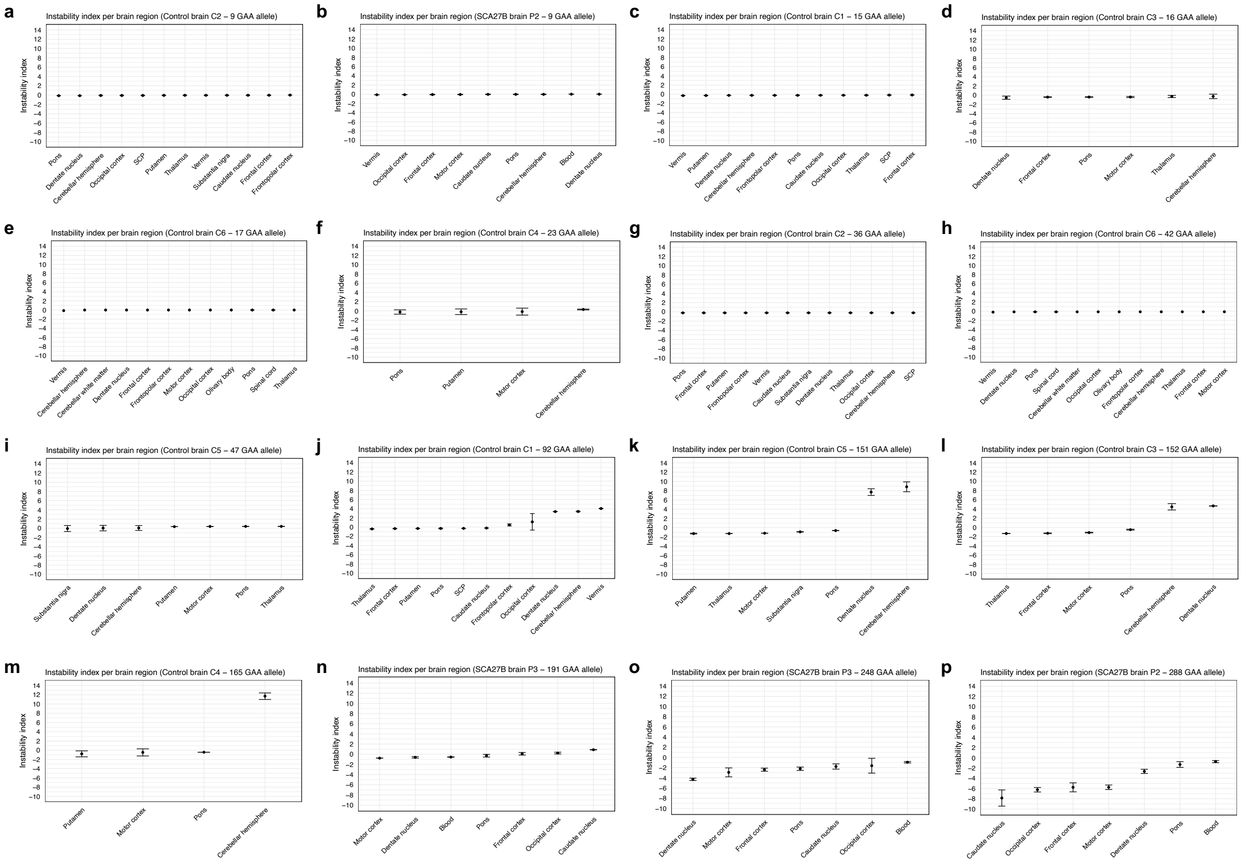


Mean instability indices for individual alleles across central nervous system regions and blood samples. In each plot, tissues are displayed from left to right in ascending order of instability index value. Points show the mean instability index values measured across replicate PCR reactions and error bars the standard deviation. For regions where multiple tissue sub-pieces were analyzed, the mean instability index across all sub-pieces is shown. Results for both alleles of patient P1 are not shown as instability indices could not be calculated. SCP: superior cerebellar peduncles.

**Supplementary Figure 49: Association between GAA•TTC repeat length and instability index across multiple central nervous system regions**

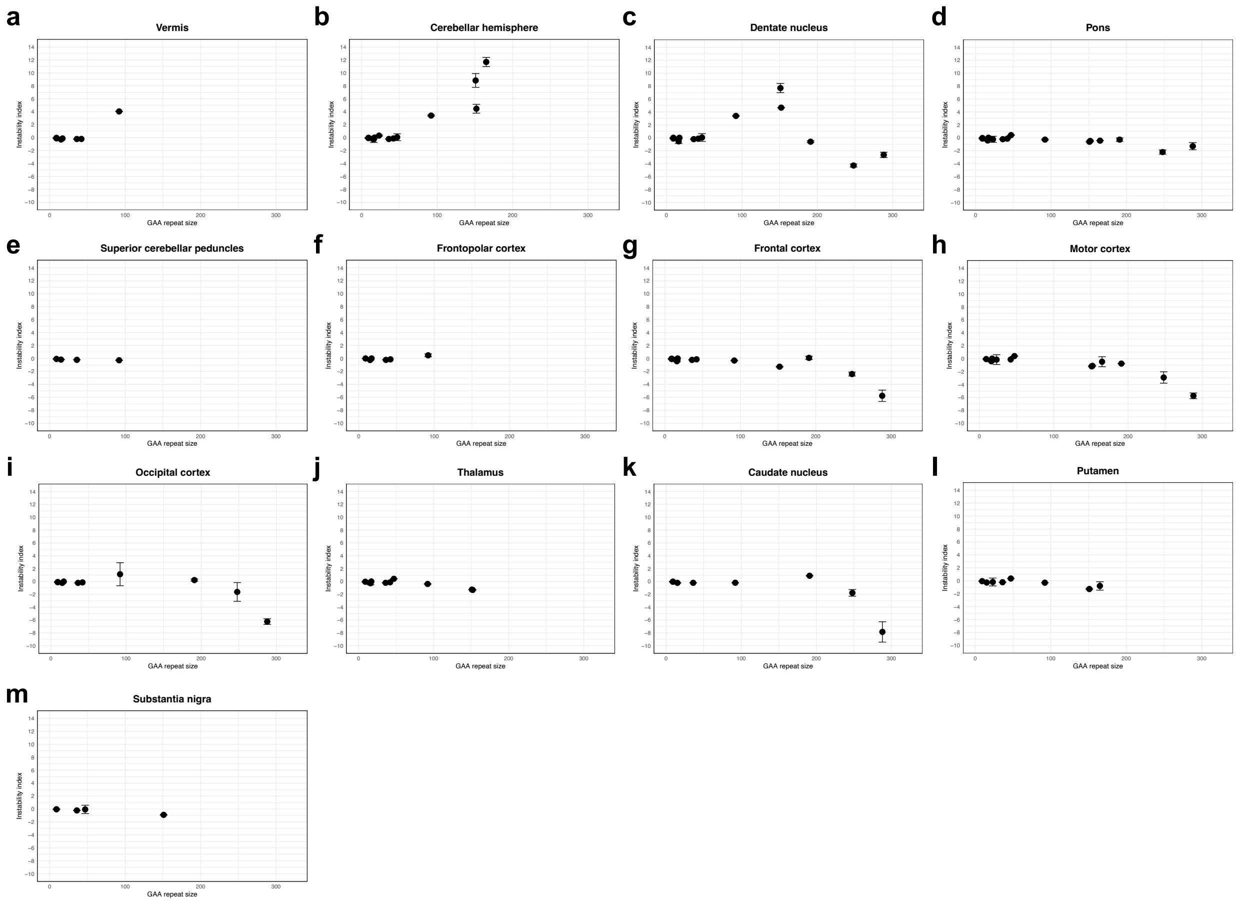


Association between *FGF14* GAA•TTC repeat length and instability index across multiple central nervous system regions. Points show the mean instability index values measured across replicate PCR reactions and error bars the standard deviation. For regions where multiple tissue sub-pieces were analyzed, the mean instability index across all sub-pieces is shown. Spearman's correlation showed a significant association between repeat length and instability index in (b) the cerebellar hemispheres (rho = 0.751, $p$=0.0031), (d) the pons (rho = -0.720, $p$=0.0017), (h) the motor cortex (rho = -0.769, $p$=0.0052), but not in (a) the vermis (rho = 0.018, $p$=0.97), (c) the dentate nucleus (rho = -0.184, $p$=0.53), (e) the superior cerebellar peduncles (rho = -1.00, $p$=0.083), (f) the frontopolar cortex (rho = 0.319, $p$=0.54), (g) the frontal cortex (rho = -0.515, $p$=0.087), (i) the occipital cortex (rho = -0.291, $p$=0.41), (j) the thalamus (rho = -0.552, $p$=0.10), (k) the caudate nucleus (rho = -0.539, $p$=0.17), (l) the putamen (rho = -0.667, $p$=0.083), and (m) the substantia nigra (rho = -0.800, $p$ = 0.33).

**Supplementary Figure 50: Contraction index of individual *FGF14* allele across central nervous system regions**

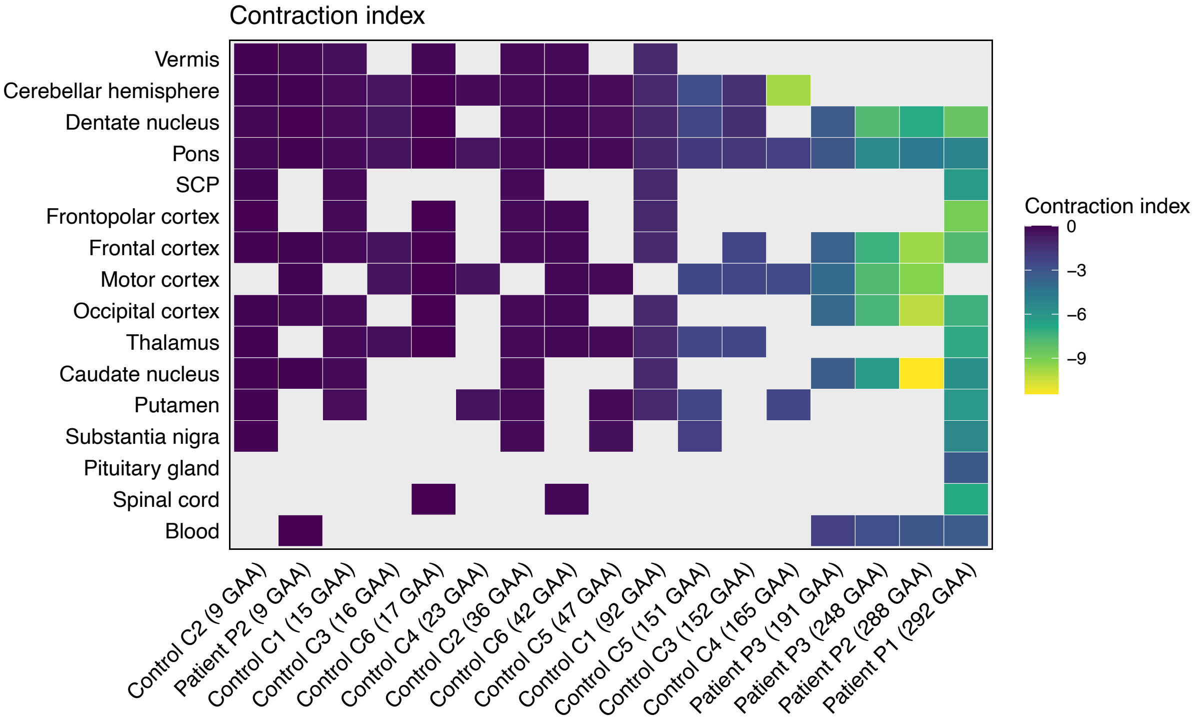


Heatmap of mean contraction indices for individual alleles across central nervous system regions and blood samples. Alleles are arranged in ascending order of length. The color of each tile represents the mean contraction index, according to the color scale shown on the right side of the heatmap. Gray tiles indicate either unavailable tissue samples for analysis or that contraction index could not be calculated. SCP: superior cerebellar peduncles.

**Supplementary Figure 51: Correlation of GAA•TTC repeat length with contraction index across multiple central nervous system regions**

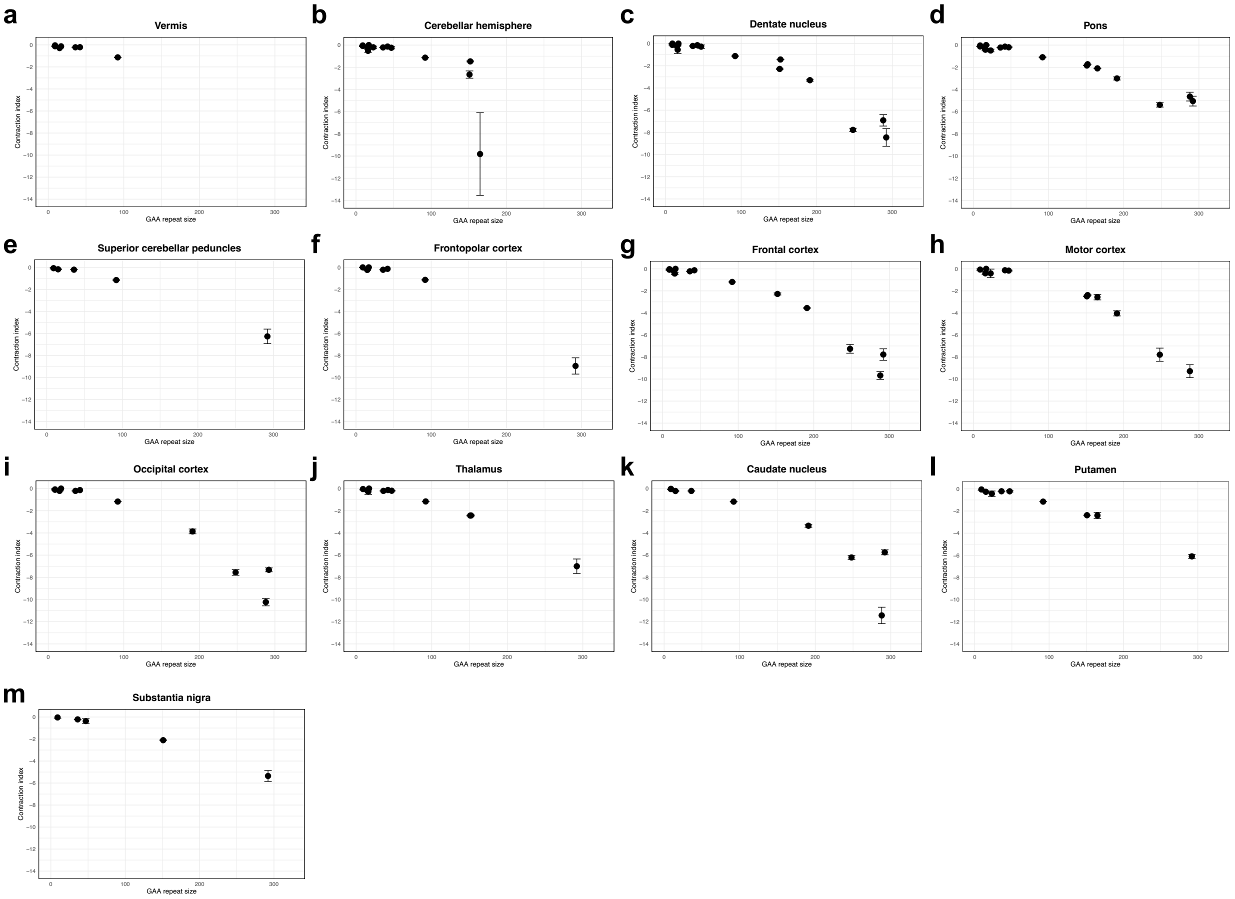


Correlation between *FGF14* GAA•TTC repeat length and contraction index across multiple central nervous system regions. Points show the mean contraction index values measured across replicate PCR reactions and error bars the standard deviation. For regions where multiple tissue sub-pieces were analyzed, the mean contraction index across all sub-pieces is shown. Regression analysis and Pearson's correlation showed a significant linear relationship between repeat length and contraction index in (a) the vermis (r = -0.93 [95% CI: -0.99 to -0.57], slope = -0.012, $p$=0.0028), (b) the cerebellar hemisphere (r = -0.73 [95% CI: -0.91 to -0.30], slope = -0.033, $p$=0.0044), (c) the dentate nucleus (r = -0.95 [95% CI: -0.98 to -0.84], slope = -0.027, $p$<0.0001), (d) the pons (r = -0.97 [95% CI: -0.99 to -0.91], slope = -0.018, $p$<0.0001), (e) the superior cerebellar peduncles (r = -0.99 [95% CI: -1.00 to -0.88], slope = -0.022, $p$=0.00083), (f) the frontopolar cortex (r = -0.98 [95% CI: -1.00 to -0.90], slope = -0.032, $p$<0.0001), (g) the frontal cortex (r = -0.96 [95% CI: -0.99 to -0.88], slope = -0.030, $p$<0.0001), (h) the motor cortex (r = -0.95 [95% CI: -0.98 to -0.81], slope = -0.030, $p$<0.0001), (i) the occipital cortex (r = -0.97 [95% CI: -0.99 to -0.88], slope = -0.031, $p$<0.0001), (j) the thalamus (r = -0.98 [95% CI: -0.99 to -0.91], slope = -0.023, $p$<0.0001), (k) the caudate nucleus (r = -0.91 [95% CI: -0.98 to -0.61], slope = -0.029, $p$=0.00071), (l) the putamen (r = -0.98 [95% CI: -1.00 to -0.90], slope = -0.020, $p$<0.0001), and (m) the substantia nigra (r = -0.99 [95% CI: -1.00 to -0.90], slope = -0.019, $p$=0.00064).

**Supplementary Figure 52: Ranking of contraction indices in central nervous system regions**

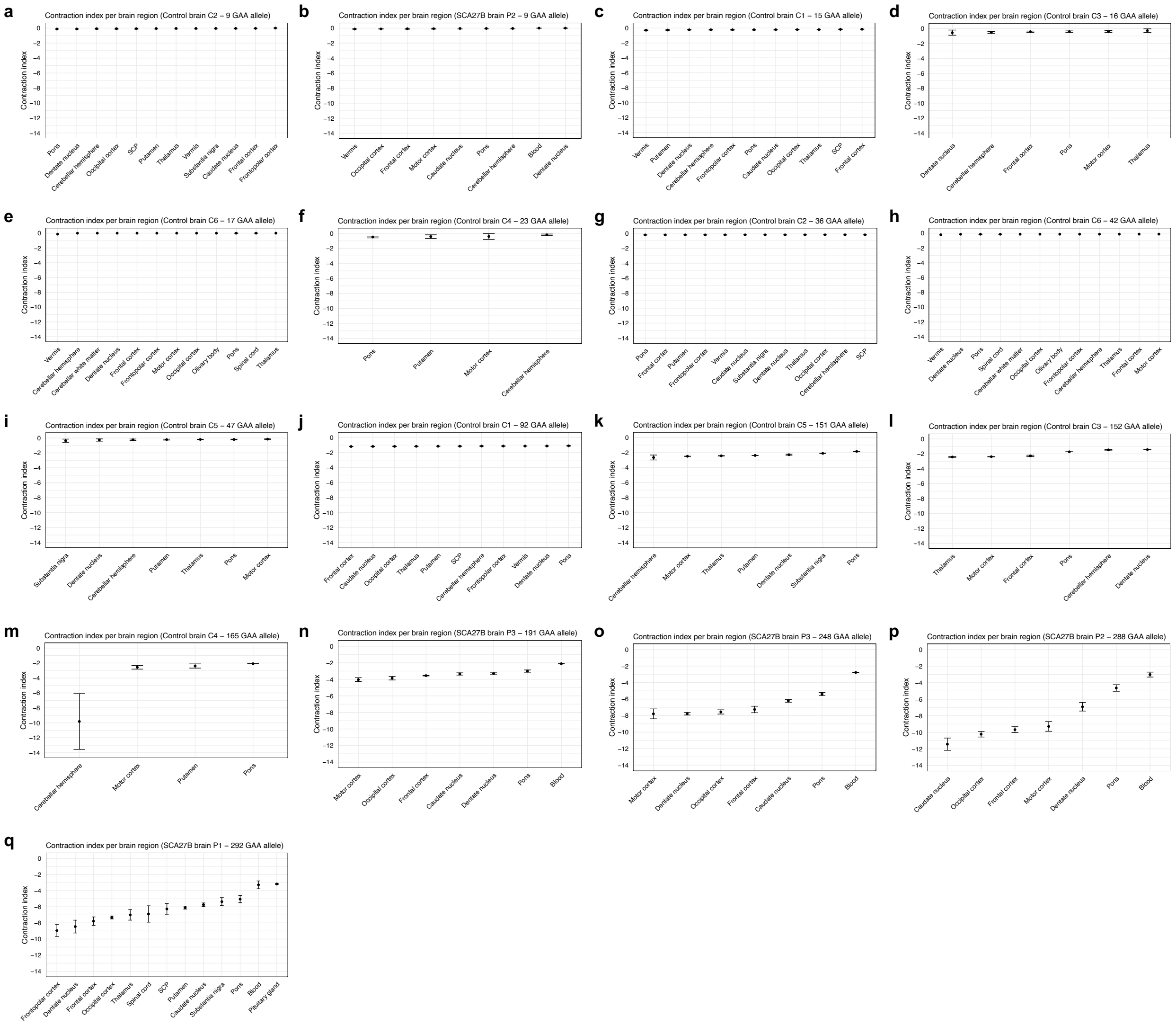


Mean contraction indices for individual alleles across central nervous system regions and blood samples. In each plot, tissues are displayed from left to right in ascending order of contraction index value. Points show the mean contraction index values measured across replicate PCR reactions and error bars the standard deviation. For regions where multiple tissue sub-pieces were analyzed, the mean contraction index across all sub-pieces is shown. Results for the longer allele of patient P1 are not shown as contraction indices could not be calculated. SCP: superior cerebellar peduncles.

**Supplementary Figure 53:** ***FGF14*** **GAA•TTC repeat length in post-mortem central nervous system tissues from SCA27B patient brain P4**

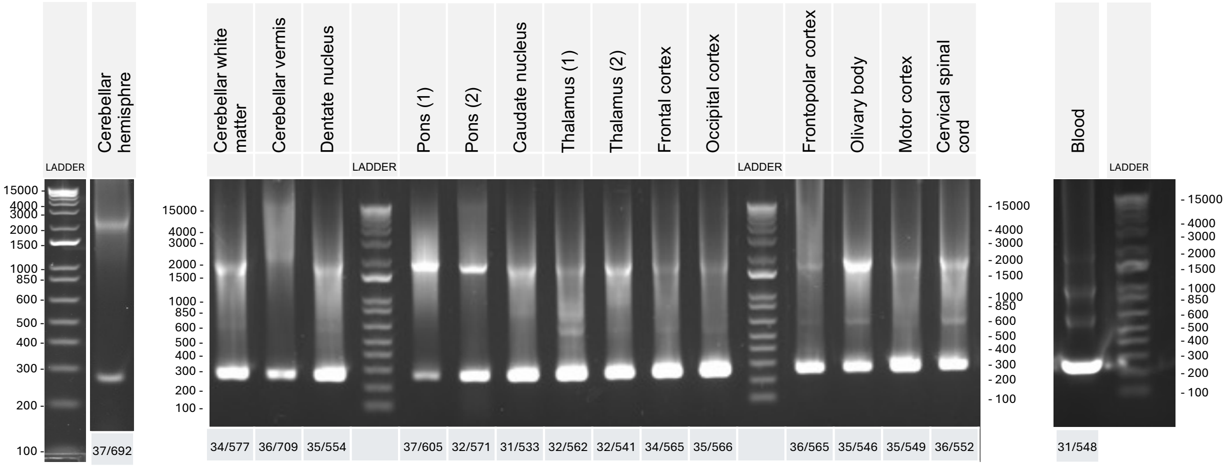


Agarose gel (1%) analysis of *FGF14* GAA•TTC repeat sizes in peripheral blood and multiple brain tissues from SCA27B patient P4. Repeat sizes for each tissue are indicated under the respective lane of the agarose gel for each tissue.

**Supplementary Figure 54: *FGF14* GAA•TTC repeat length in post-mortem central nervous system tissues from SCA27B patient brain P5**

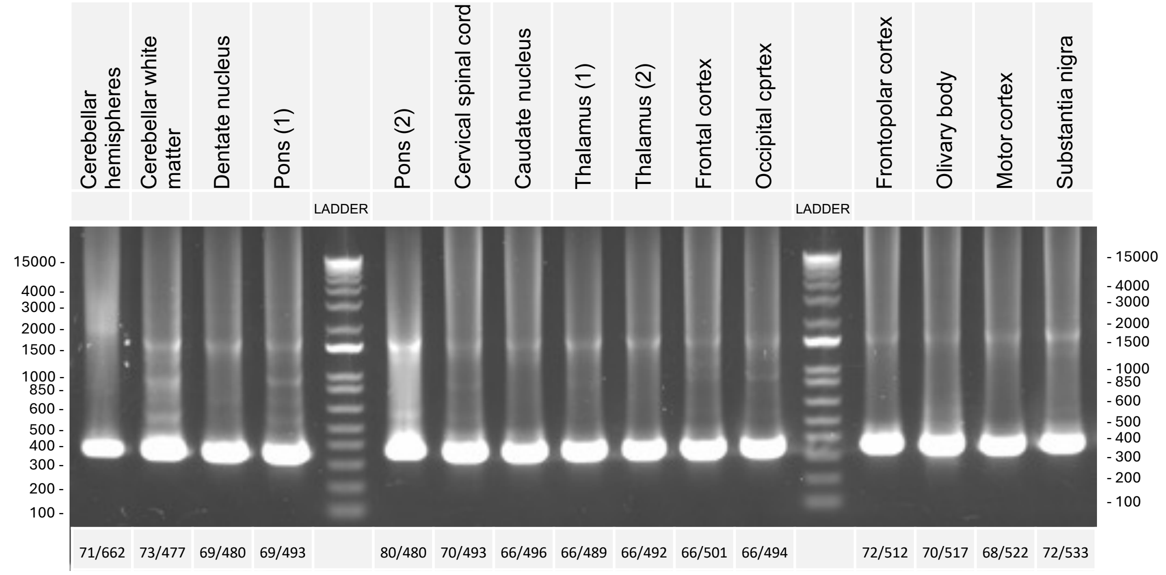


Agarose gel (1%) analysis of *FGF14* GAA•TTC repeat sizes in multiple brain tissues from SCA27B patient P5. Repeat sizes for each tissue are indicated under the respective lane of the agarose gel for each tissue.

**Supplementary Figure 55:** ***FGF14*** **GAA•TTC repeat length in post-mortem central nervous system tissues from SCA27B patient brain P6**

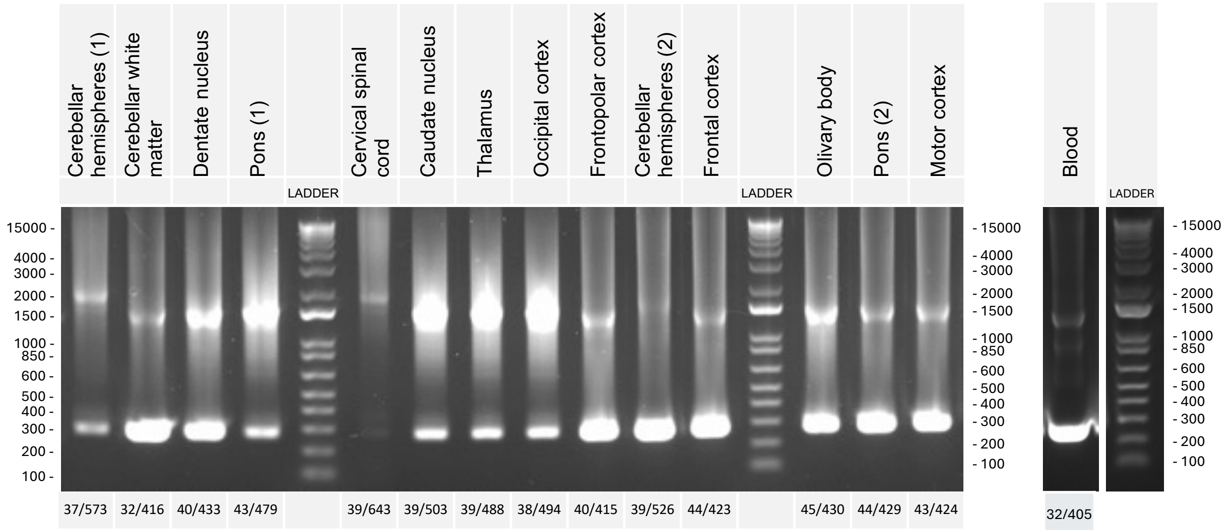


Agarose gel (1%) analysis of *FGF14* GAA•TTC repeat sizes in peripheral blood and multiple brain tissues from SCA27B patient P6. Repeat sizes for each tissue are indicated under the respective lane of the agarose gel for each tissue.

**Supplementary Tables**

**Supplementary Table 1: Number of central nervous system and blood samples analyzed in control individuals and SCA27B patients**

| Brain regions | Control brains | | | | | | SCA27B patient brains | | | | | |
|---|---|---|---|---|---|---|---|---|---|---|---|---|
| | **C1** | **C2** | **C3** | **C4** | **C5** | **C6** | **P1** | **P2** | **P3** | **P4** | **P5** | **P6** |
| | **Montreal** | | **Paris** | | | | **Montreal** | **Barcelona** | | **Paris** | | |
| Caudate nucleus | 1 | 1 | | | | | 1 | 1 | 1 | 1 | 1 | 1 |
| Cerebellar hemisphere (cortex) | 1 | 1 | 1 | 1 | 1 | 1 | 3 | 1 | 1 | 1 | 1 | 2 |
| Cerebellar vermis | 1 | 1 | | | | 1 | 2 | 1 | | 1 | | |
| Cerebellar white matter | | | | | | 1 | | | | 1 | 1 | 1 |
| Cervical spinal cord | | | | | | 1 | 3 | | | | 1 | 1 |
| Dentate nucleus | 1 | 1 | 1 | | 1 | 1 | 3 | 1 | 1 | 1 | 1 | 1 |
| Frontal cortex / middle frontal gyrus (BA8) | 1 | 1 | 1 | | | 1 | 3 | 1 | 1 | 1 | 1 | 1 |
| Frontopolar cortex / prefrontal cortex (BA9/10) | 1 | 1 | | | | 1 | 1 | | | 1 | 1 | 1 |
| Inferior olive | | | | | | 1 | | | | 1 | 1 | 1 |
| Occipital cortex | 2 | 1 | | | | 1 | 1 | 1 | 1 | 1 | 1 | 1 |
| Pituitary gland | | | | | | | 1 | | | | | |
| Pons | 2 | 1 | 1 | 1 | 1 | 1 | 3 | 2 | 2 | 2 | 2 | 2 |
| Primary motor cortex (BA4) | | | 1 | 1 | 1 | 1 | | 1 | 1 | 1 | 1 | 1 |
| Putamen | 1 | 2 | | 1 | 1 | | 1 | | | | | |
| Substantia nigra | | 1 | | | 1 | | 1 | | | 1 | 1 | |
| Superior cerebellar peduncles | 2 | 1 | | | | | 2 | | | | | |
| Thalamus | 2 | 1 | 1 | | 1 | 1 | 1 | | | 2 | 2 | 1 |
| **Peripheral tissue** | | | | | | | | | | | | |
| Blood | | | | | | | 1 | 1 | 1 | 1 | | 1 |

The number of tissue pieces analyzed for each brain region is indicated.

**Supplementary Table 2: Expansion, contraction, and instability indices of *FGF14* GAA•TTC repeat measured in peripheral blood, fibroblasts, and induced pluripotent stem cells from three patients with SCA27B**

| Patient | Sample | Allele size | Expansion index | Contraction index | Instability index |
|---|---|---|---|---|---|
| P1* | Blood | 292 | | -3.28 ± 0.49 | |
| | Fibroblasts | 295 | | -4.15 ± 0.38 | |
| | iPSC clone 4 | 293 | | -2.48 ± 0.02 | |
| | iPSC clone 5 | 295 | | -0.91 ± 0.42 | |
| | iPSC clone 6 | 291 | | -3.64 ± 0.33 | |
| | Blood | 304 | 2.57 ± 0.16 | | |
| | Fibroblasts | 304 | 4.17 ± 0.40 | | |
| | iPSC clone 4 | 303 | 3.20 ± 1.78 | | |
| | iPSC clone 5 | 304 | 2.20 ± 0.26 | | |
| | iPSC clone 6 | 315 | 10.03 ± 0.48 | | |
| P8 | Blood | 9 | 0 ± 0 | -0.10 ± 0.09 | -0.10 ± 0.09 |
| | Fibroblasts | 9 | 0 ± 0 | 0 ± 0 | 0 ± 0 |
| | iPSC clone 1 | 9 | 0 ± 0 | -0.05 ± 0.08 | -0.05 ± 0.08 |
| | iPSC clone 2 | 9 | 0.02 ± 0.01 | -0.09 ± 0.01 | -0.09 ± 0.01 |
| | iPSC clone 3 | 9 | 0 ± 0 | -0.04 ± 0.03 | -0.04 ± 0.03 |
| | iPSC clone 5 | 9 | 0 ± 0 | 0 ± 0 | 0 ± 0 |
| | Blood | 508 | | | |
| | Fibroblasts | 518 | | | |
| | iPSC clone 1 | 597 | | | |
| | iPSC clone 2 | 607 | | | |
| | iPSC clone 3 | 527 | | | |
| | iPSC clone 5 | 525 | | | |
| P9 | Blood | 16 | 0.01 ± 0.01 | -0.29 ± 0.02 | -0.29 ± 0.02 |
| | Fibroblasts | 16 | 0 ± 0 | -0.12 ± 0.01 | -0.12 ± 0.01 |
| | iPSC clone 1 | 16 | 0 ± 0 | -0.12 ± 0.003 | -0.12 ± 0.003 |
| | iPSC clone 2.1 | 16 | 0.01 ± 0.01 | -0.18 ± 0.04 | -0.18 ± 0.04 |
| | iPSC clone 2.2 | 16 | 0.01 ± 0.01 | -0.16 ± 0.02 | -0.16 ± 0.02 |
| | iPSC clone 4 | 16 | 0 ± 0 | -0.17 ± 0.03 | -0.17 ± 0.03 |
| | Blood | 389 | | | |
| | Fibroblasts | 409 | | | |
| | iPSC clone 1 | 402 | | | |
| | iPSC clone 2.1 | 411 | | | |
| | iPSC clone 2.2 | 390 | | | |
| | iPSC clone 4 | 402 | | | |

Expansion, contraction, and instability indices are expressed as the mean ± standard deviation of triplicate measurements.

*Expansion and instability indices could not be calculated for the short allele, and contraction and instability indices could not be calculated for the long allele due to the insufficient size difference between both alleles.
